# The effect of carotid atherosclerosis and revascularization on cognition and dementia: a systematic review and meta-analysis

**DOI:** 10.64898/2026.11.09.26367363

**Authors:** Camiel V. J. Box, Qingling Yu, Rosa Stroom, Hence Verhagen, Dominic P.J. Howard, Christa Niehot, M. Kamran Ikram, Frank J. Wolters, Daniel Bos

## Abstract

**Background:** Carotid artery atherosclerosis may accelerate cognitive decline and increase dementia risk, but evidence is fragmented and inconsistent, and benefit of revascularization on cognitive outcomes uncertain.

**Methods:** We systematically searched Medline, Embase, Web of Science, the Cochrane Central Register, and Google Scholar for articles published until 11 November 2024, reporting change in cognition or dementia incidence in relation to carotid artery intima-media thickness, plaque, stenosis, and revascularization procedures. We excluded studies that investigated people with dementia, lacked a reference group, or had post-intervention follow-up <30 days. We additionally included new data from the Rotterdam Study, and results from the recent CREST-2 trial. Two independent reviewers extracted study data. Random-effects meta-analyses were performed if data allowed. Risk of bias was assessed using the Newcastle-Ottawa Scale, Cochrane Risk of Bias, and ROBINS-I.

**Results:** Of 5592 records identified, 97 studies were eligible, including 43 observational studies (N=100,326) and 54 intervention studies (N=15788, 6 randomized controlled trials). In observational studies, mostly among stroke-free individuals, intima-media thickness was associated with higher dementia risk (hazard ratio per 1-SD increase: 1.07, 95%CI 1.01–1.13; I^2^=24.8%; 5 studies), as was presence of 50-100% asymptomatic steno-occlusive disease (hazard ratio: 1.48, 95%CI 1.06–2.07; I^2^=0.0%, 3 studies). Cognitive outcomes were too heterogeneous for meta-analysis, but 11/20 observational studies reported a significant association between carotid atherosclerosis and global cognition. In non-randomized intervention studies, carotid revascularization improved global cognition (standardized mean difference: 0.73, 95%CI 0.37–1.09; I^2^=89.8%), but all studies were at high risk of bias, and heterogeneity was only partially explained by intervention type and study characteristics in meta-regression. Suggested benefit of stenting on cognition among Chinese patients (Standardized effect: 0.86 [0.57-1.16], 197 patients; and 1.09 [0.64-1.53], 90 patients, 2 trials) was not observed in the larger CREST-2 trial (Standardized effect: 0.03 [−0.09 to 0.15] and 0.02 [−0.09 to 0.14]), nor in post-hoc analyses of cognition in the ACAS trial and long-term dementia risk in ACST-1 (HR: 0.98 [0.75-1.28]).

**Conclusions:** Carotid atherosclerosis is associated with increased risk of dementia. Current evidence does not support carotid revascularization for prevention of cognitive decline or dementia.

## Introduction

Carotid artery atherosclerosis is a major cause of ischemic stroke.^1^ Carotid revascularization by surgical endarterectomy or endovascular stenting is an important part of secondary stroke prevention in patients with ≥50% symptomatic carotid stenosis.^2^ Yet, whether revascularization is also beneficial for primary prevention in asymptomatic patients (i.e. those who did not suffer a stroke or TIA caused by significant carotid stenosis) is debated,^3^ especially in light of mixed results from the recent ECST-2 and CREST-2 trials.^4,5^ Benefit of revascularization on brain health, beyond clinical stroke alone, may tip the risk-benefit scale in favor of intervention. This broadens the scope from stroke prevention alone, to include cognitive decline and dementia.

Carotid artery atherosclerosis is implicated in the etiology of dementia.^6,7^ While partly mediated by stroke occurrence, the effect of carotid stenosis may be amplified by chronic hypoperfusion, (micro)embolization and covert cerebrovascular injury, and exhausted cerebrovascular reactivity. However, it remains uncertain whether some of the previously reported associations are indeed causal, and whether described effects of stenosis on subclinical brain health measures translate into clinical cognitive impairment and dementia.^3,8^ Previous systematic reviews mainly included cross-sectional studies, which limits causal inference, and focused solely on the association of carotid atherosclerosis with cognition and not dementia.^6,7^ Importantly, any potential effect of revascularization on cognition and dementia depends on a causal relationship. A prior systematic review found no evidence of cognitive improvement after revascularization but included few studies with reference subjects.^9^

We present a comprehensive overview of evidence on the effect of carotid atherosclerosis and revascularization procedures on cognitive decline and dementia, including prospective observational studies and intervention studies with reference subjects.

## Methods

This review was reported according to the Preferred Reporting Items for Systematic Reviews and Meta-analyses (PRISMA) guidelines (Table S1) and registered with the International Prospective Register of Systematic Reviews (PROSPERO) as ID CRD420251039762.^10^

### Search strategy

A search strategy was developed by an information specialist (CN) in cooperation with the lead author (CVJB). The search was developed in Embase.com, optimized for sensitivity and then translated to other databases following the method as described by Bramer et al.^11^ We searched Medline ALL via Ovid, Embase.com, Web of Science Core Collection and the Cochrane Central Register of Controlled Trials via Wiley (1992-present). Additionally, a search was performed in Google Scholar from which the 100 highest-ranked references were downloaded using the software Publish or Perish.^12^ The search strategies for Medline and Embase used relevant thesaurus terms from Medical Subject Headings (MeSH) and Emtree, respectively. In all databases, terms were searched in titles, abstracts and author keywords. The search contained terms for 1) Carotid Stenosis and 2) Cognition disorders and dementia. Terms were combined with Boolean operators AND and OR and proximity operators were used to combine terms into phrases. The full search strategies of all databases are available in the supplementary materials. The searches in Embase and Web of Science were limited to exclude conference papers. In all databases, animal only articles were excluded from the search results. No study registries were searched, but Cochrane CENTRAL retrieves the contents of ClinicalTrials.gov and World Health Organization’s International Clinical Trials Registry Platform. No authors or subject experts were contacted and no extra unindexed journals in the field were browsed.

The references were imported into EndNote and duplicates were removed by an information specialist (CN) using the method as described by Bramer et al.^13^ The searches were conducted on 18 November 2024 and updated for evidence from randomized controlled trials on 9 October 2026. We also added new data from the prospective, population-based Rotterdam Study on carotid artery stenosis, in addition to previously published IMT results.

### Study selection

Two reviewers (CVJB and RS) independently screened references by title and abstract. In case of disagreement between reviewers, consensus was reached through a consensus meeting. Covidence systematic review software, Veritas Health Innovation, Melbourne, Australia, was used for study screening and selection. We included published research articles in any language that described longitudinal observational studies investigating the effect of extracranial carotid atherosclerosis measures (exposure) on dementia or cognition (outcome), or interventional studies investigating the effect of revascularization methods on dementia or cognition, with human adults as participants. We excluded case studies/series, conference abstracts, commentaries, reviews, and letters to the editor. Studies were further excluded if they investigated people with prior dementia, lacked a reference group, or had post-intervention follow-up less than 30 days. Full-text assessment of inclusion criteria was performed by at least two independent reviewers (CVJB, RS and QY), with decisions made through consensus discussion with a third reviewer (DB or FJW) in case of disagreement.

### Data extraction

Two independent reviewers (CVJB and QY) used prespecified data collection forms specific for the study design to extract data from selected studies. We collected data on the study (calendar year of conduct, country, funding source), study population (inclusion criteria, sample size and selection, age, sex, follow-up), methodology (exposure/intervention and outcome assessment, statistical analysis, confounder adjustments), and results. For the interventional studies we further collected data on randomization and blinding, and for the outcomes in the non-randomized interventional studies specifically, we extracted pre-and postintervention mean cognition scores including standard deviations for both groups.^14^ Cognitive outcomes were grouped into different domains, including global cognition, attention, memory, processing speed, executive functioning, language, motor speed and visuospatial (Table S2). In case of multiple analyses, we included those that were best controlled for potential confounders, had longest follow-up, or least exclusion.

### Quality assessment

The methodological quality of the included studies was assessed independently by two assessors (CVJB and QY) with discrepancies resolved through consensus. We used a modified Newcastle Ottawa Scale (NOS) for cohort studies, the Risk Of Bias In Non-Randomized Studies of Interventions tools (ROBINS-I-V2) for non-randomized interventional studies, and the Cochrane Risk of Bias tools 2.0 (RoB 2) for randomized controlled trials.

### Statistical analysis

The extracted results were described narratively and combined in pooled analyses if exposure and outcome were sufficiently comparable for at least three studies. Internal meta-analysis of within-study results was used to harmonize subgroup data if necessary. For the non-randomized intervention studies, cognition change scores were calculated for each group, and the between-group difference in change (Δ Mean) was computed in intervention and reference group using the following formula:

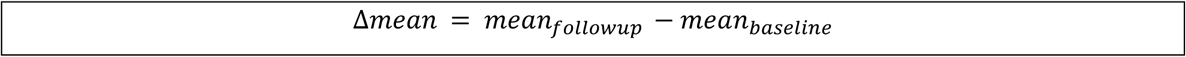

The standard deviation of the difference in change scores (Δ SD) was estimated using the following formula:

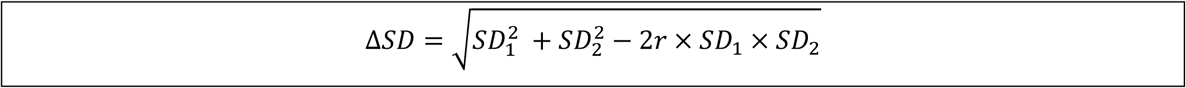

SD_1_ and SD_2_ represent the standard deviations of the change scores in the baseline and follow-up, respectively.^14^ Standardized mean differences (SMD) were calculated using Hedges’ g with small-sample correction. For the randomized control studies, standardized cognitive effects were pooled. CREST-2 reported baseline-adjusted standardized treatment effects, for other trials reporting group-specific scores, Hedges’ g was calculated from post-intervention means and standard deviations to obtain comparable standardized between-group effects. Effect estimates were pooled using a random-effects model. Forest plots were used to visualize individual and pooled effect estimates. ORs and HRs were combined in meta-analyses if similar exposures and outcomes were used. Between-study heterogeneity was quantified using the I² statistic.

We performed meta regression to investigate whether study-level characteristics could explain heterogeneity in the pooled estimates for global cognition. Covariates were prespecified according to clinical plausibility and anticipated data availability, including mean age, sex distribution, follow-up duration, prior symptoms in the intervention group, and stenosis prevalence in the reference group. For subgroup analyses, studies were stratified by intervention type (carotid endarterectomy [CEA] vs carotid artery stenting [CAS]), symptom status in the intervention group (symptomatic vs. asymptomatic), and stenosis status in the reference group (with vs. without stenosis).

Several sensitivity analyses were performed. First, we examined alternative assumptions regarding the within-study correlation used for effect-size aggregation. Second, robust variance estimation was applied to account for dependence among multiple neuropsychological outcomes within studies. Third, the influence of extreme values was assessed by trimming the upper and lower 2.5%, 5%, and 10% of effect sizes.

Publication bias was assessed using Funnel plots and Egger’s regression tests, with cluster-robust variance estimation used to account for the inclusion of multiple effect sizes per study. Alpha was set to 0.05. All statistical analyses were performed in R, using the “meta” and “metafor” packages.

### Data availability

Data on study screening and inclusion, as well as extracted data from included studies is available upon request.

## Results

### Study selection and population

Of 5591 records identified, 3040 remained after duplicate removal. Following title and abstract screening, 232 (7.6%) qualified for full-text review (Figure S1). Of 14 studies the full-text could not be retrieved, and 123 studies were excluded after full-text review, mostly because they were cross-sectional (n=50), lacked a longitudinally observed reference group (n=25) or investigated different exposures or outcomes (n=18). We identified 95 eligible studies and then additionally included new analyses from the Rotterdam Study, which we published separately, and the recently published cognitive results from the CREST-2 randomized trial. In total, we included 43 observational cohort studies (100,326 participants), 48 non-randomized intervention studies (10029 participants), and 6 randomized controlled trials (5759 participants). Observational cohort studies mostly included general-population participants, alongside memory-clinic populations and a few post-stroke, with a large concentration in Europe (20) and the USA (13), with fewer in East Asia (8) and South America (2). The interventional studies included symptomatic and asymptomatic carotid stenosis patients with diverse reference groups, including healthy controls, other surgery patients and carotid stenosis patients who did not undergo intervention, and were mainly from Europe (23), Asia (22), and the USA (9). The methodological assessment of the observational cohort studies showed mostly good quality (36), and few of fair (1) or poor (5) quality (Table S3). Among the randomized controlled trials, three studies were judged as having some concerns regarding risk of bias and two as having a high risk of bias, while all non-randomized intervention studies were at critical risk of bias (Tables S4–S5).

### Observational cohort studies

Observational cohort studies defined carotid atherosclerosis by IMT, plaque, or stenosis, assessed through ultrasound only (36), ultrasound or CT-angiography (2), CT-angiography only (2), ultrasound or MR-angiography (1) or both ultrasound and MRI (1).^15–57^ Of 43 studies, 26 reported associations with cognition and 19 with dementia. Of 19 dementia studies,^15–32^ 12 excluded participants with a history of stroke (i.e. purely asymptomatic atherosclerosis),^17–19,21–23,27,29–32,57^ whereas three included <10% participants with stroke,^20,25,28^ one had 72.3%,^16^ and two studies did not report it.^15,26^ Sample size ranged from 74 to 14,348 participants (median: 600), with follow-up duration ranging from 1 to 24 years (median: 5.5 years). In 10/19 studies all included participants at baseline were also included in the longitudinal analyses (even if lost to follow-up), in the other nine an average of 21% (median 13%) were not included. The number of participants with dementia diagnosed during follow-up ranged from 21 to 2,224 (median 105).

A 1-SD increase in IMT related to a 1.07-fold (95%CI: 1.01–1.13) higher risk of all-cause dementia (5 studies; Figure 1).^20,23,28,30,31^ This effect was consistent when IMT was analyzed dichotomously (pooled HR: 1.39 [95%CI: 1.13–1.71]; I^2^=57%; 7 studies), or as highest versus lowest quantile (pooled HR: 1.24 [1.11–1.40], I^2^=15%, 5 studies; Figure S2). The presence of carotid plaque had a somewhat smaller, not statistically significant effect on risk of dementia (pooled HR: 1.08 [0.99–1.18]; I^2^=0%, 7 studies). Six studies investigated the effect of carotid steno-occlusive disease on dementia,^16–18,24,29,57^ with four applying a 50% threshold.^16,24,29,57^ Presence of ≥50% stenosis was associated with a higher risk of dementia (pooled HR: 1.48, 95%CI: 1.06–2.07; I^2^=0%, 3 studies; Figure 1).^24,29,57^ Of the three studies not included in the meta-analysis, Zheng et al. did not provide the necessary relative risks, but dementia occurred significantly more often in people with ≥50% stenosis than those without (13/20 vs 10/54).^16^ Barnes et al.^17^ used a cut-off of ≥25%, and Li et al.^18^ provided crude risk estimates for mild (0-29%), moderate (30-69%), and severe (≥70%) stenosis among MCI patients. Including the latter two in a sensitivity analysis of any ≥25-30% stenosis yielded a pooled HR of 1.71(95% CI: 1.00–2.92, I^2^=66.9%, 5 studies; Figure S2). Two studies assessed the effect of ≥70% stenosis on dementia.^18,57^ Li et al,^18^ reported a crude HR of 11.64 (2.50–54.29) comparing ≥70% stenosis to 0-29%. Box et al,^57^ showed that the confounder-adjusted association of ≥70% stenosis on dementia depended strongly on the inclusion of 100% stenosis (compared to <70% stenosis, HR for 70-100% stenosis: 1.81 [0.91–3.59]; and for 70-99% stenosis: 1.07 [0.39–2.91]).

**Figure 1:**
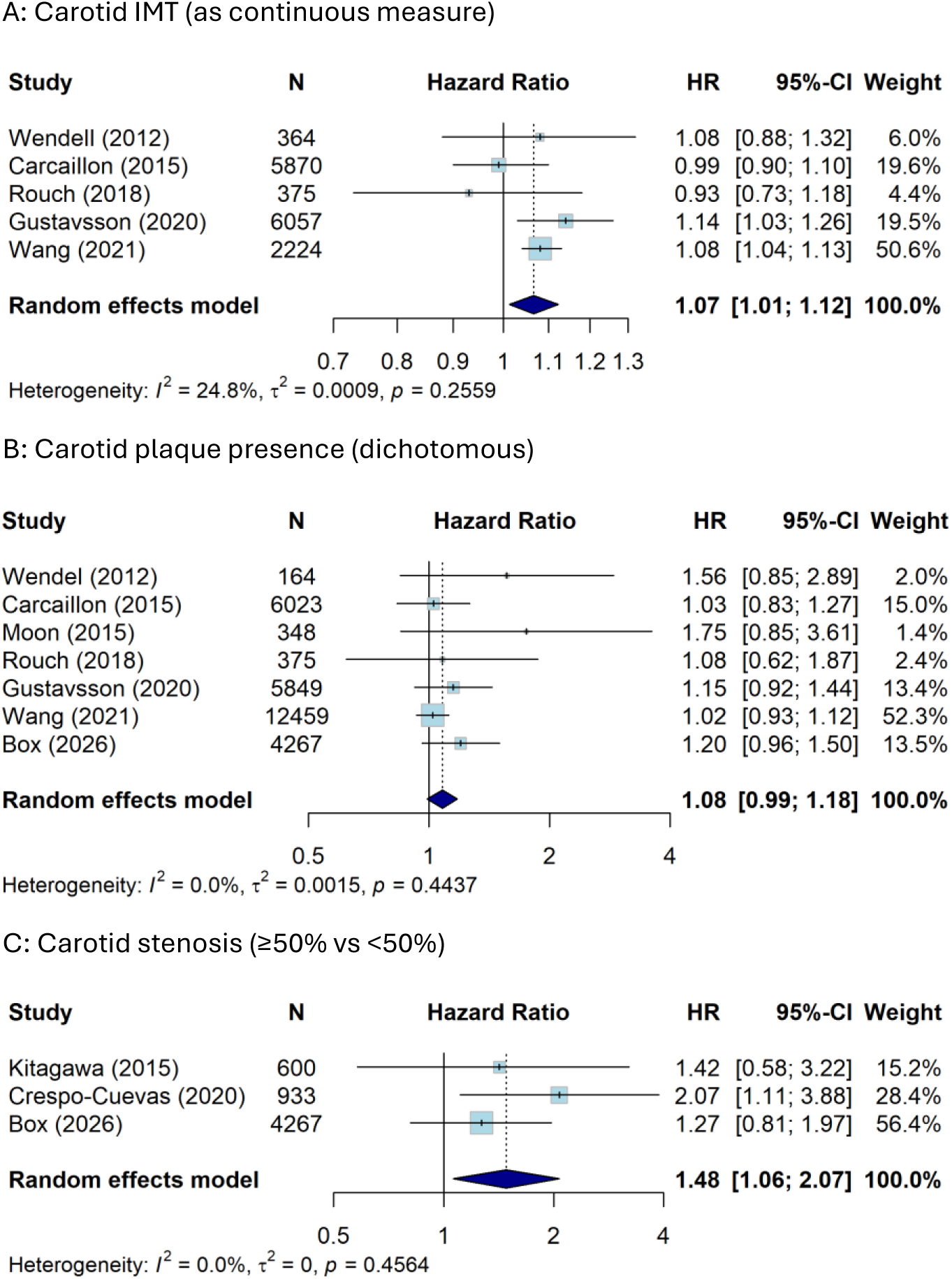
Association of carotid atherosclerosis with all-cause dementia. Legend: N = number of participants; HR = hazard ratio; CI = confidence interval, IMT = intimal medial thickness. A: The HR for the study by Wendell (2012)^20^ were adjusted to represent IMT per SD instead of IMT per 0.1mm. B: The HR for the studies by Carcaillon (2015)^23^ and Wendell (2012)^20^ were based on internal meta-analyses of multiple categories for plaque presence in order to get a dichotomous measure of plaque presence. The study by Moon (2015)^25^ used logistic regression to assess the combined outcome of MCI and dementia. C: The study by Crespo-Cueva (2020)^29^ was performed in people from the general population with moderate to high vascular risk and used a composite of mild cognitive impairment and dementia as outcome. The study by Kitagawa (2015)^24^ was performed in an out-patient clinic population with at least one vascular risk factor.

Results were similar between studies with and without prior stroke. Multiple studies reported separate estimates for the effects on clinical Alzheimer’s dementia.^15,20,22–24,30,32^ In studies that directly compared effects on all-cause dementia and clinical Alzheimer’s dementia, estimates were broadly similar.^15,20,22–24^ However, across all available studies, meta-analysis suggested a significant association of dichotomized IMT on Alzheimer’s dementia (HR: 1.80 [1.08–2.99]; I^2^=69.0%; 3 studies; Figure S3), but not of continuously modelled IMT (HR: 0.99 [0.90–1.10]; I^2^=0.0%; 3 studies), highest vs lowest quantile of IMT (HR: 1.03 [0.67–1.56]; I^2^=65.9%; 3 studies), or plaque presence (HR: 1.05 [0.91–1.22]; I^2^=35.6%; 5 studies). Three studies reported on the effects on vascular or mixed type dementia.^15,23,30^ Taken together, these found a slightly higher association of IMT (pooled HR per SD: 1.19 [0.96–1.48]; I^2^=56.7%; 2 studies) and plaque presence (pooled HR: 1.37 [1.05–1.79]; I^2^=0.0%; 3 studies) with vascular/mixed type dementia than with all-cause dementia.

Of 26 longitudinal cohort studies examining the association of carotid atherosclerosis with cognition,^18,33–48,50–57^ four included only acute stroke patients.^35,37,51,56^ A wide variety of cognitive tests were used, both for global cognition (e.g. Mini-Mental State Examination [MMSE], Montreal Cognitive Assessment [MoCA], and Cambridge Cognitive Examination [CAMCOG]) and specific cognitive domains (e.g. California Verbal Learning Test, Benton Visual Retention Test, Rey Complex Figure Test, Trail Making Test (TMT), digit span, verbal fluency, and Boston Naming Test). Several studies also used composite scores combining several measures. Due to the large methodological heterogeneity, we were unable to perform meta-analyses. Instead, we narratively summarize the findings, with results presented by outcome in Table 4. Sample sizes ranged from 30 to 14,048 participants (median 867), with follow-up durations from 3 months to 13 years (median 3.7). Attrition was higher than in dementia studies, with 7/26 including all participants and the remaining 19 averaging 26% attrition (median 23%).

Of 12 studies investigating the association of IMT with global cognition,^34–37,39,40,42,44,50,52,53,55^ eight found a significant negative association,^34,35,40,42,44,52,53,55^ and four did not.^36,37,39,50^ Studies reporting a negative association generally had a larger sample size (median 1720 vs. 538 participants; p=0.048). For carotid plaque, 2/6 studies found a significant negative association with global cognition,^35,46^ direction of effect in two other studies was consistent, but not statistically significant,^40,42^ while two other studies found no effect.^55,57^ Four of nine studies on stenosis found a significant negative effect on global cognition,^18,34,38,43^ though two of these carried a high risk of bias due to absent or limited confounder adjustments,^18,43^ and one showed inconsistent results by side of stenosis.^34^ Of the four studies including only acute stroke patients, only one found a significant association of IMT and plaque with cognition, though it did not find this for stenosis.^35^

Evidence on specific cognitive domains is detailed in Table S6. Methods and results were heterogeneous. Notably, none of six studies assessing processing speed found a significant association,^33,41,42,48,49,57^ while four of ten studies investigating executive functioning did.^39,42,45,48,51–55,57^

### Randomized controlled trials

Of the six randomized controlled trials,^58–63^ five compared carotid revascularization with medical therapy for cognitive outcomes,^58–61,63^ and one post-hoc evaluated incident dementia (Table 2).^62^ In 1,601 participants of the Asymptomatic Carotid Surgery Trial, CEA did not reduce risk of incident dementia within 20 years from baseline, compared to medical treatment (HR: 0.98 [0.75–1.28]).^62^ The CREST-2 randomized trails, including 2165 participants with ≥70% asymptomatic carotid stenosis, found no significant improvement in composite cognition with either CEA or CAS and best medical treatment compared with only best medical treatment (adjusted between-group differences: 0.03 [−0.09 to 0.15] and 0.02 [−0.09 to 0.14], respectively).^63^ In the CREST-H substudy, revascularization also did not improve 1-year composite cognitive scores among participants with reduced baseline cognition and hemodynamic impairment.^64^ Similarly, among 1,662 U.S. participants of the Asymptomatic Carotid Atherosclerosis Study trial, 5-year change in MMSE in post-hoc analysis did not differ significantly between CEA and medical therapy (p>0.05; effect estimate not reported).^58^ Two smaller trials in China assessed effect of CAS on cognitive change. (standardized effect: 0.86 [0.57-1.16], 197 patients; and 1.09 [0.64-1.53], 90 patients, respectively). ADDIN EN.CITE ^60,61^ Three of the four studies were eligible for meta-analysis, contributing four comparisons due to the separate estimates for CAS and CEA from the CREST-2 trial. The pooled standardized cognitive effect was 0.47 (95% CI −0.07 to 1.01), with large heterogeneity (I²=93.8%) due to the difference between estimates from CREST-2 compared with the earlier two RCTs (Figure 2B). With Hartung–Knapp adjustment, the point estimate was unchanged, while the 95% CI widened to −0.41 to 1.35 (Figure S5). Finally, among 41 patients with carotid artery occlusion and hemodynamic impairment, ^62^ ADDIN EN.CITE <EndNote><Cite><Author>Halliday</Author><Year>2022</Year><RecNum>359</RecNum><Displ ayText><style face=“superscript”>62</style></DisplayText><record><rec-number>359</rec-number><foreign-keys><key app=“EN” db-id=“2zwpf0ff0swtz6ep0xrvaxdl9rff0rdze0xt” timestamp=“1777454622”>359</key></foreign-keys><ref-type name=“Journal Article”>17</ref-type> <contributors> <collab> <author>Halliday, A.</author> <author>Sneade, M.</author> <author>Bjorck, M.</author><author>Pendlebury, S. T.</author> <author>Bulbulia, R.</author> <author>Parish, S.</author> <author>Llewellyn-Bennett, R.</author> <author>Pan, H.</author> <author>Whiteley, W.</author> <author>Gottsater, A.</author></collab></contributors> <titles> <title>Effect of Carotid Endarterectomy on 20 Year Incidence of Recorded Dementia: a Randomised Trial</title><secondary-title>Journal of vascular surgery</secondary-title></titles> <periodical><full-title>Journal of vascular surgery</full-title></periodical> <pages>1789</pages> <volume>75</volume> <number>5</number> <dates><ye ar>2022</year></dates> <urls><related-urls> https://www.cochranelibrary.com/central/doi/10.1002/central/CNr02421462/full</related-urls></urls><electronic-resource-num>10.1016/j.jvs.2022.03.023</electronic-resource-num></record></Cite></EndNote> extracranial-intracranial bypass was not superior to best medical therapy in improving 2-year composite cognitive scores (point estimate 0.02 [-0.5 to 0.5]).^59^ ADDIN EN.CITE <EndNote><Cite><Author>Marshall</Author><Year>2014</Year><RecNum>341</RecNum><DisplayText><style face=“superscript”>59</style></DisplayText> <record><rec-number>341</rec- number><foreign-keys><key app=“EN” db-id=“2zwpf0ff0swtz6ep0xrvaxdl9rff0rdze0xt” timestamp=“1777454622”>341</key></foreign-keys><ref-type name=“Journal Article”>17</ref-type> <contributors> <collab> <author>Marshall, R. S.</author> <author>Festa, J. R.</author> <author>Cheung, Y. K.</author> <author>Pavol, M. A.</author> <author>Derdeyn, C. P.</author> <author>Clarke, W. R.</author> <author>Videen, T. O.</author> <author>Grubb, R. L.</author> <author>Slane, K.</author> <author>Powers, W. J.</author> <author>Lazar, R. M.</author></collab></contributors> <titles> <title>Randomized Evaluation of Carotid Occlusion and Neurocognition (RECON) trial: main results</title><secondary-title>Neurology</secondary-title></titles> <periodical><full-title>Neurology</full-title></periodical><pages>744-751</pages> <volume>82</volume> <number>9</number> <dates> 2014</dates><accession-num>24477109</accession-num> <urls><related-urls> http://dx.doi.org/10.1212/wnl.0000000000000167http://www.ncbi.nlm.nih.gov/entrez/query.fcgi?holding=inleurlib_fftCamp;cmd=RetrieveCamp;db=PubMedCamp;dopt=CitationCamp;list_uids=24477109https://ovidsp.ovid.com/ovidweb.cgi?T=JSCamp;CSC=YCamp;NEWS=NCamp;PAGE=fulltextCamp;D=med11Camp;DO=10.1212</related-urls></urls></record></Cite></EndNote>

**Figure 2:**
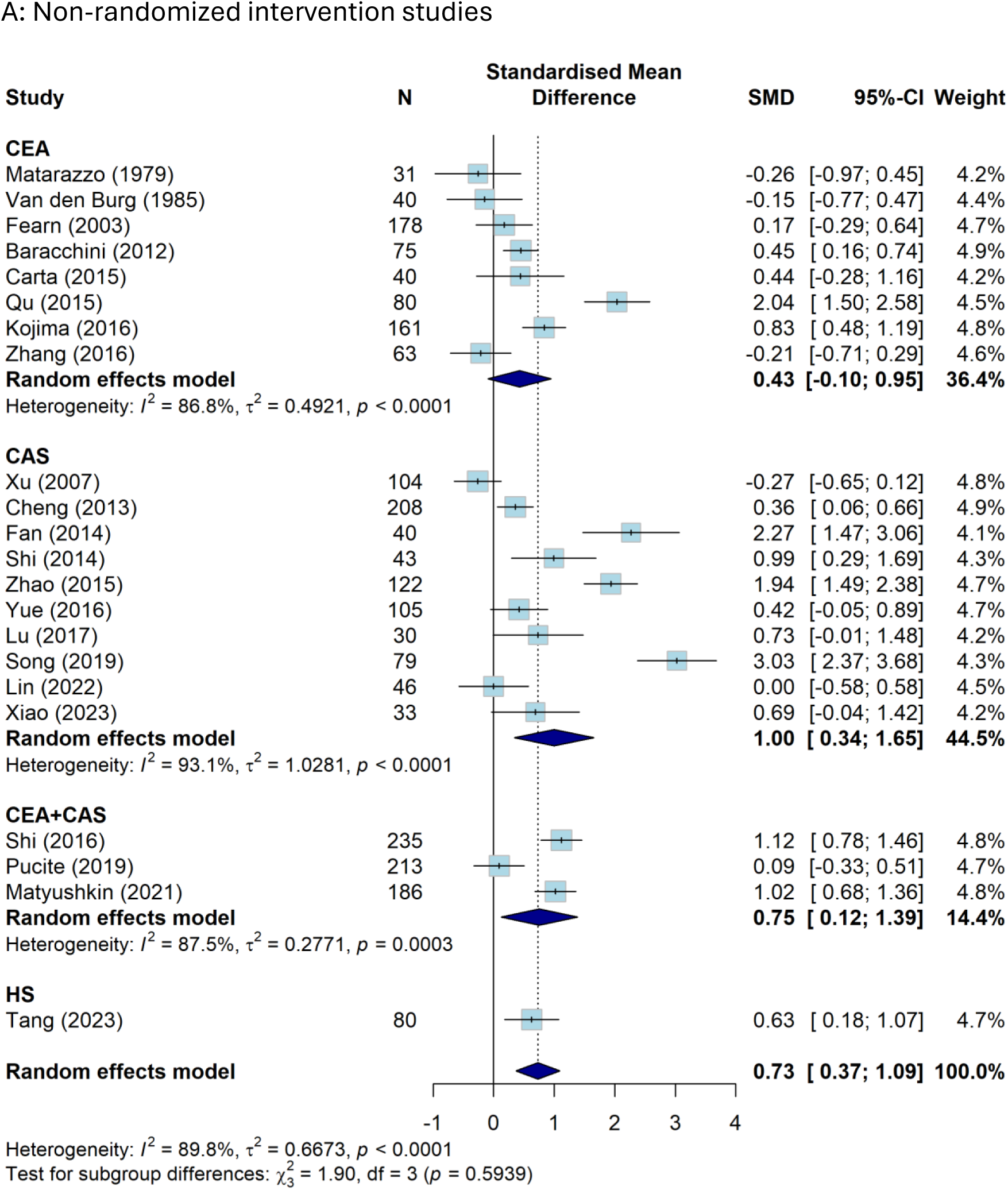
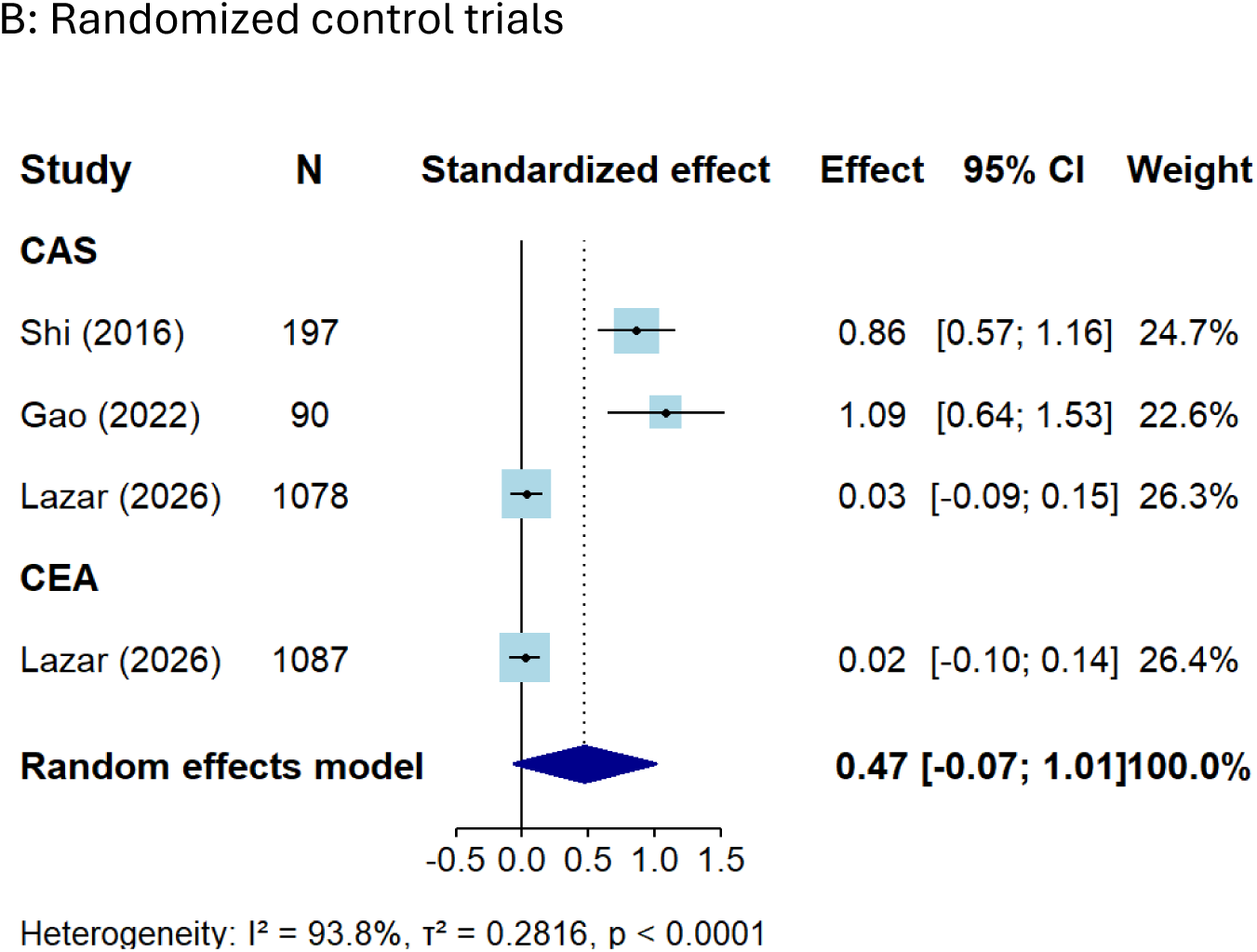
Effect of CEA/CAS on change in global cognition. Legend: N = number of participants; SMD = standardized mean difference; CI = confidence interval; CEA = carotid endarterectomy; CAS = carotid artery stenting; CEA+CAS = mixed intervention group including participants who underwent either CEA or CAS; HS = hybrid surgery.

**Table 1:**
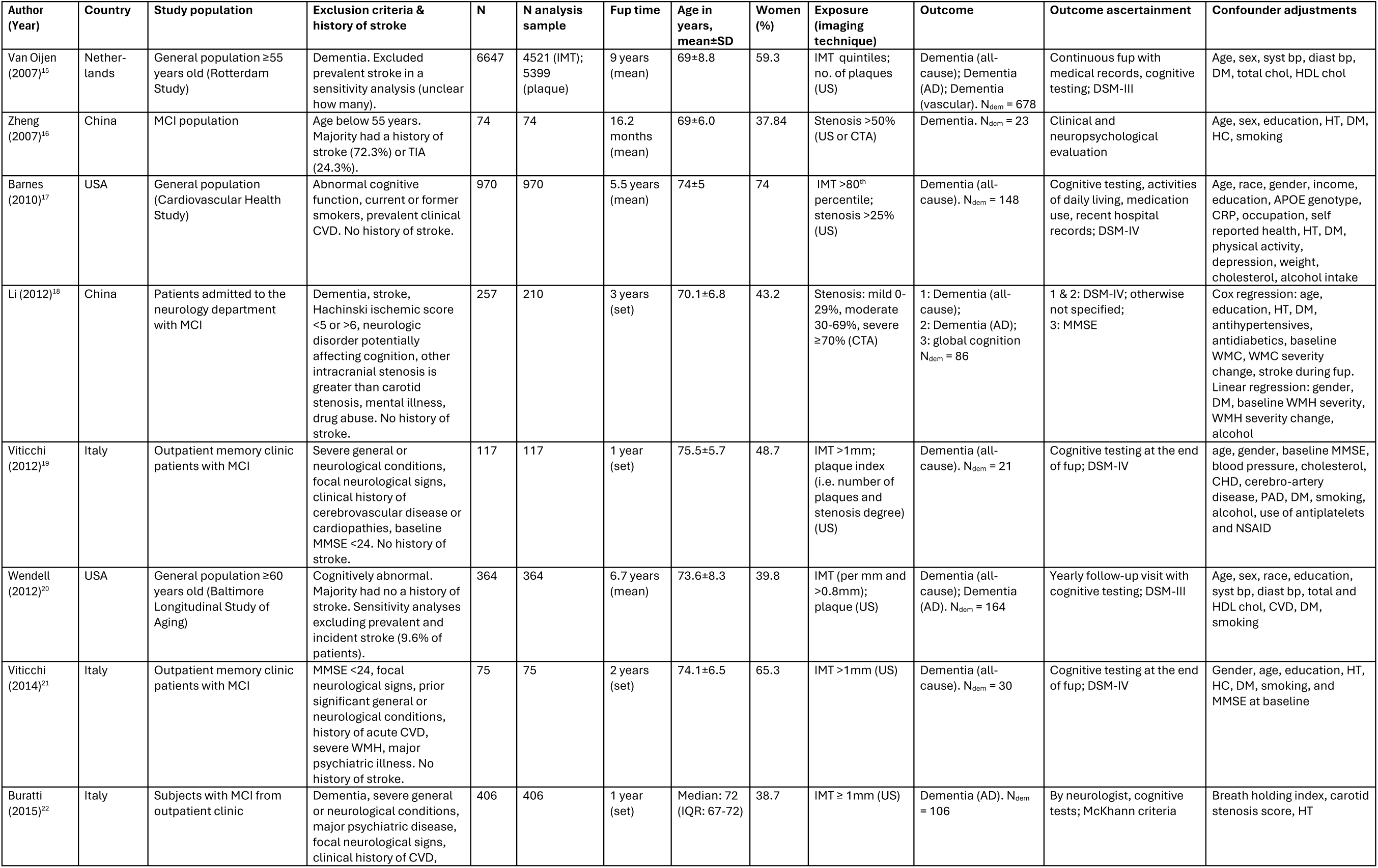
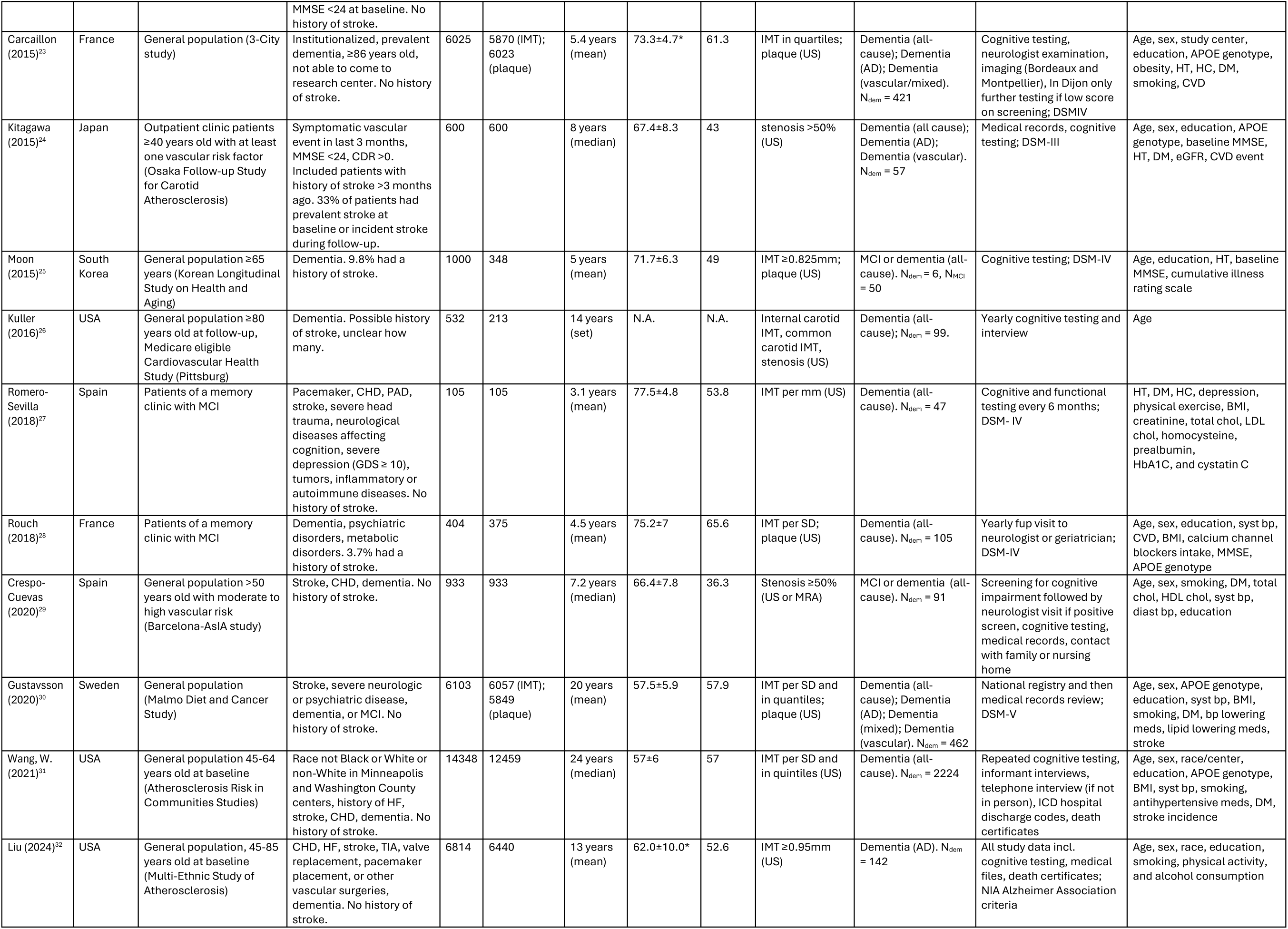
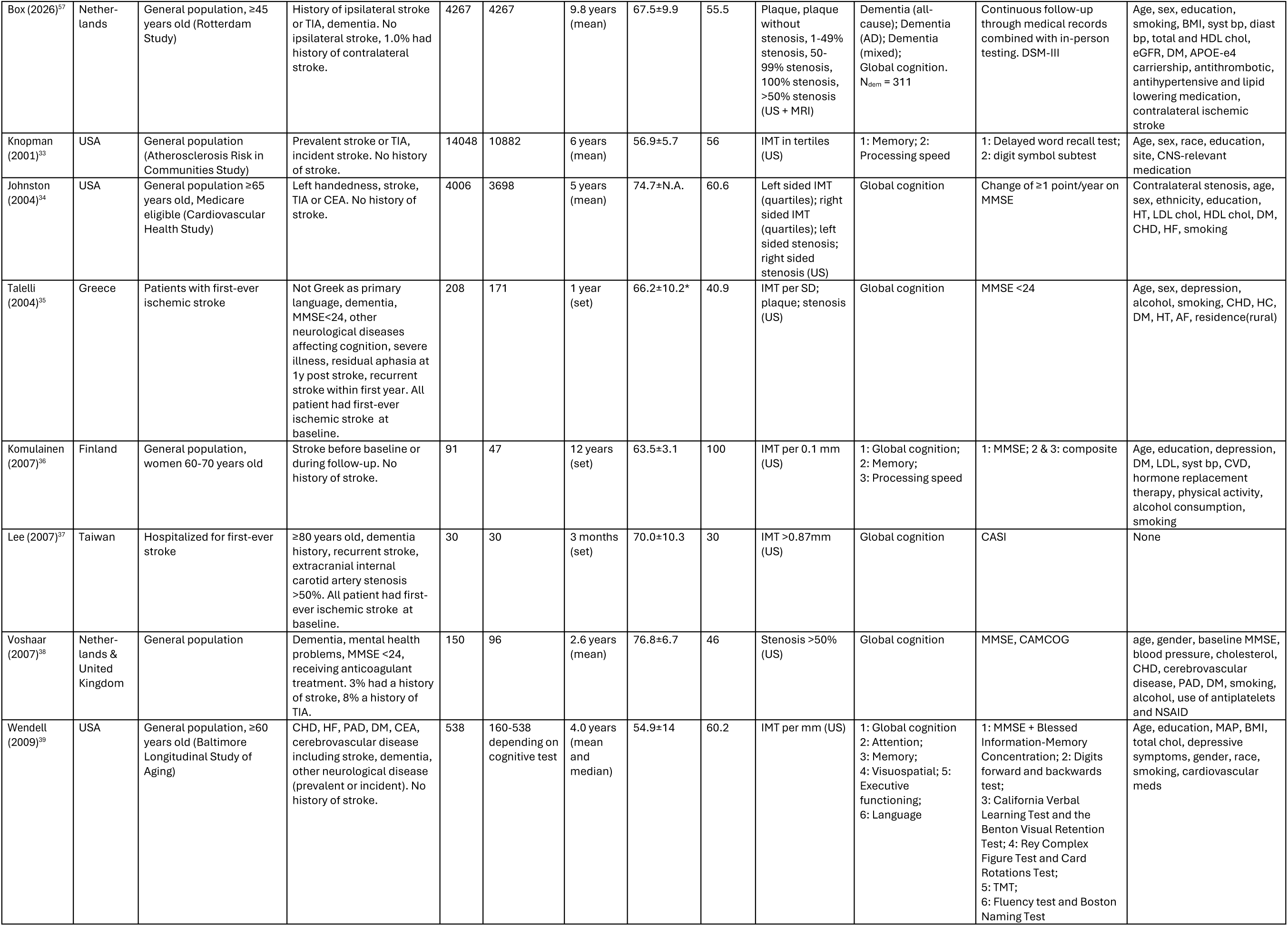
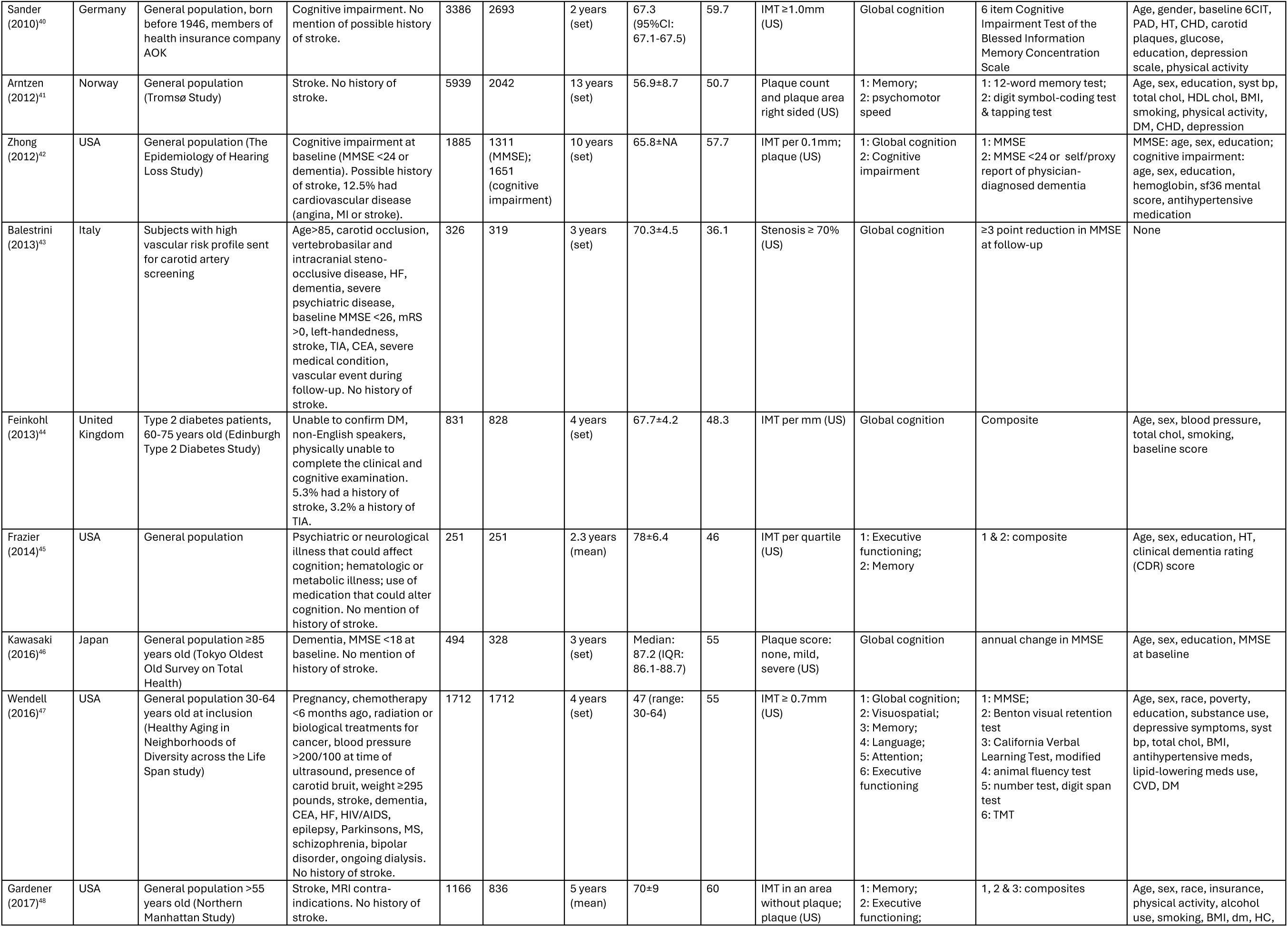
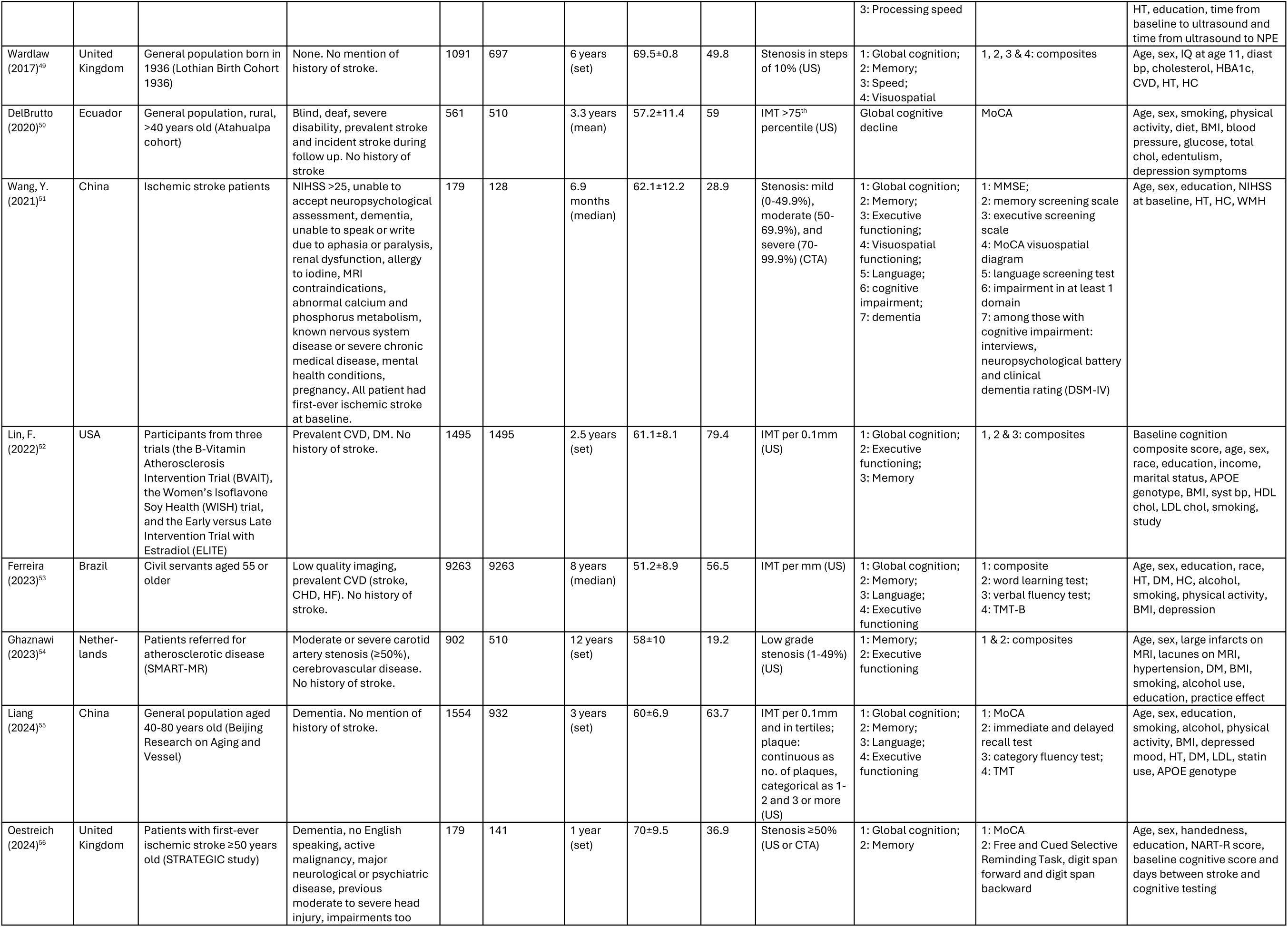
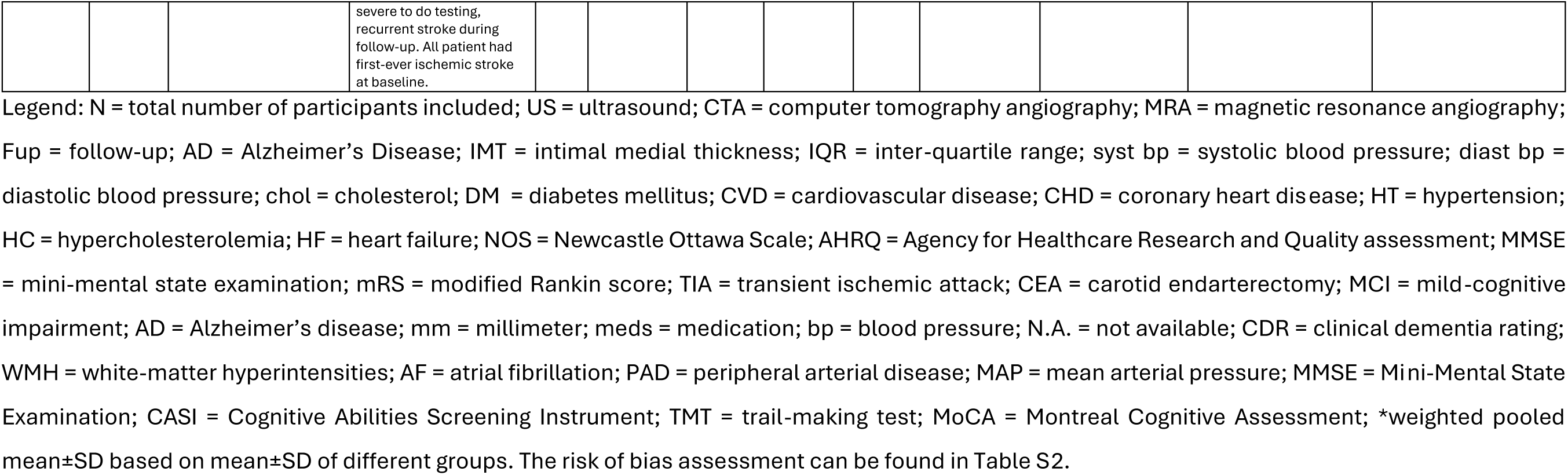
Study characteristics of observational cohort studies.

| **Author (Year)** | **Country** | **Study population** | **Exclusion criteria & history of stroke** | **N** | **N analysis sample** | **Fup time** | **Age in years, mean±SD** | **Women (%)** | **Exposure (imaging technique)** | **Outcome** | **Outcome ascertainment** | **Confounder adjustments** |
|---|---|---|---|---|---|---|---|---|---|---|---|---|
| Van Oijen (2007)<sup>15</sup> | Nether-lands | General population ≥55 years old (Rotterdam Study) | Dementia. Excluded prevalent stroke in a sensitivity analysis (unclear how many). | 6647 | 4521 (IMT); 5399 (plaque) | 9 years (mean) | 69±8.8 | 59.3 | IMT quintiles; no. of plaques (US) | Dementia (all-cause); Dementia (AD); Dementia (vascular). N<sub>dem</sub> = 678 | Continuous fup with medical records, cognitive testing; DSM-III | Age, sex, syst bp, diast bp, DM, total chol, HDL chol |
| Zheng (2007)<sup>16</sup> | China | MCI population | Age below 55 years. Majority had a history of stroke (72.3%) or TIA (24.3%). | 74 | 74 | 16.2 months (mean) | 69±6.0 | 37.84 | Stenosis >50% (US or CTA) | Dementia. N<sub>dem</sub> = 23 | Clinical and neuropsychological evaluation | Age, sex, education, HT, DM, HC, smoking |
| Barnes (2010)<sup>17</sup> | USA | General population (Cardiovascular Health Study) | Abnormal cognitive function, current or former smokers, prevalent clinical CVD. No history of stroke. | 970 | 970 | 5.5 years (mean) | 74±5 | 74 | IMT >80<sup>th</sup> percentile; stenosis >25% (US) | Dementia (all-cause). N<sub>dem</sub> = 148 | Cognitive testing, activities of daily living, medication use, recent hospital records; DSM-IV | Age, race, gender, income, education, APOE genotype, CRP, occupation, self reported health, HT, DM, physical activity, depression, weight, cholesterol, alcohol intake |
| Li (2012)<sup>18</sup> | China | Patients admitted to the neurology department with MCI | Dementia, stroke, Hachinski ischemic score <5 or >6, neurologic disorder potentially affecting cognition, other intracranial stenosis is greater than carotid stenosis, mental illness, drug abuse. No history of stroke. | 257 | 210 | 3 years (set) | 70.1±6.8 | 43.2 | Stenosis: mild 0-29%, moderate 30-69%, severe ≥70% (CTA) | 1: Dementia (all-cause);<br>2: Dementia (AD);<br>3: global cognition<br>N<sub>dem</sub> = 86 | 1 & 2: DSM-IV; otherwise not specified;<br>3: MMSE | Cox regression: age, education, HT, DM, antihypertensives, antidiabetics, baseline WMC, WMC severity change, stroke during fup. Linear regression: gender, DM, baseline WMH severity, WMH severity change, alcohol |
| Viticchi (2012)<sup>19</sup> | Italy | Outpatient memory clinic patients with MCI | Severe general or neurological conditions, focal neurological signs, clinical history of cerebrovascular disease or cardiopathies, baseline MMSE <24. No history of stroke. | 117 | 117 | 1 year (set) | 75.5±5.7 | 48.7 | IMT >1mm; plaque index (i.e. number of plaques and stenosis degree) (US) | Dementia (all-cause). N<sub>dem</sub> = 21 | Cognitive testing at the end of fup; DSM-IV | age, gender, baseline MMSE, blood pressure, cholesterol, CHD, cerebro-artery disease, PAD, DM, smoking, alcohol, use of antiplatelets and NSAID |
| Wendell (2012)<sup>20</sup> | USA | General population ≥60 years old (Baltimore Longitudinal Study of Aging) | Cognitively abnormal. Majority had no a history of stroke. Sensitivity analyses excluding prevalent and incident stroke (9.6% of patients). | 364 | 364 | 6.7 years (mean) | 73.6±8.3 | 39.8 | IMT (per mm and >0.8mm); plaque (US) | Dementia (all-cause); Dementia (AD). N<sub>dem</sub> = 164 | Yearly follow-up visit with cognitive testing; DSM-III | Age, sex, race, education, syst bp, diast bp, total and HDL chol, CVD, DM, smoking |
| Viticchi (2014)<sup>21</sup> | Italy | Outpatient memory clinic patients with MCI | MMSE <24, focal neurological signs, prior significant general or neurological conditions, history of acute CVD, severe WMH, major psychiatric illness. No history of stroke. | 75 | 75 | 2 years (set) | 74.1±6.5 | 65.3 | IMT >1mm (US) | Dementia (all-cause). N<sub>dem</sub> = 30 | Cognitive testing at the end of fup; DSM-IV | Gender, age, education, HT, HC, DM, smoking, and MMSE at baseline |
| Buratti (2015)<sup>22</sup> | Italy | Subjects with MCI from outpatient clinic | Dementia, severe general or neurological conditions, major psychiatric disease, focal neurological signs, clinical history of CVD, MMSE <24 at baseline. No history of stroke. | 406 | 406 | 1 year (set) | Median: 72 (IQR: 67-72) | 38.7 | IMT ≥ 1mm (US) | Dementia (AD). N<sub>dem</sub> = 106 | By neurologist, cognitive tests; McKhann criteria | Breath holding index, carotid stenosis score, HT |
| Carcaillon (2015)<sup>23</sup> | France | General population (3-City study) | Institutionalized, prevalent dementia, ≥86 years old, not able to come to research center. No history of stroke. | 6025 | 5870 (IMT); 6023 (plaque) | 5.4 years (mean) | 73.3±4.7* | 61.3 | IMT in quartiles; plaque (US) | Dementia (all-cause); Dementia (AD); Dementia (vascular/mixed). N<sub>dem</sub> = 421 | Cognitive testing, neurologist examination, imaging (Bordeaux and Montpellier), In Dijon only further testing if low score on screening; DSMIV | Age, sex, study center, education, APOE genotype, obesity, HT, HC, DM, smoking, CVD |
| Kitagawa (2015)<sup>24</sup> | Japan | Outpatient clinic patients ≥40 years old with at least one vascular risk factor (Osaka Follow-up Study for Carotid Atherosclerosis) | Symptomatic vascular event in last 3 months, MMSE <24, CDR >0. Included patients with history of stroke >3 months ago. 33% of patients had prevalent stroke at baseline or incident stroke during follow-up. | 600 | 600 | 8 years (median) | 67.4±8.3 | 43 | stenosis >50% (US) | Dementia (all cause); Dementia (AD); Dementia (vascular). N<sub>dem</sub> = 57 | Medical records, cognitive testing; DSM-III | Age, sex, education, APOE genotype, baseline MMSE, HT, DM, eGFR, CVD event |
| Moon (2015)<sup>25</sup> | South Korea | General population ≥65 years (Korean Longitudinal Study on Health and Aging) | Dementia. 9.8% had a history of stroke. | 1000 | 348 | 5 years (mean) | 71.7±6.3 | 49 | IMT ≥0.825mm; plaque (US) | MCI or dementia (all-cause). N<sub>dem</sub> = 6, N<sub>MCI</sub> = 50 | Cognitive testing; DSM-IV | Age, education, HT, baseline MMSE, cumulative illness rating scale |
| Kuller (2016)<sup>26</sup> | USA | General population ≥80 years old at follow-up, Medicare eligible Cardiovascular Health Study (Pittsburg) | Dementia. Possible history of stroke, unclear how many. | 532 | 213 | 14 years (set) | N.A. | N.A. | Internal carotid IMT, common carotid IMT, stenosis (US) | Dementia (all-cause); N<sub>dem</sub> = 99. | Yearly cognitive testing and interview | Age |
| Romero-Sevilla (2018)<sup>27</sup> | Spain | Patients of a memory clinic with MCI | Pacemaker, CHD, PAD, stroke, severe head trauma, neurological diseases affecting cognition, severe depression (GDS ≥ 10), tumors, inflammatory or autoimmune diseases. No history of stroke. | 105 | 105 | 3.1 years (mean) | 77.5±4.8 | 53.8 | IMT per mm (US) | Dementia (all-cause). N<sub>dem</sub> = 47 | Cognitive and functional testing every 6 months; DSM- IV | HT, DM, HC, depression, physical exercise, BMI, creatinine, total chol, LDL chol, homocysteine, prealbumin,<br>HbA1C, and cystatin C |
| Rouch (2018)<sup>28</sup> | France | Patients of a memory clinic with MCI | Dementia, psychiatric disorders, metabolic disorders. 3.7% had a history of stroke. | 404 | 375 | 4.5 years (mean) | 75.2±7 | 65.6 | IMT per SD; plaque (US) | Dementia (all-cause). N<sub>dem</sub> = 105 | Yearly fup visit to neurologist or geriatrician; DSM-IV | Age, sex, education, syst bp, CVD, BMI, calcium channel blockers intake, MMSE, APOE genotype |
| Crespo-Cuevas (2020)<sup>29</sup> | Spain | General population >50 years old with moderate to high vascular risk (Barcelona-AsIA study) | Stroke, CHD, dementia. No history of stroke. | 933 | 933 | 7.2 years (median) | 66.4±7.8 | 36.3 | Stenosis ≥50% (US or MRA) | MCI or dementia (all-cause). N<sub>dem</sub> = 91 | Screening for cognitive impairment followed by neurologist visit if positive screen, cognitive testing, medical records, contact with family or nursing home | Age, sex, smoking, DM, total chol, HDL chol, syst bp, diast bp, education |
| Gustavsson (2020)<sup>30</sup> | Sweden | General population (Malmo Diet and Cancer Study) | Stroke, severe neurologic or psychiatric disease, dementia, or MCI. No history of stroke. | 6103 | 6057 (IMT); 5849 (plaque) | 20 years (mean) | 57.5±5.9 | 57.9 | IMT per SD and in quantiles; plaque (US) | Dementia (all-cause); Dementia (AD); Dementia (mixed); Dementia (vascular). N<sub>dem</sub> = 462 | National registry and then medical records review; DSM-V | Age, sex, APOE genotype, education, syst bp, BMI, smoking, DM, bp lowering meds, lipid lowering meds, stroke |
| Wang, W. (2021)<sup>31</sup> | USA | General population 45-64 years old at baseline (Atherosclerosis Risk in Communities Studies) | Race not Black or White or non-White in Minneapolis and Washington County centers, history of HF, stroke, CHD, dementia. No history of stroke. | 14348 | 12459 | 24 years (median) | 57±6 | 57 | IMT per SD and in quintiles (US) | Dementia (all-cause). N<sub>dem</sub> = 2224 | Repeated cognitive testing, informant interviews, telephone interview (if not in person), ICD hospital discharge codes, death certificates | Age, sex, race/center, education, APOE genotype, BMI, syst bp, smoking, antihypertensive meds, DM, stroke incidence |
| Liu (2024)<sup>32</sup> | USA | General population, 45-85 years old at baseline (Multi-Ethnic Study of Atherosclerosis) | CHD, HF, stroke, TIA, valve replacement, pacemaker placement, or other vascular surgeries, dementia. No history of stroke. | 6814 | 6440 | 13 years (mean) | 62.0±10.0* | 52.6 | IMT ≥0.95mm (US) | Dementia (AD). N<sub>dem</sub> = 142 | All study data incl. cognitive testing, medical files, death certificates; NIA Alzheimer Association criteria | Age, sex, race, education, smoking, physical activity, and alcohol consumption |
| Box (2026)<sup>57</sup> | Nether-lands | General population, ≥45 years old (Rotterdam Study) | History of ipsilateral stroke or TIA, dementia. No ipsilateral stroke, 1.0% had history of contralateral stroke. | 4267 | 4267 | 9.8 years (mean) | 67.5±9.9 | 55.5 | Plaque, plaque without stenosis, 1-49% stenosis, 50-99% stenosis, 100% stenosis, >50% stenosis (US + MRI) | Dementia (all-cause); Dementia (AD); Dementia (mixed);<br>Global cognition. N<sub>dem</sub> = 311 | Continuous follow-up through medical records combined with in-person testing. DSM-III | Age, sex, education, smoking, BMI, syst bp, diast bp, total and HDL chol, eGFR, DM, APOE-e4 carriership, antithrombotic, antihypertensive and lipid lowering medication, contralateral ischemic stroke |
| Knopman (2001)<sup>33</sup> | USA | General population (Atherosclerosis Risk in Communities Study) | Prevalent stroke or TIA, incident stroke. No history of stroke. | 14048 | 10882 | 6 years (mean) | 56.9±5.7 | 56 | IMT in tertiles (US) | 1: Memory; 2: Processing speed | 1: Delayed word recall test;<br>2: digit symbol subtest | Age, sex, race, education, site, CNS-relevant medication |
| Johnston (2004)<sup>34</sup> | USA | General population ≥65 years old, Medicare eligible (Cardiovascular Health Study) | Left handedness, stroke, TIA or CEA. No history of stroke. | 4006 | 3698 | 5 years (mean) | 74.7±N.A. | 60.6 | Left sided IMT (quartiles); right sided IMT (quartiles); left sided stenosis; right sided stenosis (US) | Global cognition | Change of ≥1 point/year on MMSE | Contralateral stenosis, age, sex, ethnicity, education, HT, LDL chol, HDL chol, DM, CHD, HF, smoking |
| Talelli (2004)<sup>35</sup> | Greece | Patients with first-ever ischemic stroke | Not Greek as primary language, dementia, MMSE<24, other neurological diseases affecting cognition, severe illness, residual aphasia at 1y post stroke, recurrent stroke within first year. All patient had first-ever ischemic stroke at baseline. | 208 | 171 | 1 year (set) | 66.2±10.2* | 40.9 | IMT per SD; plaque; stenosis (US) | Global cognition | MMSE <24 | Age, sex, depression, alcohol, smoking, CHD, HC, DM, HT, AF, residence(rural) |
| Komulainen (2007)<sup>36</sup> | Finland | General population, women 60-70 years old | Stroke before baseline or during follow-up. No history of stroke. | 91 | 47 | 12 years (set) | 63.5±3.1 | 100 | IMT per 0.1 mm (US) | 1: Global cognition;<br>2: Memory;<br>3: Processing speed | 1: MMSE; 2 & 3: composite | Age, education, depression, DM, LDL, syst bp, CVD, hormone replacement therapy, physical activity, alcohol consumption, smoking |
| Lee (2007)<sup>37</sup> | Taiwan | Hospitalized for first-ever stroke | ≥80 years old, dementia history, recurrent stroke, extracranial internal carotid artery stenosis >50%. All patient had first-ever ischemic stroke at baseline. | 30 | 30 | 3 months (set) | 70.0±10.3 | 30 | IMT >0.87mm (US) | Global cognition | CASI | None |
| Voshaar (2007)<sup>38</sup> | Nether-lands & United Kingdom | General population | Dementia, mental health problems, MMSE <24, receiving anticoagulant treatment. 3% had a history of stroke, 8% a history of TIA. | 150 | 96 | 2.6 years (mean) | 76.8±6.7 | 46 | Stenosis >50% (US) | Global cognition | MMSE, CAMCOG | age, gender, baseline MMSE, blood pressure, cholesterol, CHD, cerebrovascular disease, PAD, DM, smoking, alcohol, use of antiplatelets and NSAID |
| Wendell (2009)<sup>39</sup> | USA | General population, ≥60 years old (Baltimore Longitudinal Study of Aging) | CHD, HF, PAD, DM, CEA, cerebrovascular disease including stroke, dementia, other neurological disease (prevalent or incident). No history of stroke. | 538 | 160-538 depending on cognitive test | 4.0 years (mean and median) | 54.9±14 | 60.2 | IMT per mm (US) | 1: Global cognition<br>2: Attention;<br>3: Memory;<br>4: Visuospatial; 5: Executive functioning;<br>6: Language | 1: MMSE + Blessed Information-Memory Concentration; 2: Digits forward and backwards test;<br>3: California Verbal Learning Test and the Benton Visual Retention Test; 4: Rey Complex Figure Test and Card Rotations Test;<br>5: TMT;<br>6: Fluency test and Boston Naming Test | Age, education, MAP, BMI, total chol, depressive symptoms, gender, race, smoking, cardiovascular meds |
| Sander (2010)<sup>40</sup> | Germany | General population, born before 1946, members of health insurance company AOK | Cognitive impairment. No mention of possible history of stroke. | 3386 | 2693 | 2 years (set) | 67.3 (95%CI: 67.1-67.5) | 59.7 | IMT ≥1.0mm (US) | Global cognition | 6 item Cognitive Impairment Test of the Blessed Information Memory Concentration Scale | Age, gender, baseline 6CIT, PAD, HT, CHD, carotid plaques, glucose, education, depression scale, physical activity |
| Arntzen (2012)<sup>41</sup> | Norway | General population (Tromsø Study) | Stroke. No history of stroke. | 5939 | 2042 | 13 years (set) | 56.9±8.7 | 50.7 | Plaque count and plaque area right sided (US) | 1: Memory;<br>2: psychomotor speed | 1: 12-word memory test;<br>2: digit symbol-coding test & tapping test | Age, sex, education, syst bp, total chol, HDL chol, BMI, smoking, physical activity, DM, CHD, depression |
| Zhong (2012)<sup>42</sup> | USA | General population (The Epidemiology of Hearing Loss Study) | Cognitive impairment at baseline (MMSE <24 or dementia). Possible history of stroke, 12.5% had cardiovascular disease (angina, MI or stroke). | 1885 | 1311 (MMSE); 1651 (cognitive impairment) | 10 years (set) | 65.8±NA | 57.7 | IMT per 0.1mm; plaque (US) | 1: Global cognition<br>2: Cognitive impairment | 1: MMSE<br>2: MMSE <24 or self/proxy report of physician-diagnosed dementia | MMSE: age, sex, education; cognitive impairment: age, sex, education, hemoglobin, sf36 mental score, antihypertensive medication |
| Balestrini (2013)<sup>43</sup> | Italy | Subjects with high vascular risk profile sent for carotid artery screening | Age>85, carotid occlusion, vertebrobasilar and intracranial steno-occlusive disease, HF, dementia, severe psychiatric disease, baseline MMSE <26, mRS >0, left-handedness, stroke, TIA, CEA, severe medical condition, vascular event during follow-up. No history of stroke. | 326 | 319 | 3 years (set) | 70.3±4.5 | 36.1 | Stenosis ≥ 70% (US) | Global cognition | ≥3 point reduction in MMSE at follow-up | None |
| Feinkohl (2013)<sup>44</sup> | United Kingdom | Type 2 diabetes patients, 60-75 years old (Edinburgh Type 2 Diabetes Study) | Unable to confirm DM, non-English speakers, physically unable to complete the clinical and cognitive examination. 5.3% had a history of stroke, 3.2% a history of TIA. | 831 | 828 | 4 years (set) | 67.7±4.2 | 48.3 | IMT per mm (US) | Global cognition | Composite | Age, sex, blood pressure, total chol, smoking, baseline score |
| Frazier (2014)<sup>45</sup> | USA | General population | Psychiatric or neurological illness that could affect cognition; hematologic or metabolic illness; use of medication that could alter cognition. No mention of history of stroke. | 251 | 251 | 2.3 years (mean) | 78±6.4 | 46 | IMT per quartile (US) | 1: Executive functioning;<br>2: Memory | 1 & 2: composite | Age, sex, education, HT, clinical dementia rating (CDR) score |
| Kawasaki (2016)<sup>46</sup> | Japan | General population ≥85 years old (Tokyo Oldest Old Survey on Total Health) | Dementia, MMSE <18 at baseline. No mention of history of stroke. | 494 | 328 | 3 years (set) | Median: 87.2 (IQR: 86.1-88.7) | 55 | Plaque score: none, mild, severe (US) | Global cognition | annual change in MMSE | Age, sex, education, MMSE at baseline |
| Wendell (2016)<sup>47</sup> | USA | General population 30-64 years old at inclusion (Healthy Aging in Neighborhoods of Diversity across the Life Span study) | Pregnancy, chemotherapy <6 months ago, radiation or biological treatments for cancer, blood pressure >200/100 at time of ultrasound, presence of carotid bruit, weight ≥295 pounds, stroke, dementia, CEA, HF, HIV/AIDS, epilepsy, Parkinsons, MS, schizophrenia, bipolar disorder, ongoing dialysis. No history of stroke. | 1712 | 1712 | 4 years (set) | 47 (range: 30-64) | 55 | IMT ≥ 0.7mm (US) | 1: Global cognition;<br>2: Visuospatial;<br>3: Memory;<br>4: Language;<br>5: Attention;<br>6: Executive functioning | 1: MMSE;<br>2: Benton visual retention test<br>3: California Verbal Learning Test, modified<br>4: animal fluency test<br>5: number test, digit span test<br>6: TMT | Age, sex, race, poverty, education, substance use, depressive symptoms, syst bp, total chol, BMI, antihypertensive meds, lipid-lowering meds use, CVD, DM |
| Gardener (2017)<sup>48</sup> | USA | General population >55 years old (Northern Manhattan Study) | Stroke, MRI contra-indications. No history of stroke. | 1166 | 836 | 5 years (mean) | 70±9 | 60 | IMT in an area without plaque; plaque (US) | 1: Memory;<br>2: Executive functioning;<br>3: Processing speed | 1, 2 & 3: composites | Age, sex, race, insurance, physical activity, alcohol use, smoking, BMI, dm, HC, HT, education, time from baseline to ultrasound and time from ultrasound to NPE |
| Wardlaw (2017)<sup>49</sup> | United Kingdom | General population born in 1936 (Lothian Birth Cohort 1936) | None. No mention of history of stroke. | 1091 | 697 | 6 years (set) | 69.5±0.8 | 49.8 | Stenosis in steps of 10% (US) | 1: Global cognition;<br>2: Memory;<br>3: Speed;<br>4: Visuospatial | 1, 2, 3 & 4: composites | Age, sex, IQ at age 11, diast bp, cholesterol, HBA1c, CVD, HT, HC |
| DelBrutto (2020)<sup>50</sup> | Ecuador | General population, rural, >40 years old (Atahualpa cohort) | Blind, deaf, severe disability, prevalent stroke and incident stroke during follow up. No history of stroke | 561 | 510 | 3.3 years (mean) | 57.2±11.4 | 59 | IMT >75<sup>th</sup> percentile (US) | Global cognitive decline | MoCA | Age, sex, smoking, physical activity, diet, BMI, blood pressure, glucose, total chol, edentulism, depression symptoms |
| Wang, Y. (2021)<sup>51</sup> | China | Ischemic stroke patients | NIHSS >25, unable to accept neuropsychological assessment, dementia, unable to speak or write due to aphasia or paralysis, renal dysfunction, allergy to iodine, MRI contraindications, abnormal calcium and phosphorus metabolism, known nervous system disease or severe chronic medical disease, mental health conditions, pregnancy. All patient had first-ever ischemic stroke at baseline. | 179 | 128 | 6.9 months (median) | 62.1±12.2 | 28.9 | Stenosis: mild (0-49.9%), moderate (50-69.9%), and severe (70-99.9%) (CTA) | 1: Global cognition;<br>2: Memory;<br>3: Executive functioning;<br>4: Visuospatial functioning;<br>5: Language;<br>6: cognitive impairment;<br>7: dementia | 1: MMSE;<br>2: memory screening scale<br>3: executive screening scale<br>4: MoCA visuospatial diagram<br>5: language screening test<br>6: impairment in at least 1 domain<br>7: among those with cognitive impairment: interviews, neuropsychological battery and clinical dementia rating (DSM-IV) | Age, sex, education, NIHSS at baseline, HT, HC, WMH |
| Lin, F. (2022)<sup>52</sup> | USA | Participants from three trials (the B-Vitamin Atherosclerosis Intervention Trial (BVAIT), the Women’s Isoflavone Soy Health (WISH) trial, and the Early versus Late Intervention Trial with Estradiol (ELITE) | Prevalent CVD, DM. No history of stroke. | 1495 | 1495 | 2.5 years (set) | 61.1±8.1 | 79.4 | IMT per 0.1mm (US) | 1: Global cognition;<br>2: Executive functioning;<br>3: Memory | 1, 2 & 3: composites | Baseline cognition composite score, age, sex, race, education, income, marital status, APOE genotype, BMI, syst bp, HDL chol, LDL chol, smoking, study |
| Ferreira (2023)<sup>53</sup> | Brazil | Civil servants aged 55 or older | Low quality imaging, prevalent CVD (stroke, CHD, HF). No history of stroke. | 9263 | 9263 | 8 years (median) | 51.2±8.9 | 56.5 | IMT per mm (US) | 1: Global cognition;<br>2: Memory;<br>3: Language;<br>4: Executive functioning | 1: composite<br>2: word learning test;<br>3: verbal fluency test;<br>4: TMT-B | Age, sex, education, race, HT, DM, HC, alcohol, smoking, physical activity, BMI, depression |
| Ghaznawi (2023)<sup>54</sup> | Nether-lands | Patients referred for atherosclerotic disease (SMART-MR) | Moderate or severe carotid artery stenosis (≥50%), cerebrovascular disease. No history of stroke. | 902 | 510 | 12 years (set) | 58±10 | 19.2 | Low grade stenosis (1-49%) (US) | 1: Memory;<br>2: Executive functioning | 1 & 2: composites | Age, sex, large infarcts on MRI, lacunes on MRI, hypertension, DM, BMI, smoking, alcohol use, education, practice effect |
| Liang (2024)<sup>55</sup> | China | General population aged 40-80 years old (Beijing Research on Aging and Vessel) | Dementia. No mention of history of stroke. | 1554 | 932 | 3 years (set) | 60±6.9 | 63.7 | IMT per 0.1mm and in tertiles; plaque: continuous as no. of plaques, categorical as 1-2 and 3 or more (US) | 1: Global cognition;<br>2: Memory;<br>3: Language;<br>4: Executive functioning | 1: MoCA<br>2: immediate and delayed recall test<br>3: category fluency test;<br>4: TMT | Age, sex, education, smoking, alcohol, physical activity, BMI, depressed mood, HT, DM, LDL, statin use, APOE genotype |
| Oestreich (2024)<sup>56</sup> | United Kingdom | Patients with first-ever ischemic stroke ≥50 years old (STRATEGIC study) | Dementia, no English speaking, active malignancy, major neurological or psychiatric disease, previous moderate to severe head injury, impairments too severe to do testing, recurrent stroke during follow-up. All patient had first-ever ischemic stroke at baseline. | 179 | 141 | 1 year (set) | 70±9.5 | 36.9 | Stenosis ≥50% (US or CTA) | 1: Global cognition;<br>2: Memory | 1: MoCA<br>2: Free and Cued Selective Reminding Task, digit span forward and digit span backward | Age, sex, handedness, education, NART-R score, baseline cognitive score and days between stroke and cognitive testing |

Legend: N = total number of participants included; US = ultrasound; CTA = computer tomography angiography; MRA = magnetic resonance angiography; Fup = follow-up; AD = Alzheimer’s Disease; IMT = intimal medial thickness; IQR = inter-quartile range; syst bp = systolic blood pressure; diast bp = diastolic blood pressure; chol = cholesterol; DM  = diabetes mellitus; CVD = cardiovascular disease; CHD = coronary heart disease; HT = hypertension; HC = hypercholesterolemia; HF = heart failure; NOS = Newcastle Ottawa Scale; AHRQ = Agency for Healthcare Research and Quality assessment; MMSE = mini-mental state examination; mRS = modified Rankin score; TIA = transient ischemic attack; CEA = carotid endarterectomy; MCI = mild-cognitive impairment; AD = Alzheimer’s disease; mm = millimeter; meds = medication; bp = blood pressure; N.A. = not available; CDR = clinical dementia rating; WMH = white-matter hyperintensities; AF = atrial fibrillation; PAD = peripheral arterial disease; MAP = mean arterial pressure; MMSE = Mini-Mental State Examination; CASI = Cognitive Abilities Screening Instrument; TMT = trail-making test; MoCA = Montreal Cognitive Assessment; \*weighted pooled mean±SD based on mean±SD of different groups. The risk of bias assessment can be found in Table S2.

**Table 2:** Study characteristics and findings of randomized controlled trials.

| **Author (Year)** | **Trial name** | **Journal** | **Country** | **Blinding** | **Symptoms (INT group)** | **Stenosis criterion** | **Intervention type** | **Reference group** | **Follow up duration** | **n (total)** | **n (total attrition)** | **n (INT)** | **n (REF)** | **Mean age (total)** | **% women (total)** | **Cognitive domains** | **Main findings** |
|---|---|---|---|---|---|---|---|---|---|---|---|---|---|---|---|---|---|
| Pettigrew (2000)<sup>58</sup> | ACAS | Neurology | USA | no | Asymptomatic | ≥60% stenosis | CEA | patients with medical management | 1 month + 3 months + every 6 months until 5 years | 1662 | 3 | 828 | 834 | 67 (40-79) | 34.0 | Global cognition (MMSE) | No significant difference in MMSE change between surgical and medical groups over 5 years. |
| Marshall (2014)<sup>59</sup> | RECON | Neurology | USA | single | Symptomatic | occlusion | EC-IC surgery | patients with medical management | 24 months | 41 | 12 | 19 | 22 | 57±9.3 | 20.7 | Global cognition (composite cognitive z-score*) | No significant difference in 2-year composite cognitive z-score change between surgical and medical groups. (Effect estimate 0.02, 95% CI −0.50 to 0.54; p=0.93). |
| Shi, H. (2016)<sup>60</sup> | / | Chin J Cerebrovasc Dis | China | no | Asymptomatic | ≥70% ipsilateral stenosis, with contralateral stenosis <30% | CAS | patients with medical management | 12 months | 200 | 3 | 100 | 100 | 66±7 | 29.5 | Global cognition (MoCA) | MoCA score improved more after CAS than medical management (SMD = 1.02 [0.73-1.31], P < 0.01) |
| Gao (2022)<sup>61</sup> | / | Medicine | China | single | Symptomatic | >70% stenosis | CAS | patients with medical management | 1 month +3 months +6 months | 90 | 0 | 45 | 45 | 64.3±8.1 | 41.1 | Global cognition (MMSE, MoCA)<br>Attention<br>Language<br>Memory<br>Visuospatial/executive functioning | CAS showed greater 6-month improvement than medical management in cognition (standardized effect: 0.86 [0.57-1.16]), with significant benefits in the MoCA attention and language subscale scores. |
| Halliday (2022)<sup>62</sup> | ACST-1 | Eur J Vasc Endovasc Surg | United Kingdom and Sweden | single | Asymptomatic | 60–99% stenosis | CEA | Patients with delayed CEA | 19.4 years (median) | 1601 | 1 | 796 | 804 | 69±7 | 36.1 | Dementia | No evidence that CEA reduced dementia risk compared with deferred CEA. (Effect estimate: HR 0.98, 95% CI 0.75–1.28; p=0.89). |
| Lazar (2026)<sup>63</sup> | CREST-2 | JAMA | USA | Single | Asymptomatic | ≥70% stenosis | CEA + CAS | patients with intensive medical management | 2.24 years (mean) | 2366 | 201 | 1072 | 1093 | 70.1±7.7 | 37.8 | Memory, executive function, attention, processing speed | No significant difference in composite cognition between revascularization plus IMM and IMM alone at 4 years in either the CAS trial (adjusted difference 0.03, 95% CI −0.09 to 0.15) or the CEA trial (0.02, 95% CI −0.09 to 0.14). |

Legend: SD = standard deviation; CEA = carotid endarterectomy; EC-IC surgery = Extracranial-intracranial surgical bypass; CAS = carotid artery stenting; INT = intervention; REF = reference; all = the combined intervention and reference groups; ACAS: Asymptomatic Carotid Atherosclerosis Study; RECON: Randomized Evaluation of Carotid Occlusion and Neurocognition; ACST-1: Asymptomatic Carotid Surgery Trial; IMM: intensive medical management.
\*Composite neurocognitive z score: derived from 14 standardized neuropsychological tests assessing global, left-hemisphere, and right-hemisphere functions.

### Non-randomized intervention studies

Across all 48 non-randomized intervention studies,^65–112^ 22 evaluated carotid endarterectomy (CEA),^65–72,74,75,78,79,82,87,89,94,96,99,102,108,111,112^ 16 evaluated carotid artery stenting (CAS),^73,76,80,81,83–86,91–93,97,101,106,107,110^ and 10 included both procedures (Table 3).^77,88,90,95,98,100,103–105,109^ Study sample size ranged from 19 to 2,098 participants (median 68), and follow-up ranged from 1 to 96 months (median 6 months). Thirty-one studies reported no loss to follow-up;^65,67,68,70,75,77–83,85–87,89–93,95,98–101,103,104,106,108,109,111^ among the remaining 17, attrition ranged from 0.6% to 63.5% (median 9.1%).^66,69,71–74,76,84,88,94,96,97,102,105,107,110,112^ Symptom status was reported in 47 studies, 17 studies included symptomatic patients,^65–68,71,77,83,85,92,93,97,101,104,106,109,110^ 10 included asymptomatic patients,^70,74,75,80,86,94,103,105,107,111^ and 20 included both, with asymptomatic patients comprising 31– 81% of mixed populations (median 52%).^69,72,73,76,78,79,81,82,84,87,88,90,91,96,100–102,108,112^ The characteristics of the reference groups also varied substantially. In 25/48 studies, controls met the same eligibility criteria as the intervention group but did not undergo carotid revascularization (17 by patient preference, 2 following clinician-determined treatment decisions, 1 for mixed reasons including health status, and 5 for unknown reasons),^73,76,81,83–87,89–92,95,97–104,106,107,109,110^ 8/48 studies included other surgical patients,^65,67,71,72,82,93,105^ 7/48 used patients with other diseases,^66,70,74,80,88,96,111^ and 8/48 used healthy volunteers as controls.^68,75,77–79,94,108,112^

**Table 3:** Study characteristics of non-randomized intervention studies.

| **Author (Year)** | **Journal** | **Country** | **Randomization** | **Blinding** | **Symptoms (INT group)** | **Intervention type** | **Reference group** | **Follow up duration** | **n (total)** | **n (total attrition)** | **n (INT)** | **INT: Mean age ± SD** | **INT: female%** | **n (REF)** | **REF: Mean age ± SD** | **REF: female%** | **Cognitive domains** |
|---|---|---|---|---|---|---|---|---|---|---|---|---|---|---|---|---|---|
| King (1977)<sup>65</sup> | Journal of Clinical Psychology | USA | no | no | symptomatic | CEA | Underwent major non-neurological surgery | 6 weeks | 19 | 0 | 11 | 65.3±5.7 | 72.7 | 8 | 66.9±9.3 | 87.5 | Language<br>Visual spatial<br>Processing speed<br>Executive functioning |
| Matarazzo (1979)<sup>66</sup> | Journal of Clinical Neuropsychology | USA | no | no | symptomatic | CEA | Patients with diffuse cerebrovascular disease | 20 weeks | 33 | 2 | 17 | 62<sup>a</sup> | 11.8 | 16 | 60<sup>a</sup> | NA | Global cognition<br>Memory<br>Attention<br>Language<br>Processing speed<br>Executive functioning<br>Visual spatial<br>Motor speed |
| Van den Burg (1985)<sup>67</sup> | Psychological Medicine | Netherlands | no | no | symptomatic | CEA | Patients with peripheral vascular surgery | 11 weeks | 40 | 0 | 20 | 59.4<sup>a</sup> | 35 | 20 | 56.8<sup>a</sup> | 10 | Global cognition<br>Memory<br>Attention<br>Language<br>Processing speed<br>Executive functioning<br>Motor speed |
| Hemmingsen (1986)<sup>68</sup> | Ann Neurol | Denmark | no | no | symptomatic | CEA | Patients without symptoms of cerebrovascular or other brain disease | 3 months | 42 | 0 | 31 | 56.8±8.3 | 45.2 | 11 | 58.5±7.0 | 18.2 | Memory<br>Attention<br>Processing speed<br>Executive functioning |
| Heyer (2002)<sup>69</sup> | Arch Neurol | USA | no | no | 52% asymptomatic | CEA | Patients following lumbar spine surgery | 1 month | 108 | 43 | 83 | 70.5±9.2 | 29 | 25 | 73.4±7.6 | 24 | Global cognition<br>Language<br>Visual spatial<br>Processing speed<br>Executive functioning |
| Aharon-Peretz (2003)<sup>70</sup> | European Journal of Neurology | Israel | no | NA | asymptomatic | CEA | Patients with vascular risk factors * | 1 month | 36 | 0 | 22 | 68.6±6.9 | 45.5 | 14 | 63.2±7.4 | 57.1 | Global cognition<br>Executive functioning<br>Attention<br>Memory<br>Visual spatial |
| Fearn (2003)<sup>71</sup> | Eur J Vasc Endovasc Surg | United Kingdom | no | no | symptomatic | CEA | Patients undergoing urological surgery | 8 weeks | 179 | 1 | 159 | 68±9.6 | 29 | 20 | 74<sup>b</sup> | NA | Global cognition<br>Processing speed<br>Attention<br>Memory<br>Visual spatial |
| Bossema (2005)<sup>72</sup> | Journal of vascular surgery | Netherlands | no | no | 36% asymptomatic | CEA | Patients with remote endarterectomy of the superficial femoral artery | 3 months +12 months | 83 | 4 | 60 | 66.2±7.2 | 16 | 23 | 65.6±8 | 13 | Attention<br>Memory<br>Processing speed<br>Executive functioning<br>Motor speed |
| Lehrner (2005)<sup>73</sup> | Journal of Clinical and Experimental Neuropsychology | Austria | no | no | 55% asymptomatic | CAS | Patients without surgery** | 3 months +6 months | 35 | 2 | 20 | 65.7±10.7 | 30 | 15 | 70.4±8.1 | 46.7 | Attention<br>Language<br>Processing speed<br>Executive functioning |
| Aleksic (2006)<sup>74</sup> | Eur J Vasc Endovasc Surg | Germany | no | NA | asymptomatic | CEA | Patients with peripheral arterial occlusive disease | 3-10 months | 58 | 14 | 33 | 68±8 | 42.4 | 25 | 62±11 | 40 | Memory<br>Executive functioning |
| Bossema (2007)<sup>75</sup> | Journal of clinical and experimental neuropsychology | Netherlands | no | no | asymptomatic | CEA | Healthy male participants with right-hand preference | 3 months | 70 | 0 | 45 | 66.6±7.2 | 0 | 25 | 66.7±4.7 | 0 | Language<br>Motor speed<br>Memory |
| Xu (2007)<sup>76</sup> | Neurological Research | China | no | single | both | CAS | Patients with similar medical condition undergoing carotid angiography | 12 weeks | 120 | 16 | 54 | 65.8±7.9 | 22.2 | 66 | 65.2±7.1 | 22.7 | Global cognition<br>Memory<br>Attention<br>Language<br>Processing speed<br>Executive functioning<br>Visual spatial<br>Motor speed |
| Jansen (2008)<sup>77</sup> | Der Nervenarzt | Germany | no | no | symptomatic | 37%CEA | Healthy volunteers | 4-6 weeks + 6 months | 40 | 0 | 27 | 69±7.5 | 14.8 | 13 | 64.3±6.3 | 31 | Attention<br>Memory |
| Soinne (2009)<sup>78</sup> | Brain and Cognition | Finland | no | single | 52% asymptomatic | CEA | Healthy volunteers | 100 days | 66 | 0 | 44 | 65.3±8.4 | 36.4 | 22 | 67±8.9 | 40.9 | Global cognition<br>Executive functioning |
| Czerny (2010)<sup>79</sup> | Journal of vascular surgery | Austria | no | no | 40% asymptomatic | CEA | Patient with no history or evidence of systemic atherosclerosis | 1 year + 5 years | 50 | 0 | 25 | 68±11 | 36 | 25 | 70±13 | 32 | Global cognition<br>Processing speed |
| Grunwald (2010)<sup>80</sup> | Neuroradiology | Germany | no | no | asymptomatic | CAS | Patients with incidental anterior communicating artery aneurysms | 3 months | 48 | 0 | 41 | 68.4<sup>a</sup> | 26.8 | 7 | 54<sup>a</sup> | 57.1 | Processing speed<br>Attention<br>Memory<br>Visual spatial |
| Huang (2011)<sup>81</sup> | Eur Neurol | China | no | no | 69% asymptomatic | CAS | Patients without surgery** | 6-12 months | 131 | 0 | 99 | 69.5±8.7 | 13.1 | 32 | 68.9±9.9 | 6 | Global cognition |
| Baracchini (2012)<sup>82</sup> | Surgery | Italy | no | single | 48% asymptomatic | CEA | Patients undergoing LC for gallbladder disease | 3 months +12 months | 213 | 0 | 145 | 74.7±6.4 | 30.3 | 68 | 73.9±7.4 | 25 | Global cognition |
| Yakhno (2012)<sup>83</sup> | Neuroscience and Behavioral Physiology | Russia | no | no | symptomatic | CAS | Patients without surgery** | 6 months +12 months | 56 | 0 | 35 | 63.2±7.3 (all) | 57.8 (all) | 21 | / | / | Attention<br>Language<br>Memory |
| Cheng (2013)<sup>84</sup> | Experimental and therapeutic medicine | China | no | single | both | CAS | Patients without surgery** | 6 months | 240 | 32 | 167 | 67±7.8 | 33.3 | 73 | 69.3±7.7 | 34.4 | Global cognition<br>Attention<br>Memory |
| Fan (2014)<sup>85</sup> | Vascular and Endovascular Surgery | China | no | no | symptomatic | CAS | Patients without surgery** | 1 month +3 months +6 months | 40 | 0 | 22 | 65.2±4.2 | 11.1 | 18 | 65.27±4.4 | 13.6 | Global cognition |
| Shi (2014)<sup>86</sup> | Chin J Cerebrovasc Dis | China | no | no | asymptomatic | CAS | Patients without surgery** | 3 months | 43 | 0 | 31 | 66±9 (all) | 16.28 (all) | 12 | / | / | Global cognition |
| Carta (2015)<sup>87</sup> | BMC Psychiatry | Italy | no | no | 67% asymptomatic | CEA | Patients without surgery** | 6 months | 40 | 0 | 30 | 73.2±7.2 | 43.3 | 10 | 72.1±8.8 | 20 | Global cognition<br>Language<br>Processing speed |
| Plessers (2015)<sup>88</sup> | Journal of Clinical and Experimental Neuropsychology | Belgium | no | no | 61% asymptomatic | 56.5%CEA | Patients with peripheral arterial disease | 12 months | 72 | 32 | 46 | 68.0±7.4 | 39.1 | 26 | 67.3±7.1 | 35 | Global cognition |
| Qu (2015)<sup>89</sup> | Journal of vascular surgery | China | no | no | symptomatic | CEA | Patients without surgery** | 3 months | 80 | 0 | 40 | 71.4±6.1 | 15 | 40 | 72.4±5.1 | 20 | Global cognition |
| Wapp (2015)<sup>90</sup> | Swiss Medical Weekly | Switzerland | no | no | 53% asymptomatic | 66.7%CEA | Patients without surgery** | 12 months | 58 | 0 | 30 | 67.6±8.4 | 26.7 | 28 | 71.3±8.1 | 25 | Memory<br>Language<br>Processing speed<br>Executive functioning |
| Yoon (2015)<sup>91</sup> | J Clin Neurol | South Korea | no | no | 52% asymptomatic | CAS | Patients without surgery** | 3 months | 33 | 0 | 23 | 67.7±8.5 | 17.4 | 10 | 61.1±7 | 40 | Global cognition<br>Memory<br>Attention<br>Language<br>Executive functioning<br>Visual spatial<br>Motor speed |
| Zhao (2015)<sup>93</sup> | Neuropsychiatric Disease and Treatment | China | no | no | symptomatic | CAS | Patients with non-stent-assisted angioplasty | 4 weeks+8 weeks | 122 | 0 | 78 | 60.8±10 | 42.3 | 44 | 56.9±9.3 | 40.9 | Global cognition |
| Kojima (2016)<sup>94</sup> | Neurological Research | Japan | no | single | asymptomatic | CEA | Healthy volunteers | 1 month | 167 | 6 | 116 | 67.3±4.8 | 9 | 51 | 68.4±5.1 | 20 | Global cognition<br>Language<br>Memory<br>Executive functioning |
| Shi, G.M. (2016)<sup>95</sup> | Current Neurovascular Research | China | no | single | NA | 49%CEA | Patients without surgery** | 3 months | 235 | 0 | 158 | 68.1±9.0 | 13.3 | 77 | 68.7±9.9 | 11.7 | Global cognition |
| Yue (2016)<sup>92</sup> | Biomedical Research | China | no | single | symptomatic | CAS | Patients without surgery** | 127 days | 105 | 0 | 83 | 64±8.3 | 14.7 | 22 | 61.7±9.8 | 28.6 | Global cognition<br>Memory<br>Attention<br>Language<br>Executive functioning<br>Visual spatial |
| Zhang (2016)<sup>96</sup> | Turk Neurosurg | China | no | no | 31% asymptomatic | CEA | Patients with femoropopliteal occlusive diseases | 3 months | 67 | 4 | 36 | 67±9 | 31 | 31 | 65±8 | 29 | Global cognition<br>Visual spatial |
| Lu (2017)<sup>97</sup> | Natl Med J China | China | no | no | symptomatic | CAS | Patients with conservative treatment | 6 months | 33 | 3 | 17 | 68±7 | 6.7 | 16 | 68±8 | 20 | Global cognition<br>Memory<br>Attention<br>Processing speed<br>Executive functioning<br>Visual spatial |
| Watanabe (2017)<sup>98</sup> | Journal of stroke and cerebrovascular diseases | Japan | no | no | both | 78%CEA | Patients without surgery** | 12 months | 73 | 0 | 50 | 73.3±6.8 | 8 | 23 | 67.5±4.0 | 30.4 | Global cognition<br>Memory<br>Attention<br>Language<br>Executive functioning<br>Visual spatial |
| Belov (2018)<sup>99</sup> | N.I. Pirogova | Russia | no | no | 35% asymptomatic | CEA | Patients without surgery** | 3 months +6 months | 120 | 0 | 100 | 65.4±9.0 | 28 | 20 | 67.8±5.3 | 25 | Global cognition<br>Executive functioning |
| Pucite (2019)<sup>100</sup> | Current Neurovascular Research | Latvia | no | single | 73% asymptomatic | 85%CEA | Patients without surgery** | 6 months +12 months | 213 | 0 | 188 | 69.8±9.2 | 38.8 | 25 | 73±8.1 | 24 | Global cognition<br>Memory<br>Attention<br>Language<br>Executive functioning |
| Song (2019)<sup>101</sup> | Journal of the Neurological Sciences | China | no | no | symptomatic | CAS | Patients without surgery** | 1 month +6 months +12 months | 79 | 0 | 48 | 70.5±3.3 | 27.1 | 31 | 69.8±3.1 | 41.9 | Global cognition<br>Memory<br>Attention<br>Language<br>Executive functioning<br>Visual spatial |
| Thirumala (2020)<sup>102</sup> | Clinical Neurology and Neurosurgery | USA | no | no | 81% asymptomatic | CEA | Patients without surgery** | 8+13,4 years | 5888 | 3790 | 167 | 71.5±4.7 | 41.3 | 5721 | 72.9±5.6 | 58.1 | Global cognition |
| Whooley (2020)<sup>103</sup> | Journal of Stroke and Cerebrovascular Diseases | USA | no | no | asymptomatic | 83%CEA | Patients with conservative treatment | 1 month +6 months +12 months | 75 | 0 | 64 | 71±8.5 (all) | 36.8 (all) | 11 | / | / | Memory<br>Language<br>Processing speed<br>Executive functioning |
| Matyushkin (2021)<sup>104</sup> | Pirogov Journal of Surgery | Russia | no | no | symptomatic | CEA/CAS | Patients without surgery** | 12 months | 186 | 0 | 136 | 65±2 (all) | 34.4 (all) | 50 | / | / | Global cognition |
| Turowicz (2021)<sup>105</sup> | Journal of Stroke and Cerebrovascular Diseases | Poland | no | single | asymptomatic | 71%CEA | Patients with lower extremity artery disease AND inguinal hernia surgery | 6 months | 90 | 8 | 70 | 67.81<sup>a</sup> | 34.3 | 20 | 68.4<sup>a</sup> | 30 | Global cognition<br>Memory<br>Attention<br>Language<br>Processing speed<br>Executive functioning<br>Visual spatial |
| Huang (2021)<sup>106</sup> | Frontiers in Aging Neuroscience | China | no | single | symptomatic | CAS | Patients with conservative treatment | 6 months | 85 | 0 | 64 | 66.7±8.4 | 4.7 | 21 | 64.9±4.2 | 20 | Visual spatial<br>Memory<br>Executive functioning<br>Language<br>Processing speed |
| Lin, C.J. (2022)<sup>107</sup> | Journal of the Formosan Medical Association | China | no | single | asymptomatic | CAS | Patients without surgery** | 3 months | 69 | 23 | 38 | 67.8±9.4 | 15.8 | 31 | 68±8.4 | 16.1 | Global cognition<br>Memory<br>Attention<br>Processing speed<br>Executive functioning<br>Visual spatial |
| Relander (2022)<sup>108</sup> | Frontiers in Neurology | Finland | no | no | 52% asymptomatic | CEA | Healthy volunteers | 3 months | 144 | 0 | 44 | 63.8±8.8 | 36 | 100 | 60.4±8.6 | 14 | Memory<br>Processing speed<br>Executive functioning<br>Motor speed |
| Tang (2023)<sup>109</sup> | Scientific Reports | China | no | single | symptomatic | HS | Patients without surgery** | 6 months | 80 | 0 | 40 | 65.1±8.4 | 20 | 40 | 63.1±8.2 | 25 | Global cognition<br>Language<br>Processing speed<br>Executive functioning |
| Xiao (2023)<sup>110</sup> | Neuropsychiatric Disease and Treatment | China | no | no | symptomatic | CAS | Patients without surgery** | 3 months | 41 | 8 | 20 | 61.3±8.4 | 25 | 21 | 63.8±9.3 | 19 | Global cognition |
| Bernstein (2024)<sup>111</sup> | Journal of Stroke and Cerebrovascular Diseases | USA | no | no | asymptomatic | CEA | Patients with < 70% stenosis or occlusion of at least one carotid artery | 4-6 months | 26 | 0 | 13 | 69.9±8.1 | 23.1 | 13 | 68.8±8.6 | 30.8 | Global cognition<br>Executive functioning<br>Visual spatial |
| Relander (2024)<sup>112</sup> | Journal of the Neurological Sciences | Finland | no | Single | 44% asymptomatic | CEA | Healthy volunteers | 6 months | 98 | 5 | 60 | 68.7±6.7 | 16 | 38 | 67.4±5.4 | 24 | Global cognition<br>Memory<br>Attention<br>Processing speed<br>Executive functioning<br>Motor speed |

Legend: SD= standard deviation; CEA = carotid endarterectomy; CAS = carotid artery stenting; INT= intervention; REF= reference. NA= not available; HS= hybrid surgery (A surgery combines CAS and CEA); all = the combined intervention and reference groups. \*VRF including heart disease, Hypertension, Type 2 diabetes mellitus, smoking. \*\* Patients from the same source population as the intervention group did not undergo carotid intervention. a : The article only provides the mean of age, no information for SD. b: The article only provides the median of the age, no information for mean and SD.

**Table 4:**
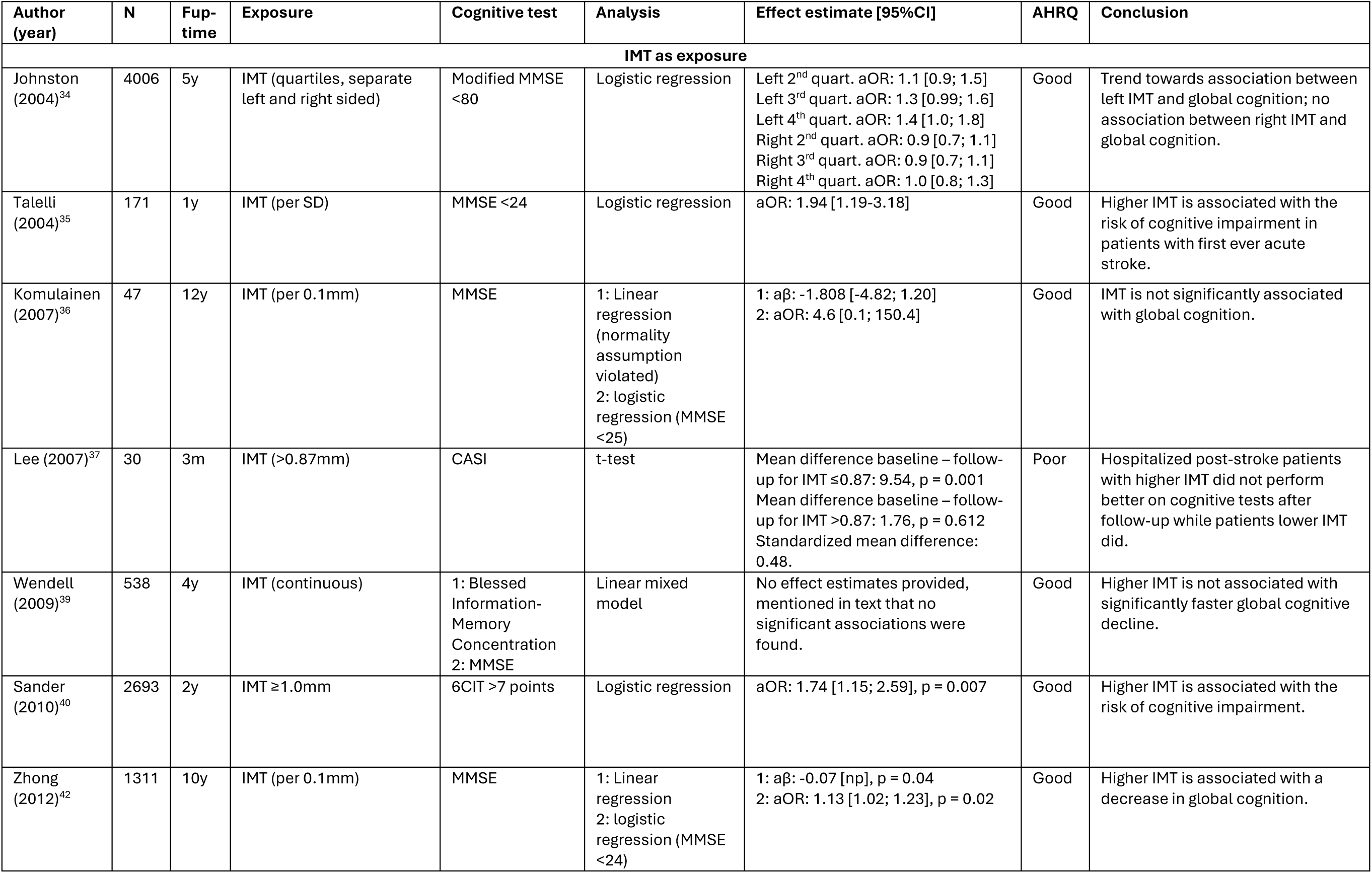
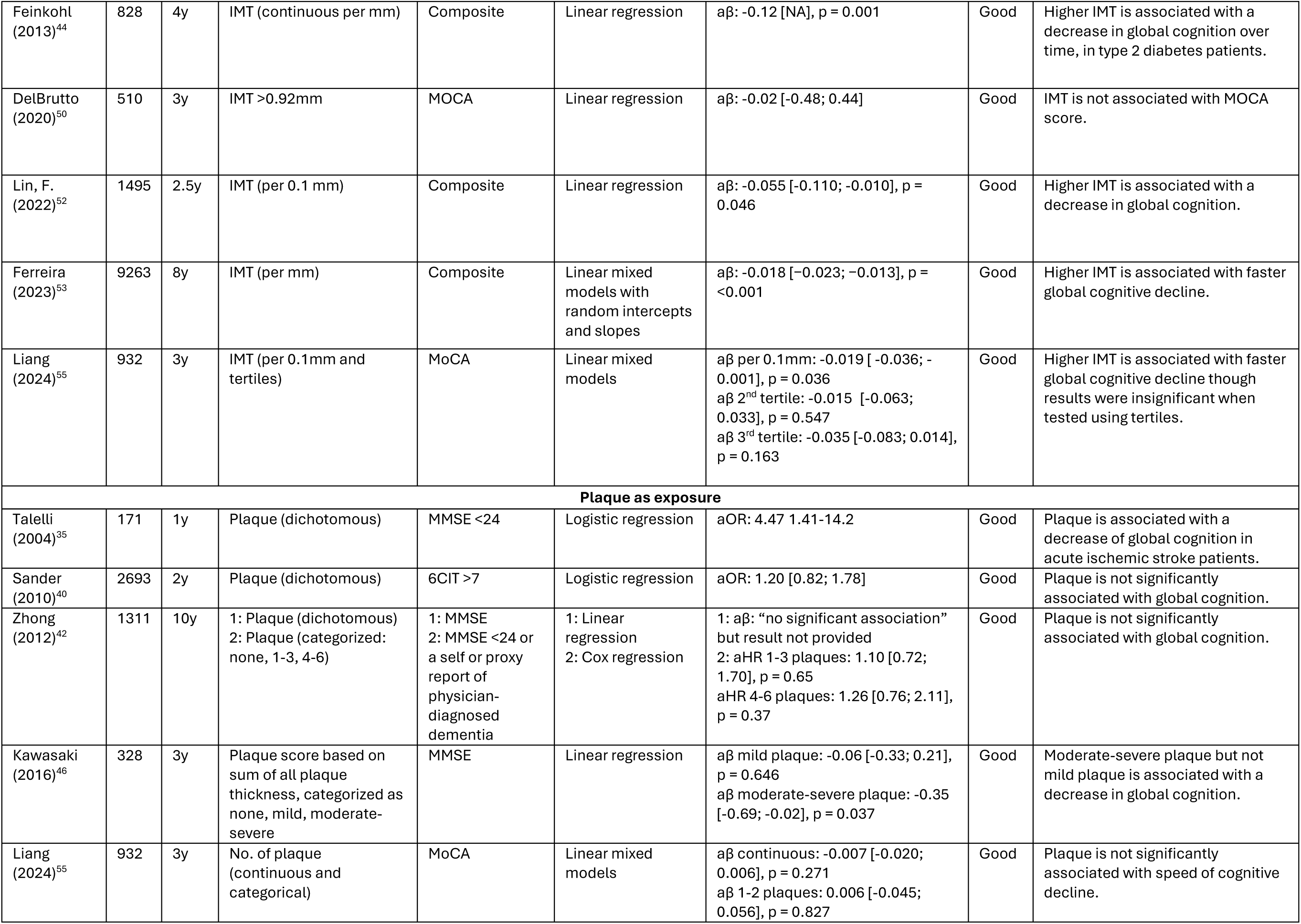
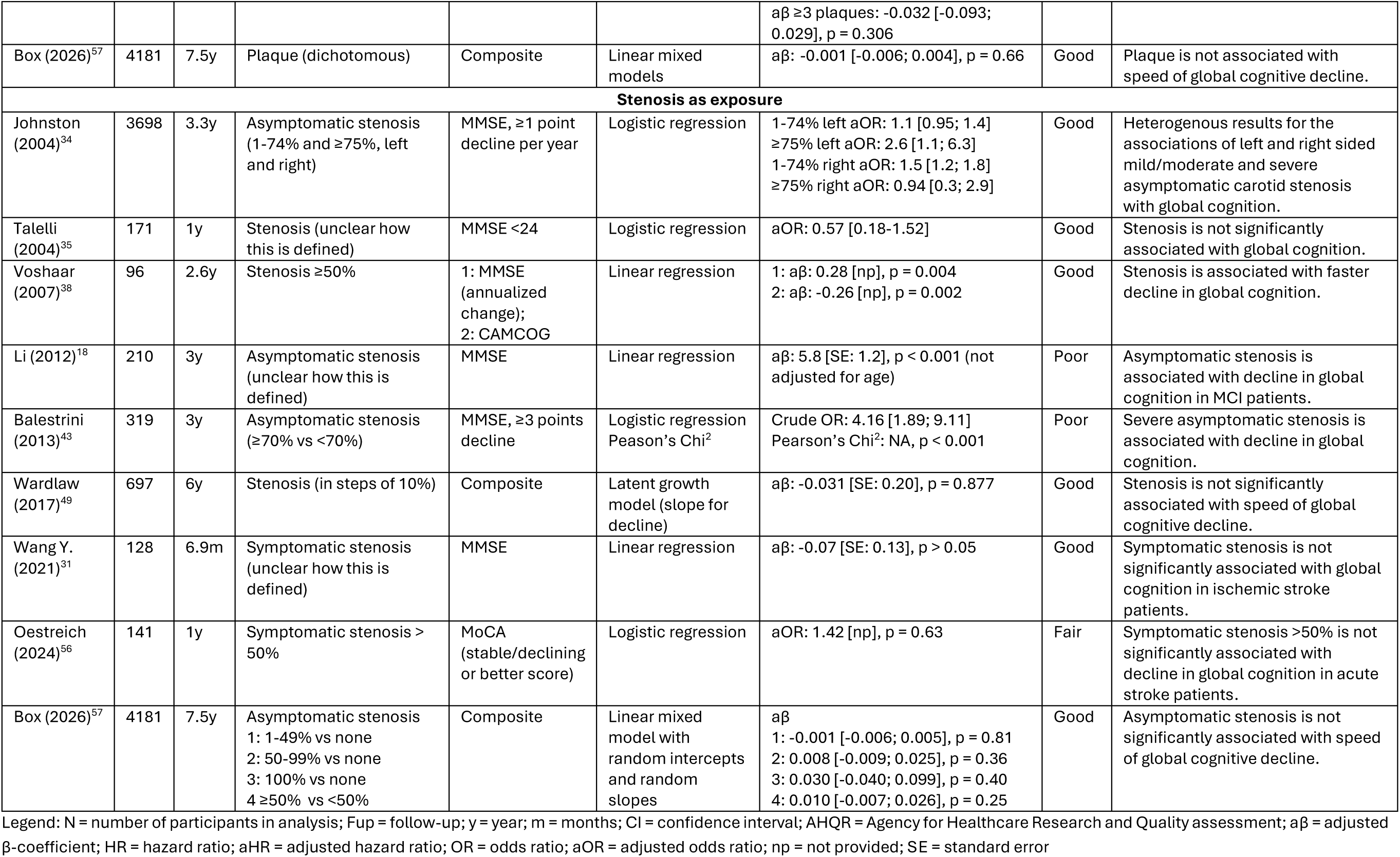
Association of carotid atherosclerosis with cognition.

| Author (year) | N | Fup-time | Exposure | Cognitive test | Analysis | Effect estimate [95%CI] | AHRQ | Conclusion |
|---|---|---|---|---|---|---|---|---|
| **IMT as exposure** |  |  |  |  |  |  |  |  |
| Johnston (2004)<sup>34</sup> | 4006 | 5y | IMT (quartiles, separate left and right sided) | Modified MMSE <80 | Logistic regression | Left 2<sup>nd</sup> quart. aOR: 1.1 [0.9; 1.5]<br>Left 3<sup>rd</sup> quart. aOR: 1.3 [0.99; 1.6]<br>Left 4<sup>th</sup> quart. aOR: 1.4 [1.0; 1.8]<br>Right 2<sup>nd</sup> quart. aOR: 0.9 [0.7; 1.1]<br>Right 3<sup>rd</sup> quart. aOR: 0.9 [0.7; 1.1]<br>Right 4<sup>th</sup> quart. aOR: 1.0 [0.8; 1.3] | Good | Trend towards association between left IMT and global cognition; no association between right IMT and global cognition. |
| Talelli (2004)<sup>35</sup> | 171 | 1y | IMT (per SD) | MMSE <24 | Logistic regression | aOR: 1.94 [1.19-3.18] | Good | Higher IMT is associated with the risk of cognitive impairment in patients with first ever acute stroke. |
| Komulainen (2007)<sup>36</sup> | 47 | 12y | IMT (per 0.1mm) | MMSE | 1: Linear regression (normality assumption violated)<br>2: logistic regression (MMSE <25) | 1: aβ: -1.808 [-4.82; 1.20]<br>2: aOR: 4.6 [0.1; 150.4] | Good | IMT is not significantly associated with global cognition. |
| Lee (2007)<sup>37</sup> | 30 | 3m | IMT (>0.87mm) | CASI | t-test | Mean difference baseline – follow-up for IMT ≤0.87: 9.54, p = 0.001<br>Mean difference baseline – follow-up for IMT >0.87: 1.76, p = 0.612<br>Standardized mean difference: 0.48. | Poor | Hospitalized post-stroke patients with higher IMT did not perform better on cognitive tests after follow-up while patients lower IMT did. |
| Wendell (2009)<sup>39</sup> | 538 | 4y | IMT (continuous) | 1: Blessed Information-Memory Concentration<br>2: MMSE | Linear mixed model | No effect estimates provided, mentioned in text that no significant associations were found. | Good | Higher IMT is not associated with significantly faster global cognitive decline. |
| Sander (2010)<sup>40</sup> | 2693 | 2y | IMT ≥1.0mm | 6CIT >7 points | Logistic regression | aOR: 1.74 [1.15; 2.59], p = 0.007 | Good | Higher IMT is associated with the risk of cognitive impairment. |
| Zhong (2012)<sup>42</sup> | 1311 | 10y | IMT (per 0.1mm) | MMSE | 1: Linear regression<br>2: logistic regression (MMSE <24) | 1: aβ: -0.07 [np], p = 0.04<br>2: aOR: 1.13 [1.02; 1.23], p = 0.02 | Good | Higher IMT is associated with a decrease in global cognition. |
| Feinkohl (2013)<sup>44</sup> | 828 | 4y | IMT (continuous per mm) | Composite | Linear regression | aβ: -0.12 [NA], p = 0.001 | Good | Higher IMT is associated with a decrease in global cognition over time, in type 2 diabetes patients. |
| DelBrutto (2020)<sup>50</sup> | 510 | 3y | IMT >0.92mm | MOCA | Linear regression | aβ: -0.02 [-0.48; 0.44] | Good | IMT is not associated with MOCA score. |
| Lin, F. (2022)<sup>52</sup> | 1495 | 2.5y | IMT (per 0.1 mm) | Composite | Linear regression | aβ: -0.055 [-0.110; -0.010], p = 0.046 | Good | Higher IMT is associated with a decrease in global cognition. |
| Ferreira (2023)<sup>53</sup> | 9263 | 8y | IMT (per mm) | Composite | Linear mixed models with random intercepts and slopes | aβ: -0.018 [−0.023; −0.013], p = <0.001 | Good | Higher IMT is associated with faster global cognitive decline. |
| Liang (2024)<sup>55</sup> | 932 | 3y | IMT (per 0.1mm and tertiles) | MoCA | Linear mixed models | aβ per 0.1mm: -0.019 [ -0.036; -0.001], p = 0.036<br>aβ 2<sup>nd</sup> tertile: -0.015 [-0.063; 0.033], p = 0.547<br>aβ 3<sup>rd</sup> tertile: -0.035 [-0.083; 0.014], p = 0.163 | Good | Higher IMT is associated with faster global cognitive decline though results were insignificant when tested using tertiles. |
| **Plaque as exposure** |  |  |  |  |  |  |  |  |
| Talelli (2004)<sup>35</sup> | 171 | 1y | Plaque (dichotomous) | MMSE <24 | Logistic regression | aOR: 4.47 1.41-14.2 | Good | Plaque is associated with a decrease of global cognition in acute ischemic stroke patients. |
| Sander (2010)<sup>40</sup> | 2693 | 2y | Plaque (dichotomous) | 6CIT >7 | Logistic regression | aOR: 1.20 [0.82; 1.78] | Good | Plaque is not significantly associated with global cognition. |
| Zhong (2012)<sup>42</sup> | 1311 | 10y | 1: Plaque (dichotomous)<br>2: Plaque (categorized: none, 1-3, 4-6) | 1: MMSE<br>2: MMSE <24 or a self or proxy report of physician-diagnosed dementia | 1: Linear regression<br>2: Cox regression | 1: aβ: “no significant association” but result not provided<br>2: aHR 1-3 plaques: 1.10 [0.72; 1.70], p = 0.65<br>aHR 4-6 plaques: 1.26 [0.76; 2.11], p = 0.37 | Good | Plaque is not significantly associated with global cognition. |
| Kawasaki (2016)<sup>46</sup> | 328 | 3y | Plaque score based on sum of all plaque thickness, categorized as none, mild, moderate-severe | MMSE | Linear regression | aβ mild plaque: -0.06 [-0.33; 0.21], p = 0.646<br>aβ moderate-severe plaque: -0.35 [-0.69; -0.02], p = 0.037 | Good | Moderate-severe plaque but not mild plaque is associated with a decrease in global cognition. |
| Liang (2024)<sup>55</sup> | 932 | 3y | No. of plaque (continuous and categorical) | MoCA | Linear mixed models | aβ continuous: -0.007 [-0.020; 0.006], p = 0.271<br>aβ 1-2 plaques: 0.006 [-0.045; 0.056], p = 0.827<br>aβ ≥3 plaques: -0.032 [-0.093; 0.029], p = 0.306 | Good | Plaque is not significantly associated with speed of cognitive decline. |
| Box (2026)<sup>57</sup> | 4181 | 7.5y | Plaque (dichotomous) | Composite | Linear mixed models | aβ: -0.001 [-0.006; 0.004], p = 0.66 | Good | Plaque is not associated with speed of global cognitive decline. |
| **Stenosis as exposure** |  |  |  |  |  |  |  |  |
| Johnston (2004)<sup>34</sup> | 3698 | 3.3y | Asymptomatic stenosis (1-74% and ≥75%, left and right) | MMSE, ≥1 point decline per year | Logistic regression | 1-74% left aOR: 1.1 [0.95; 1.4]<br>≥75% left aOR: 2.6 [1.1; 6.3]<br>1-74% right aOR: 1.5 [1.2; 1.8]<br>≥75% right aOR: 0.94 [0.3; 2.9] | Good | Heterogenous results for the associations of left and right sided mild/moderate and severe asymptomatic carotid stenosis with global cognition. |
| Talelli (2004)<sup>35</sup> | 171 | 1y | Stenosis (unclear how this is defined) | MMSE <24 | Logistic regression | aOR: 0.57 [0.18-1.52] | Good | Stenosis is not significantly associated with global cognition. |
| Voshaar (2007)<sup>38</sup> | 96 | 2.6y | Stenosis ≥50% | 1: MMSE (annualized change);<br>2: CAMCOG | Linear regression | 1: aβ: 0.28 [np], p = 0.004<br>2: aβ: -0.26 [np], p = 0.002 | Good | Stenosis is associated with faster decline in global cognition. |
| Li (2012)<sup>18</sup> | 210 | 3y | Asymptomatic stenosis (unclear how this is defined) | MMSE | Linear regression | aβ: 5.8 [SE: 1.2], p < 0.001 (not adjusted for age) | Poor | Asymptomatic stenosis is associated with decline in global cognition in MCI patients. |
| Balestrini (2013)<sup>43</sup> | 319 | 3y | Asymptomatic stenosis (≥70% vs <70%) | MMSE, ≥3 points decline | Logistic regression<br>Peason’s Chi<sup>2</sup> | Crude OR: 4.16 [1.89; 9.11]<br>Pearson’s Chi<sup>2</sup>: NA, p < 0.001 | Poor | Severe asymptomatic stenosis is associated with decline in global cognition. |
| Wardlaw (2017)<sup>49</sup> | 697 | 6y | Stenosis (in steps of 10%) | Composite | Latent growth model (slope for decline) | aβ: -0.031 [SE: 0.20], p = 0.877 | Good | Stenosis is not significantly associated with speed of global cognitive decline. |
| Wang Y. (2021)<sup>31</sup> | 128 | 6.9m | Symptomatic stenosis (unclear how this is defined) | MMSE | Linear regression | aβ: -0.07 [SE: 0.13], p > 0.05 | Good | Symptomatic stenosis is not significantly associated with global cognition in ischemic stroke patients. |
| Oestreich (2024)<sup>56</sup> | 141 | 1y | Symptomatic stenosis > 50% | MoCA (stable/declining or better score) | Logistic regression | aOR: 1.42 [np], p = 0.63 | Fair | Symptomatic stenosis >50% is not significantly associated with decline in global cognition in acute stroke patients. |
| Box (2026)<sup>57</sup> | 4181 | 7.5y | Asymptomatic stenosis<br>1: 1-49% vs none<br>2: 50-99% vs none<br>3: 100% vs none<br>4 ≥50% vs <50% | Composite | Linear mixed model with random intercepts and random slopes | aβ<br>1: -0.001 [-0.006; 0.005], p = 0.81<br>2: 0.008 [-0.009; 0.025], p = 0.36<br>3: 0.030 [-0.040; 0.099], p = 0.40<br>4: 0.010 [-0.007; 0.026], p = 0.25 | Good | Asymptomatic stenosis is not significantly associated with speed of global cognitive decline. |

Legend: N = number of participants in analysis; Fup = follow-up; y = year; m = months; CI = confidence interval; AHQR = Agency for Healthcare Research and Quality assessment; aβ = adjusted β-coefficient; HR = hazard ratio; aHR = adjusted hazard ratio; OR = odds ratio; aOR = adjusted odds ratio; np = not provided; SE = standard error

Of the 35 eligible studies assessing global cognition, 22 were included in the meta-analysis,^66,67,71,76,82,84–87,89,92–97,100,101,104,107,109,110^ whereas the remaining 13 were for qualitative synthesis only.^69,70,78,79,81,88,91,98,99,102,105,111,112^ Nine did not report the mean or standard deviation required to calculate SMDs,^70,78,79,81,88,98,99,105,111^ one reported annualized cognitive change rates over variable follow-up,^98^ and three reported adjusted means and standard deviations that could not be pooled with unadjusted data ^65,87,108^. In the meta-analysis, revascularization was associated with a moderate improvement in global cognition (SMD: 0.73 [0.37–1.09]; I^2^=89.8%; 22 studies, Figure 2A). Meta-regression identified the reference-group type as a significant moderator, with larger cognitive effects in studies using eligible non-revascularized patients than healthy or other surgical controls. (SMD_difference_: 0.93 [0.16–1.69]; p= 0.018, Figure 3). Consistent with this finding, studies with carotid stenosis controls showed larger effects than those with other controls (SMD: 0.94 [0.53–1.36]; I^2^=90.5%; 17 studies; vs 0.07 [-0.24 to 0.38]; I^2^=51.4; 5 studies; p_difference_=0.0009, Figure S6). No significant differences in intervention effects were observed between CEA and CAS (SMD_difference_: 0.57 [−0.26 to 1.40]; p=0.176) or between symptomatic and asymptomatic patients (SMD_difference_: 0.41 [−0.63 to 1.46]; p=0.438), and effect estimates were not significantly associated with follow-up duration, age, or sex composition. Effects tended to be somewhat larger in Asian populations than in non-Asian populations (SMD_difference_: 0.68 [-0.05–1.41], p=0.068). In multivariable analyses, the association with reference-group type remained after adjustment for symptomatic status (SMD_difference_:0.90 [0.14-1.66]; p=0.020) but was no longer statistically significant after adjustment for intervention type (SMD_difference_: 1.11 [−0.09 to 2.30]; p=0.070).

**Figure 3:**
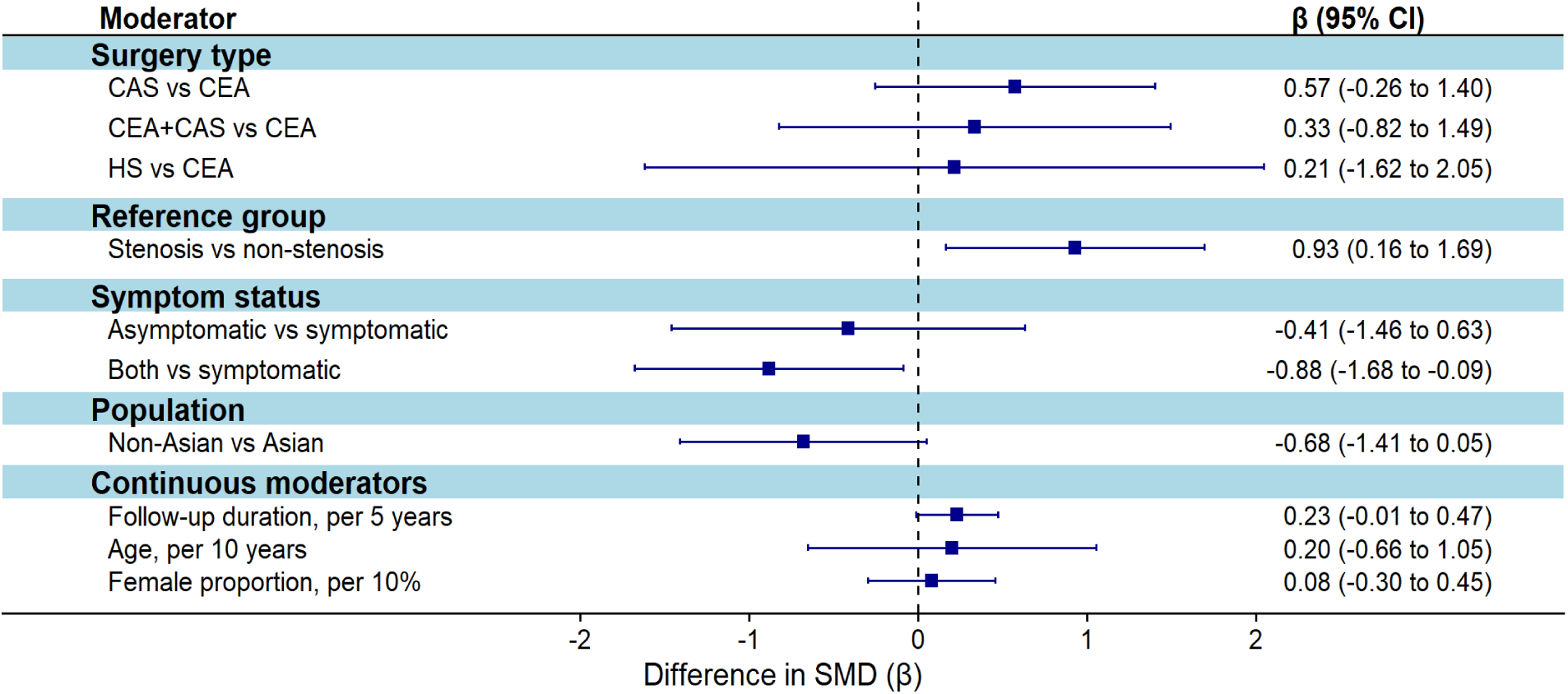
Effect modifiers for cognitive change after carotid revascularization in univariable meta-regression. Legend: For categorical moderators, the group after “vs” serves as the reference, β = changes in the SMD associated with each moderator, positive β values indicate higher SMDs relative to the reference group. CEA = carotid endarterectomy; CAS = carotid artery stenting; HS = hybrid surgery; CI = confidence interval; SMD = standardized mean difference.

For individual cognitive domains, 13 of 21 studies assessing attention provided sufficient data for meta-analysis, as did 17/26 for memory, 16/28 for executive functioning, 11/21 for processing speed, 12/20 for language, 8/16 for visuospatial function, and 5/8 for motor speed. Overall, SMDs for different domains were substantially smaller than those observed for global cognition (Figure S8), significant improvement was observed only for visuospatial function (SMD:0.35 [0.06–0.64]).

In sensitivity analyses, findings were robust to varying the assumed within-study correlation, the robust variance estimation correlation parameter, and trimming of extreme values (Tables S8–S10). Funnel plots showed some asymmetry (Figure S10), with the standard Egger test suggesting publication bias for the language and visuospatial domains. These findings were not significant in the cluster-robust Egger test (Table S11), providing no clear evidence of publication bias overall.

## Discussion

In this comprehensive systematic review and meta-analysis, carotid atherosclerosis was associated with higher risk of dementia, consistent across different measures of atherosclerosis. However, few studies determined the effect of stenosis on dementia, especially for ≥70% stenosis. Results of studies on cognition were generally consistent with those on dementia. Revascularization improved cognition compared to reference subjects in non-randomized intervention studies, but with large, mostly unexplained heterogeneity, and high risk of bias. Evidence from six randomized controlled trials was inconsistent.

Building on earlier systematic reviews of mostly cross-sectional data covering data up till 2022,^6,7^ the current summary of longitudinal studies strengthens evidence that carotid atherosclerosis contributes causally to dementia risk and –albeit with more heterogeneity– cognitive decline. These results appeared similar for measurements of IMT, plaque and stenosis. As no published studies jointly analyzed stenosis and atherosclerosis, it remains undetermined whether observations for carotid stenosis are reflective of stenosis degree or broader burden of atherosclerosis. In a separately published analysis of the Rotterdam Study cohort,^57^ we found that risk estimates for stenosis did not exceed those of plaque presence without stenosis, suggesting the underlying atherosclerosis may be the primary driver of dementia risk. In the Rotterdam Study cohort, carotid occlusion increased dementia risk to a larger extent than 70-99% stenosis, suggesting that reported risk estimates for steno-occlusive disease in this review could be explained, at least in part, by the inclusion of individuals with 100% stenosis, not amenable by revascularization. Although these results from the Rotterdam Study seem to support the hypothesis that carotid stenosis affects cognition through hemodynamic impairment, the CREST-H substudy found no effect of revascularization in patients with asymptomatic carotid artery stenosis and proven hemodynamic impairment.^64^

The neutral results of the larger ACAS, ACST-1 and particularly CREST-2 trials temper enthusiasm about benefit of revascularization on cognition. Two smaller trials among Asian patients did report positive effects.^58,60–63^ This may reflect genuine clinical heterogeneity between populations or healthcare systems, supported by somewhat larger effect estimates for Asian populations in meta regression analyses of non-randomized interventions. At the same time, smaller trials are at higher likelihood to overestimate the effect (i.e., the winner’s curse). The large degree of heterogeneity in RCTs cautions against interpretation of pooled estimates, as it leads the inverse variance weighting in random effects models to virtually ignore within-study variance, causing the weights of the different trials to converge to equality (∼25% each) despite 10-fold differences in sample size. From a methodological perspective, differences in follow-up duration may have contributed to the observed heterogeneity, as the two smaller trials assessed relatively short-term cognitive changes compared with CREST-2. Similarly, the apparent benefit of revascularization observed in non-randomized intervention studies should be interpreted with caution, as all studies were at critical risk of bias and heterogeneity was high. Risk of bias was due mostly to the lack of randomization, missing data, and selection of non-comparable controls. Indeed, meta-regression showed that reference-group type influenced the observed effect estimates. Other unmeasured sources of heterogeneity may include broader carotid atherosclerotic disease and baseline carotid stenosis severity. Although eligibility criteria described stenosis severity, available data were insufficiently standardized for assessment as an effect modifier. Revascularization seemed to have a larger effect on global cognition than on individual cognitive domains, which may reflect cumulative effects across domains, or more robust assessment of global cognitive function. Overall, the contrasting results from randomized controlled trials and non-randomized intervention studies indeed suggest that the cognitive improvements observed in non-randomized studies may at least partly reflect confounding and other sources of bias, and all in all current evidence is insufficient to support carotid revascularization for the purpose of improving cognition.

Our study extends previous work by including both symptomatic and asymptomatic populations and by incorporating evidence from both longitudinal observational and intervention studies. Several limitations also need to be considered. First, this systematic review focused solely on extracranial carotid atherosclerosis without considering the strong association with atherosclerosis in other locations. Observed associations of carotid atherosclerosis might be reflective of generalized atherosclerotic disease or intracranial arteriosclerosis. Neither did we investigate the effect of plaque characteristics, which are generally not assessed on ultrasound, the main imaging technique used in the included studies. Second, we were unable to differentiate based on ancestry, which may be of particular relevance given established differences in prevalence of extracranial vs intracranial carotid atherosclerosis. Third, heterogeneity in study methods on cognition restricted meta-analysis of the observational studies investigating cognition. Fourth, the limited number of non-randomized intervention studies may have reduced the power of subgroup and meta-regression analyses to detect meaningful effect modification. Fifth, studies reporting multiple tests within the same cognitive domain may have contributed disproportionately to the pooled estimates, although robust variance estimation sensitivity analyses showed only minor changes in standard errors.

In conclusion, carotid atherosclerosis is associated with increased risk of dementia, but current evidence does not support carotid revascularization for prevention of cognitive decline or dementia. The consistent link between carotid atherosclerosis and dementia risk underlines the importance of optimal risk factor management in patients with carotid atherosclerosis. Additional high quality, well-powered cohort studies and trials are needed to determine sources of heterogeneity across studies for long-term cognitive outcome data, accounting for plaque characteristics and hemodynamic consequences.

## Data Availability

All data used in this systematic review and meta-analysis were extracted from published studies. The data supporting the findings of this study are provided in the manuscript and its supplementary materials.

## Acknowledgements

The authors acknowledge the contributions of the employees of the Erasmus MC medical library for their help in designing the search strategy.

