## Supplementary Material for "The effect of carotid atherosclerosis and revascularization on cognition and dementia: a systematic review and meta-analysis"

### Supplements

#### Methods S1: Search strategies

Medline

(exp "Carotid Stenosis"/ OR exp "Endarterectomy, Carotid"/ OR (((caroti*) AND (atherosclero* OR atherom* OR plaque* OR stent* OR stenos* OR narrow* OR restenos* OR endarterectom* OR desobstruct* OR des-obstruct* OR endovascular*)) OR ((caroti*) ADJ6 (arter* OR occlusive*) ADJ3 (disease*))).ab,ti,kf. OR (((intracranial* OR intra-cranial*) AND (arter*) AND (atherosclero* OR atherom* OR plaque* OR stent* OR stenos* OR narrow* OR restenos* OR endarterectom* OR desobstruct* OR des-obstruct* OR endovascular* OR occlusion*))).ti.) AND (exp "Cognition Disorders"/ OR exp "Dementia"/ OR (((cognitive* OR neurocognitive* OR neurodegenerat* OR neuro-degenerat*) ADJ3 (decline* OR impair* OR defect* OR complaint* OR difficult* OR disorder* OR problem* OR dysfunct* OR deficien*)) OR dement* OR Alzheimer* OR Binswanger-encephalopath OR CADASIL OR Lewy-body OR diffuse-neurofibrillary-tangles-with-calcification* OR hereditary-diffuse-leukoencephalopath* OR Huntington-chorea OR Kluver-Bucy OR mental-deteriorat OR Nasu-Hakola OR neuronal-ceroid-lipofuscinosis OR prion-disease OR pseudodementi OR Rett-syndrom OR tauopath* OR ((cognit* OR neurocognit*) ADJ3 (function* OR change* OR performan* OR result*))).ab,ti,kf. OR (((cognit* OR neurocognit*) ADJ3 (outcome*))).ti.) NOT (news OR congres* OR abstract* OR book* OR chapter* OR dissertation abstract*).pt. NOT (exp animals/ NOT humans/)

Embase

('carotid stenosis'/exp OR 'carotid atherosclerosis'/exp OR 'carotid artery stent'/exp OR 'carotid endarterectomy'/exp OR (('endovascular treatment'/exp OR 'endovascular surgery'/exp OR 'atherosclerosis'/exp) AND ('carotid artery'/exp OR 'carotid artery disease'/exp)) OR 'carotid artery occlusion'/exp/mj OR (((caroti*) AND (atherosclero* OR atherom* OR plaque* OR stent* OR stenos* OR narrow* OR restenos* OR endarterectom* OR desobstruct* OR des-obstruct* OR endovascular*)) OR ((caroti*) NEAR/6 (arter* OR occlusive*) NEAR/3 (disease*))):ab,ti,kw OR (((intracranial* OR intra-cranial*) AND (arter*) AND (atherosclero* OR atherom* OR plaque* OR stent* OR stenos* OR narrow* OR restenos* OR endarterectom* OR desobstruct* OR des-obstruct* OR endovascular* OR occlusion*))):ti) AND ('cognitive defect'/exp OR (((cognitive* OR neurocognitive* OR neurodegenerat* OR neuro-degenerat*) NEAR/3 (decline* OR impair* OR defect* OR complaint* OR difficult* OR disorder* OR problem* OR dysfunct* OR deficien*)) OR dement* OR Alzheimer* OR Binswanger-encephalopath OR CADASIL OR Lewy-body OR diffuse-neurofibrillary-tangles-with-calcification* OR hereditary-diffuse-leukoencephalopath* OR Huntington-chorea OR Kluver-Bucy OR mental-deteriorat OR Nasu-Hakola OR neuronal-ceroid-lipofuscinosis OR prion-disease OR pseudodementi OR Rett-syndrom OR tauopath* OR ((cognit* OR neurocognit*) NEAR/3 (function* OR change* OR performan* OR result*))):ab,ti,kw OR (((cognit* OR neurocognit*) NEAR/3 (outcome*))):ti) NOT ([Conference Abstract]/lim OR [preprint]/lim) NOT ((animal/exp OR animal*:de OR nonhuman/de) NOT ('human'/exp))

Web of Science

(TS=(((caroti*) AND (atherosclero* OR atherom* OR plaque* OR stent* OR stenos* OR narrow* OR restenos* OR endarterectom* OR desobstruct* OR des-obstruct* OR endovascular*)) OR ((caroti*) NEAR/6 (arter* OR occlusive*) NEAR/3 (disease*))) OR TI=(((intracranial* OR intra-cranial*) AND (arter*) AND (atherosclero* OR atherom* OR plaque* OR stent* OR stenos* OR narrow* OR restenos* OR endarterectom* OR desobstruct* OR des-obstruct* OR endovascular* OR occlusion*)))) AND (TS=(((cognitive* OR neurocognitive* OR neurodegenerat* OR neuro-degenerat*) NEAR/3 (decline* OR impair* OR defect* OR complaint* OR difficult* OR disorder* OR problem* OR dysfunct* OR deficien*)) OR dement* OR Alzheimer* OR Binswanger-encephalopath OR CADASIL OR Lewy-body OR diffuse-neurofibrillary-tangles-with-calcification* OR hereditary-diffuse-leukoencephalopath* OR Huntington-chorea OR Kluver-Bucy OR mental-deteriorat OR Nasu-Hakola OR neuronal-ceroid-lipofuscinosis OR prion-disease OR pseudodementi OR Rett-syndrom OR tauopath* OR ((cognit* OR neurocognit*) NEAR/3 (function* OR change* OR performan* OR result*))) OR TI=(((cognit* OR neurocognit*) NEAR/3 (outcome*)))) NOT TS=(((animal* OR rat OR rats OR mouse OR mice OR murine OR dog OR dogs OR canine OR cat OR cats OR feline OR rabbit OR cow OR cows OR bovine OR rodent* OR sheep OR ovine OR pig OR swine OR porcine OR veterinar* OR chick* OR zebrafish* OR baboon* OR nonhuman* OR primate* OR cattle* OR goose OR geese OR duck OR macaque* OR avian* OR bird* OR fish*) NOT (human* OR patient* OR women OR woman OR men OR man))) NOT DT=(Meeting Abstract OR Meeting Summary)

Cochrane CENTRAL

((((caroti*) AND (atherosclero* OR atherom* OR plaque* OR stent* OR stenos* OR narrow* OR restenos* OR endarterectom* OR desobstruct* OR des NEXT/1 obstruct* OR endovascular*)) OR ((caroti*) NEAR/6 (arter* OR occlusive*) NEAR/3 (disease*))):ab,ti,kw OR (((intracranial* OR intra NEXT/1 cranial*) AND (arter*) AND (atherosclero* OR atherom* OR plaque* OR stent* OR stenos* OR narrow* OR restenos* OR endarterectom* OR desobstruct* OR des NEXT/1 obstruct* OR endovascular* OR occlusion*))):ti) AND ((((cognitive* OR neurocognitive* OR neurodegenerat* OR neuro NEXT/1 degenerat*) NEAR/3 (decline* OR impair* OR defect* OR complaint* OR difficult* OR disorder* OR problem* OR dysfunct* OR deficien*)) OR dement* OR Alzheimer* OR Binswanger NEXT/1 encephalopath OR CADASIL OR Lewy NEXT/1 body OR diffuse NEXT/1 neurofibrillary NEXT/1 tangles NEXT/1 with NEXT/1 calcification* OR hereditary NEXT/1 diffuse NEXT/1 leukoencephalopath* OR Huntington NEXT/1 chorea OR Kluver NEXT/1 Bucy OR mental NEXT/1 deteriorat OR Nasu NEXT/1 Hakola OR neuronal NEXT/1 ceroid NEXT/1 lipofuscinosis OR prion NEXT/1 disease OR pseudodementi OR Rett NEXT/1 syndrom OR tauopath* OR ((cognit* OR neurocognit*) NEAR/3 (function* OR change* OR performan* OR result*))):ab,ti,kw OR (((cognit* OR neurocognit*) NEAR/3 (outcome*))):ti)

Google Scholar

carotid atherosclerosis|stenosis dementia|dement|Alzheimer|'cognitive decline|impairment|defect|complaint|disorder|problem|dysfunction|deficiency| impairments|defects|complaints|disorders|problems|dysfunctions|deficiencies'

**Table S1: PRISMA checklist**

| Section and Topic | Item # | Checklist item | Location where item is reported |
|---|---|---|---|
| **TITLE** | | | |
| Title | 1 | Identify the report as a systematic review. | p1 |
| **ABSTRACT** | | | |
| Abstract | 2 | See the PRISMA 2020 for Abstracts checklist. | p2 |
| **INTRODUCTION** | | | |
| Rationale | 3 | Describe the rationale for the review in the context of existing knowledge. | p4 |
| Objectives | 4 | Provide an explicit statement of the objective(s) or question(s) the review addresses. | p4 |
| **METHODS** | | | |
| Eligibility criteria | 5 | Specify the inclusion and exclusion criteria for the review and how studies were grouped for the syntheses. | p5 |
| Information sources | 6 | Specify all databases, registers, websites, organisations, reference lists and other sources searched or consulted to identify studies. Specify the date when each source was last searched or consulted. | p5 |
| Search strategy | 7 | Present the full search strategies for all databases, registers and websites, including any filters and limits used. | Methods S1 |
| Selection process | 8 | Specify the methods used to decide whether a study met the inclusion criteria of the review, including how many reviewers screened each record and each report retrieved, whether they worked independently, and if applicable, details of automation tools used in the process. | p5 |
| Data collection process | 9 | Specify the methods used to collect data from reports, including how many reviewers collected data from each report, whether they worked independently, any processes for obtaining or confirming data from study investigators, and if applicable, details of automation tools used in the process. | p5-6 |
| Data items | 10a | List and define all outcomes for which data were sought. Specify whether all results that were compatible with each outcome domain in each study were sought (e.g. for all measures, time points, analyses), and if not, the methods used to decide which results to collect. | p5-6 |
| | 10b | List and define all other variables for which data were sought (e.g. participant and intervention characteristics, funding sources). Describe any assumptions made about any missing or unclear information. | p5-6 |
| Study risk of bias assessment | 11 | Specify the methods used to assess risk of bias in the included studies, including details of the tool(s) used, how many reviewers assessed each study and whether they worked independently, and if applicable, details of automation tools used in the process. | p6 |
| Effect measures | 12 | Specify for each outcome the effect measure(s) (e.g. risk ratio, mean difference) used in the synthesis or presentation of results. | p6 |
| Synthesis methods | 13a | Describe the processes used to decide which studies were eligible for each synthesis (e.g. tabulating the study intervention characteristics and comparing against the planned groups for each synthesis (item #5)). | P7 |
| | 13b | Describe any methods required to prepare the data for presentation or synthesis, such as handling of missing summary statistics, or data conversions. | p6 |
| | 13c | Describe any methods used to tabulate or visually display results of individual studies and syntheses. | p6-7 |

| Section and Topic | Item # | Checklist item | Location where item is reported |
|---|---|---|---|
| | 13d | Describe any methods used to synthesize results and provide a rationale for the choice(s). If meta-analysis was performed, describe the model(s), method(s) to identify the presence and extent of statistical heterogeneity, and software package(s) used. | p6-7 |
| | 13e | Describe any methods used to explore possible causes of heterogeneity among study results (e.g. subgroup analysis, meta-regression). | p7 |
| | 13f | Describe any sensitivity analyses conducted to assess robustness of the synthesized results. | p7 |
| Reporting bias assessment | 14 | Describe any methods used to assess risk of bias due to missing results in a synthesis (arising from reporting biases). | p7 |
| Certainty assessment | 15 | Describe any methods used to assess certainty (or confidence) in the body of evidence for an outcome. | - |
| **RESULTS** | | | |
| Study selection | 16a | Describe the results of the search and selection process, from the number of records identified in the search to the number of studies included in the review, ideally using a flow diagram. | p7-8, Fig 1 |
| | 16b | Cite studies that might appear to meet the inclusion criteria, but which were excluded, and explain why they were excluded. | p7-8 |
| Study characteristics | 17 | Cite each included study and present its characteristics. | p8, Tables 1-2 |
| Risk of bias in studies | 18 | Present assessments of risk of bias for each included study. | Tables S2-S4 |
| Results of individual studies | 19 | For all outcomes, present, for each study: (a) summary statistics for each group (where appropriate) and (b) an effect estimate and its precision (e.g. confidence/credible interval), ideally using structured tables or plots. | Tables 1-3 & S5-S6, Fig 2-3 & S1-S7 |
| Results of syntheses | 20a | For each synthesis, briefly summarise the characteristics and risk of bias among contributing studies. | p7-11 |
| | 20b | Present results of all statistical syntheses conducted. If meta-analysis was done, present for each the summary estimate and its precision (e.g. confidence/credible interval) and measures of statistical heterogeneity. If comparing groups, describe the direction of the effect. | p7-11, Fig 2-3 & S1-S7, Tables S5-S6 |
| | 20c | Present results of all investigations of possible causes of heterogeneity among study results. | p10-11, Fig 4 |
| | 20d | Present results of all sensitivity analyses conducted to assess the robustness of the synthesized results. | p10-11, Tables S7-S9 |
| Reporting biases | 21 | Present assessments of risk of bias due to missing results (arising from reporting biases) for each synthesis assessed. | p11, Table S10 |
| Certainty of | 22 | Present assessments of certainty (or confidence) in the body of evidence for each outcome assessed. | - |

| Section and Topic | Item # | Checklist item | Location where item is reported |
|---|---|---|---|
| evidence | | | |
| **DISCUSSION** | | | |
| Discussion | 23a | Provide a general interpretation of the results in the context of other evidence. | p11-12 |
| | 23b | Discuss any limitations of the evidence included in the review. | p13 |
| | 23c | Discuss any limitations of the review processes used. | p13 |
| | 23d | Discuss implications of the results for practice, policy, and future research. | p12-14 |
| **OTHER INFORMATION** | | | |
| Registration and protocol | 24a | Provide registration information for the review, including register name and registration number, or state that the review was not registered. | p5 |
| | 24b | Indicate where the review protocol can be accessed, or state that a protocol was not prepared. | p5 |
| | 24c | Describe and explain any amendments to information provided at registration or in the protocol. | |
| Support | 25 | Describe sources of financial or non-financial support for the review, and the role of the funders or sponsors in the review. | p14 |
| Competing interests | 26 | Declare any competing interests of review authors. | p14 |
| Availability of data, code and other materials | 27 | Report which of the following are publicly available and where they can be found: template data collection forms; data extracted from included studies; data used for all analyses; analytic code; any other materials used in the review. | p16 |

902

**Table S2: Neuropsychological tests used to assess individual cognitive domains in the included studies**

| Cognitive domain | Neuropsychological tests |
|---|---|
| Attention | Wechsler Adult Intelligence Scale-digital span (WAIS-DS); Digit Span: total, forward, backward; Digit Symbol; Symbol Digit Modalities Test (SDMT); MMSE attention; MoCA attention; Concentration (AKT); Seashore Rhythm; Number Vigilance; Sequential Number-Letter Trail Making Test |
| Memory | Auditory verbal learning test (AVLT): AVLT-1, AVLT-5, AVLT-short delay, AVLT-long delay, AVLT-recognition; Word learning (WL): total score, delayed score; Doors Test A/B; Letter number span (LNS); Fuld Object Memory Evaluation (FOME); Rapid verbal retrieval (RVR); Visual gestalts: learning, reproduction; Word pairs: learning, reproduction; Story recall I/II; Wechsler Memory Scale (WMS); Rey recall; Figure test recall: immediate recall, delay recall; MoCA: memory, orientation; MMSE: memory, orientation; Tactual Performance Test (TPT): memory; location; 15 Words Test: immediate recall, delayed recall; 12 word test; Recurring Faces Test1; |
| Processing speed | Performance IC; Number Connection Test; Trail Making Test A; Color Speed; Color Trails Test A; |
| Executive functioning | Trail Making Test B; Design Fluency; Stroop Test (Interference); Frontal Assessment Battery (FAB); MoCA: abstraction; Reasoning; Color Trails Test B; Choice; Four-choice reaction time task: decision time, movement time; Dual reaction time task: decision time, movement time, reaction time; |
| Language | Verbal fluency test: letters, categories; MMSE naming; MoCA: naming, sentence repeating; Boston naming test; Fluency: animals, professions; Wechsler Adult Intelligence Scale–Revised (WAIS-R): Verbal IQ; Word speed; Speech sounds |
| Motor speed | Finger tapping test; Motor planning; Motor dexterity; Manipulation test; Luria loop test |
| Visuospatial | Word Recognition; Picture recognition; Reading; Construction; Visual Motor; Copy figure test (CFT); Rey Copy; Rey Complex Figure Copy; Copy cube; Line connection; MMSE recognition; MoCA recognition |

**Table S3: Methodological assessment observational cohort studies (Newcastle Ottawa Scale)**

| Author (year) | NOS Selection | | | | | NOS Comparability | | NOS Outcome | | | | NOS Total | AHRQ |
|---|---|---|---|---|---|---|---|---|---|---|---|---|---|
| | Q1 | Q2 | Q3 | Q4 | Points | Q1 | Points | Q1 | Q2 | Q3 | Points | Points | |
| Van Oijen (2007)[15] | a | a | a | a | 4 | ab | 2 | b | a | b | 3 | 9 | Good |
| Zheng (2007)[16] | b | a | a | a | 4 | b | 1 | e | a | b | 3 | 8 | Good |
| Barnes (2010)[17] | b | a | a | a | 4 | ab | 2 | a | a | a | 3 | 9 | Good |
| Li (2012)[18] | c | a | b | a | 3 | c | 0 | e | a | b | 3 | 6 | Poor |
| Viticchi (2012)[19] | b | a | a | a | 4 | ab | 2 | e | a | a | 3 | 9 | Good |
| Wendell (2012)[20] | b | a | a | a | 4 | ab | 2 | a | a | a | 3 | 9 | Good |
| Viticchi (2014)[21] | b | a | a | a | 4 | ab | 2 | e | a | a | 3 | 9 | Good |
| Buratti (2015)[22] | c | a | a | a | 3 | c | 0 | a | a | a | 3 | 6 | Poor |
| Carcaillon (2015)[23] | b | a | a | a | 4 | ab | 2 | b | a | b | 3 | 9 | Good |
| Kitagawa (2015)[24] | c | a | a | a | 3 | ab | 2 | b | a | b | 3 | 8 | Good |
| Moon (2015)[25] | b | a | a | b | 3 | a | 1 | e | a | c | 2 | 6 | Good |
| Kuller (2016)[26] | c | a | a | a | 3 | a | 1 | e | a | c | 2 | 6 | Good |
| Romero-Sevilla (2018)[27] | c | a | a | a | 3 | b | 1 | a | a | b | 3 | 7 | Good |
| Rouch (2018)[28] | c | a | a | a | 3 | ab | 2 | b | a | b | 3 | 8 | Good |
| Crespo-Cuevas (2020)[29] | b | a | a | a | 4 | ab | 2 | b | a | a | 3 | 9 | Good |
| Gustavsson (2020)[30] | b | a | a | b | 3 | ab | 2 | b | a | b | 3 | 8 | Good |
| Wang, W. (2021)[31] | b | a | a | a | 4 | ab | 2 | b | a | b | 3 | 9 | Good |
| Liu (2024)[32] | b | a | a | a | 4 | a | 1 | e | a | a | 3 | 8 | Good |
| Box (2026) | a | a | a | a | 4 | ab | 2 | b | a | b | 3 | 9 | Good |
| Knopman (2001)[33] | b | a | a | a | 4 | a | 1 | e | a | c | 2 | 7 | Good |
| Johnston (2004)[34] | b | a | a | a | 4 | ab | 2 | e | a | b | 3 | 9 | Good |
| Talelli (2004)[35] | c | a | a | a | 3 | ab | 2 | a | a | b | 3 | 8 | Good |
| Komulainen (2007)[36] | c | a | a | a | 3 | ab | 2 | e | a | c | 2 | 7 | Good |
| Lee (2007)[37] | c | a | a | a | 3 | c | 0 | e | b | a | 2 | 5 | Poor |
| Voshaar (2007)[38] | b | a | a | a | 4 | ab | 2 | e | a | c | 2 | 8 | Good |
| Wendell (2009)[39] | b | a | a | a | 4 | ab | 2 | e | a | c | 2 | 8 | Good |
| Sander (2010)[40] | a | a | a | a | 4 | ab | 2 | b | a | b | 3 | 9 | Good |
| Arntzen (2012)[41] | a | a | a | b | 3 | ab | 2 | e | a | c | 2 | 7 | Good |
| Zhong (2012)[42] | b | a | a | a | 4 | a | 1 | e | a | c | 2 | 7 | Good |
| Balestrini (2013)[43] | c | c | a | a | 2 | c | 0 | a | a | a | 3 | 5 | Poor |
| Feinkohl (2013)[44] | c | a | a | a | 3 | a | 1 | e | a | b | 3 | 7 | Good |
| Frazier (2014)[45] | b | a | a | a | 4 | a | 1 | e | a | b | 3 | 8 | Good |
| Kawasaki (2016)[46] | b | a | a | a | 4 | a | 1 | e | a | c | 2 | 7 | Good |
| Wendell (2016)[47] | b | a | a | a | 4 | ab | 2 | e | a | b | 3 | 9 | Good |
| Gardener (2017)[48] | b | a | a | a | 4 | ab | 2 | e | a | c | 2 | 8 | Good |
| Wardlaw (2017)[49] | b | a | a | a | 4 | ab | 2 | e | a | c | 2 | 8 | Good |
| DelBrutto (2020)[50] | b | a | a | a | 4 | ab | 2 | e | a | a | 3 | 9 | Good |
| Wang, Y. (2021)[51] | c | a | a | a | 3 | a | 1 | e | a | c | 2 | 6 | Good |
| Lin, F. (2022)[52] | b | a | a | a | 4 | ab | 2 | a | a | d | 2 | 8 | Good |
| Ferreira (2023)[53] | b | a | a | a | 4 | ab | 2 | e | a | b | 3 | 9 | Good |
| Ghaznawi (2023)[54] | b | a | a | a | 4 | ab | 2 | e | a | c | 2 | 8 | Good |
| Liang (2024)[55] | a | a | a | a | 4 | ab | 2 | e | a | c | 2 | 8 | Good |
| Oestreich (2024)[56] | c | b | a | a | 2 | a | 1 | a | a | c | 2 | 5 | Fair |

Legend: NOS = Newcastle-Ottawa Scale; AHRQ = Agency for Healthcare Research and Quality assessment; Q = question. NOS selection Q1 a/b = 1 point, Q2 a = 1 point, Q3 a/b = 1 point, Q4 a = 1 point; NOS comparability ab = 2 point, a/b = 1 point; NOS outcome Q1 a/b/e = 1 point (e = assessment method described but blindness not mentioned), Q2 a = 1 point, Q3 a/b = 1 point. AHRQ: Good = 3 or 4 points in selection domain AND 1 or 2 stars in comparability domain AND 2 or 3 points in outcome domain; Fair = 2 points in selection domain AND 1 or 2 points in comparability domain AND 2 or 3 points in outcome/exposure domain; Poor = 0 or 1 points in selection domain OR 0 points in comparability domain OR 0 or 1 points in outcome domain.

**Table S4: Methodological assessment interventional studies (ROBINS-I-V2)**

| Author (year) | Preliminary: Decide whether to proceed with a risk-of-bias assessment | | | Risk of bias |
|---|---|---|---|---|
| | B1 | B2 | B3 | |
| King (1977)[65] | N | Y | | Critical |
| Matarazzo (1979)[66] | N | Y | | Critical |
| Van den Burg (1985)[67] | N | Y | | Critical |
| Hemmingsen (1986)[68] | N | Y | | Critical |
| Heyer (2002)[69] | N | Y | | Critical |
| Aharon-Peretz (2003)[70] | N | Y | | Critical |
| Fearn (2003)[71] | N | Y | | Critical |
| Bossema (2005)[72] | N | Y | | Critical |
| Lehrner (2005)[73] | N | Y | | Critical |
| Aleksic (2006)[74] | N | Y | | Critical |
| Bossema (2007)[75] | N | Y | | Critical |
| Xu (2007)[76] | N | Y | | Critical |
| Jansen (2008)[77] | N | Y | | Critical |
| Soinne (2009)[78] | N | Y | | Critical |
| Czerny (2010)[79] | N | Y | | Critical |
| Grunwald (2010)[80] | N | Y | | Critical |
| Huang (2011)[81] | N | Y | | Critical |
| Baracchini (2012)[82] | N | Y | | Critical |
| Yakhno (2012)[83] | N | Y | | Critical |
| Cheng (2013)[84] | N | Y | | Critical |
| Fan (2014)[85] | N | Y | | Critical |
| Shi (2014)[86] | N | Y | | Critical |
| Carta (2015)[87] | N | Y | | Critical |
| Plessers (2015)[88] | N | Y | | Critical |
| Qu (2015)[89] | N | Y | | Critical |
| Wapp (2015)[90] | N | Y | | Critical |
| Yoon (2015)[91] | N | Y | | Critical |
| Zhao (2015)[93] | N | Y | | Critical |

| | | | | |
|---|---|---|---|---|
| Kojima (2016)[94] | N | Y | | Critical |
| Shi, G.M. (2016)[95] | N | Y | | Critical |
| Yue (2016)[92] | N | Y | | Critical |
| Zhang (2016)[96] | N | Y | | Critical |
| Lu (2017)[97] | N | Y | | Critical |
| Watanabe (2017)[98] | N | Y | | Critical |
| Belov (2018)[99] | N | Y | | Critical |
| Pucite (2019)[100] | N | Y | | Critical |
| Song (2019)[101] | N | Y | | Critical |
| Thirumala (2020)[102] | N | Y | | Critical |
| Whooley (2020)[103] | N | Y | | Critical |
| Matyushkin (2021)[104] | N | Y | | Critical |
| Turowicz (2021)[105] | N | Y | | Critical |
| Huang (2021)[106] | N | Y | | Critical |
| Lin, C.J. (2022)[107] | N | Y | | Critical |
| Relander (2022)[108] | Y | NA | PN | Critical |
| Tang (2023)[109] | N | Y | | Critical |
| Xiao (2023)[110] | N | Y | | Critical |
| Bernstein (2024)[111] | N | Y | | Critical |
| Relander (2024)[112] | Y | NA | PN | Critical |

Legend: ROBINS-I-V2= Risk of Bias in Non-randomized Studies of Interventions version 2; N= no; Y= yes; NA= not applicable; PN = probably no. B1: Did the authors make any attempt to control for confounding in the result being assessed? B2: If N/PN to B1: Is there sufficient potential for confounding that this result should not be considered further? B3: Was the method of measuring the outcome inappropriate? According to ROBINS-I-V2 guidance, if the answer to either B2 or B3 is 'Yes' or 'Probably yes', the result should be considered to be at 'Critical risk of bias' and no further assessment is required. Relander (2022)[108] and Relander (2024)[112] adjusted for age, sex, and education level; however, after a complete ROBINS-I-V2 domain assessment, the overall risk bias remained critical.

**Table S5: Methodological assessment randomized control studies RoB2**

| Author (year) | Domain 1: Randomization process | | | | Domain 2: Deviations from intended interventions | | | | | | | | Domain 3: Missing outcome data | | | | | Domain 4: Measurement of the outcome | | | | | | Domain 5: Selection of the reported result | | | | Overall bias |
|---|---|---|---|---|---|---|---|---|---|---|---|---|---|---|---|---|---|---|---|---|---|---|---|---|---|---|---|---|
| | 1.1 | 1.2 | 1.3 | Risk of bias | 2.1 | 2.2 | 2.3 | 2.4 | 2.5 | 2.6 | 2.7 | Risk of bias | 3.1 | 3.2 | 3.3 | 3.4 | Risk of bias | 4.1 | 4.2 | 4.3 | 4.4 | 4.5 | Risk of bias | 5.1 | 5.2 | 5.3 | Risk of bias | |
| Pettigrew (2000)[58] | Y | NI | Y | low | NI | NI | PN | NA | NA | Y | NA | Some concerns | PY | NA | NA | NA | low | N | N | NI | PN | NI | low | NI | NI | NI | Some concerns | Some concerns |
| Marshall (2014)[59] | Y | NI | N | Some concerns | Y | Y | NI | NA | NA | PY | NA | Some concerns | PN | N | Y | PY | high | N | N | N | NA | NA | low | PN | PN | NI | Some concerns | high |
| Shi, H. (2016)[60] | Y | NI | N | Some concerns | Y | Y | PN | NA | NA | PN | PN | Some concerns | PN | N | Y | PY | high | N | PN | NI | PY | NI | high | NI | PN | PN | Some concerns | high |
| Gao (2022)[61] | PY | NI | N | Some concerns | Y | Y | NI | NA | NA | Y | NA | Some concerns | Y | NA | NA | NA | low | N | N | N | NA | NA | low | NI | NI | NI | Some concerns | Some concerns |
| Halliday (2022)[62] | Y | PY | Y | Some concerns | NI | NI | PN | NA | NA | Y | NA | Some concerns | Y | NA | NA | NA | Some concerns * | N | N | N | NA | NA | low | Y | N | N | low | Some concerns |
| Lazar (2026)[63] | Y | PY | N | low | Y | Y | PN | NA | NA | Y | NA | low | N | N | PY | PY | high | N | N | N | NA | NA | low | PY | N | N | low | Some concerns * * |

Legend: RoB2 = Cochrane risk of bias for randomized trails. Y = yes; PY = probably yes; PN = probably no; N = NO; NI = no information; NA = not applicable; * According to the standard RoB 2 algorithm, the outcome assessment domain would be judged at low risk of bias. However, as dementia outcomes were ascertained through long-term registry linkage, which may have introduced differential outcome ascertainment between trial arms, we overruled the standard RoB 2 ruling and judged this domain to have some concerns of bias. * * According to the official RoB2 scoring this would be high risk of bias to reflect the lowest score, however since all domains except domain 3 is judged at low risk we overruled the RoB2 standard ruling and judge this study to contain some concerns of bias instead of high risk.

### Table S6: Association of carotid atherosclerosis with specific cognitive domains

| Study (year) | N | Fup-time | Exposure | Cognitive test | Analysis | Effect estimate [95%CI] | AHQR | Conclusion |
|---|---|---|---|---|---|---|---|---|
| **Memory as outcome** | | | | | | | | |
| **IMT as exposure** | | | | | | | | |
| Knopman (2001)[33] | 10882 | 6y | IMT (tertiles) | Delayed word recall test | Linear regression | aβ not provided but all were not statistically significant according to text. | Good | Higher IMT is not significantly associated with a decrease in memory. |
| Wendell (2009)[39] | 437 | 4y | IMT (continuous) | California Learning test -<br>1: learning slope<br>2: immediate free recall<br>3: short delay free recall<br>4: long delay free recall<br>5: Benton Visual Retention Test<br>6: Rey Complex Figure Test – immediate recall<br>7: Delayed recall | Linear mixed model | aβ [SE]<br>1: -0.02 [0.01], p >0.05<br>2: -0.65 [0.29], p <0.05<br>3: -0.14 [0.08], p >0.05<br>4: -0.19 [0.08], p <0.05<br>5: 0.17 [0.09], p <0.05<br>6: -1.30 [0.58], p <0.05<br>7: -1.32 [0.55], p <0.05 | Good | Higher IMT is significantly associated with faster decline in memory in 5 of 7 memory tests. |
| Zhong (2012)[42] | 1143 | 10y | IMT (per 0.1mm) | Rey Auditory Verbal Learning Test | Linear regression | aβ [SE]: 0.03 [ 0.11], p = 0.82 | Good | Higher IMT is not associated with a significant decrease in memory. |
| Frazier (2014)[45] | 251 | 2.3y | IMT (mean of left and right, quartiles) | Recall test | Linear mixed model | aβ [SE]<br>stepwise per quantile increase: 1.89 [1.19,] p >0.05<br>2nd quartile: 2.71 [1.34], p >0.05<br>3rd quartile: 1.56 [1.18], p >0.05<br>4th quartile: -0.54 [1.15], p >0.05 | Good | Higher IMT is not significantly associated with faster decline in memory. |
| Gardener (2017)[48] | 836 | 5y | IMT (per mm, in area without plaque) | 1: Composite for episodic memory<br>2: composite for semantic memory | Linear regression | aβ [95%CI]<br>1: 0.869 [np], p = 0.039<br>2: 0.244 [np], p = 0.569 | Good | Higher IMT in areas without plaque is associated with a decrease in episodic but not semantic memory. |
| Lin, F. (2022)[52] | 1495 | 2.5y | IMT (per 0.1 mm) | 1: Composite for verbal memory<br>2: Composite for visual memory | Linear regression | aβ [95%CI]:<br>1: -0.0236 [-0.079; 0.032], p = 0.41<br>2: -0.0231 [-0.064; 0.018], p = 0.27 | Good | Higher IMT is not significantly associated with memory over time. |
| Ferreira (2023)[53] | 9263 | 8y | IMT (per mm) | Word Learning Task | Linear mixed models with random intercepts and slopes | aβ [95%CI]: −0.018 [−0.026; −0.011] p = <0.001 | Good | Higher IMT is associated with faster decline in memory. |

| | | | | | | | | |
|---|---|---|---|---|---|---|---|---|
| Liang (2024)[55] | 932 | 3y | IMT (per 0.1mm and tertiles) | Immediate and delayed recall | Linear mixed models | aβ [95%CI]<br>per 0.1mm: -0.011 [-0.031; 0.010], p = 0.322<br>2nd tertile: -0.025 [-0.082; 0.033], p = 0.398<br>3rd tertile: -0.043 [-0.101; 0.014], p = 0.142 | Good | Higher IMT is not significantly associated with faster decline in memory. |
| **Plaque as exposure** | | | | | | | | |
| Gardener (2017)[48] | 836 | 5y | 1: Plaque (dichotomous)<br>2: Plaque area (continuous) | 1: Composite for episodic memory<br>2: composite for semantic memory | Linear regression | aβ [SE]<br>1.1: 0.085 [np], p = 0.233<br>1.2: 0.062 [np], p = 0.399<br>2.1: 0.001 [np], p = 0.761<br>2.2: 0.002 [np], p = 0.292 | Good | Plaque is associated with memory. |
| Arntzen (2012)[41] | 5939 | 13y | 1: Plaque count<br>2: Plaque area (right sided) | 12 word memory test | Linear regression | aβ [95%CI]:<br>1: 0.03 [-0.02; 0.08], p = 0.22<br>2: 0.03 [-0.02; 0.08], p = 0.23 | Good | Plaque is not significantly associated with memory. |
| Zhong (2012)[42] | 1311 | 10y | Plaque (categorized: none, 1-3, 4-6) | Rey Auditory Verbal Learning Test | Linear regression | aβ [SE]<br>1-3 vs 0: 0.27 [0.44], p = 0.54<br>4-6 vs 0: 0.92 [0.71], p = 0.20 | Good | Plaque is not significantly associated with memory over time. |
| Liang (2024)[55] | 932 | 3y | No. of plaque (continuous and categorical) | Immediate and delayed recall | Linear mixed models | aβ [95%CI]<br>per 0.1mm: -0.004 [-0.011; 0.019], p = 0.568<br>2nd tertile: -0.003 [-0.062; 0.057], p = 0.929<br>3rd tertile: 0.031 [-0.041; 0.103], p = 0.40 | Good | Plaque is not significantly associated with a decline in memory. |
| **Stenosis as exposure** | | | | | | | | |
| Wardlaw (2017)[49] | 697 | 6y | Stenosis (in steps of 10%) | Composite | Latent growth model (slope for decline) | aβ [SE]: -0.238 [0.187], p = 0.203 | Good | Stenosis is not significantly associated with a decline in memory. |
| Wang, Y. (2021)[51] | 128 | 6.9m | Symptomatic stenosis (unclear how this is defined) | Composite | Linear regression | aβ [SE] 0.08 [0.15], p >0.05 | Good | Symptomatic stenosis is not significantly associated with memory in ischemic stroke patients. |
| Ghaznawi (2023)[54] | 510 | 4y | Asymptomatic stenosis 1-49% | Composite of 15 word learning test and Rey Ostterrieth Complex Figure test | Linear mixed model | aβ [95%CI] : -0.012 [-0.02; -0.001], p = 0.032 | Good | Mild asymptomatic stenosis is associated with faster decline in memory in patients referred for treatment of other atherosclerotic disease. |
| Oestreich (2024)[56] | 141 | 1y | Symptomatic stenosis > 50% | 1: delayed recall<br>2: total recall<br>3: digit span forward<br>4: digit span backward | Logistic regression | aOR [95CI]:<br>1. 1.17 [np], p = 0.82<br>2. 1.45 [np], p = 0.84<br>3. 2.17 [np], p = 0.46<br>4. 9.23 [np], p = 0.017 | Fair | In ischemic stroke patients, symptomatic stenosis >50% is not significantly associated with decline in memory, though one of four tests was significant. |

| | | | | | | | | |
|---|---|---|---|---|---|---|---|---|
| Box (2026)[57] | 4181 | 7.5y | Asymptomatic stenosis<br>1. 1-49% vs 0%<br>2. 50-99% vs 0%<br>3. 100% vs 0% | Word learning test – delayed recall | Linear mixed models | aβ [95%CI]:<br>1: 0.004 [-0.004; 0.012], p = 0.34<br>2: 0.016 [-0.009; 0.040], p = 0.22<br>3: 0.027 [-0.073; 0.127], p = 0.60 | Good | Asymptomatic stenosis is not associated with faster decline in executive functioning. |
| **Executive functioning as outcome** | | | | | | | | |
| **IMT as exposure** | | | | | | | | |
| Wendell (2009)[39] | 257 | 4y | IMT (continuous) | 1: TMT A<br>2: TMT B | Linear mixed model | aβ [SE]<br>1: 0.76 [0.72], p >0.05<br>2: 0.02 [0.01], p >0.05 | Good | Higher IMT is not significantly associated with faster decline in executive functioning. |
| Zhong (2012)[42] | 1185 | 10y | IMT (per 0.1mm) | 1: TMT A<br>2: TMT B | Linear regression | aβ [SE]<br>1: 0.74 [0.46], p = 0.11<br>2: 2.27 [0.98], p = 0.02 | Good | Mixed results, higher IMT might be associated with executive functioning over time. |
| Frazier (2014)[45] | 251 | 2.3y | IMT (mean of left and right, quartiles) | Composite | Linear mixed model | stepwise per quantile increase: 0.016 [0.77], p >0.05<br>2nd quartile: -0.007 [0.92], p >0.05<br>3rd quartile: 1.46 [0.83], p >0.05<br>4th quartile: -2.95 [0.77], p <0.01 | Good | Mixed results, higher IMT does not seem to be significantly associated with faster decline in executive functioning, though those with highest quartile of IMT did decline significantly faster. |
| Gardener (2017)[48] | 836 | 5y | IMT (per mm, in area without plaque) | Color trail test + odd-man-out subtest | Linear regression | aβ [95%CI]: -0.47 [np], p = 0.27 | Good | Higher IMT in areas without plaque is not associated with executive functioning. |
| Lin, F. (2022)[52] | 1495 | 2.5y | IMT (per 0.1 mm) | Composite | Linear regression | aβ [95%CI]: -0.0179 [-0.055; 0.019], p = 0.34 | Good | Higher IMT is not significantly associated with executive functioning over time. |
| Ferreira (2023)[53] | 9263 | 8y | IMT (per mm) | TMT B | Linear mixed models with random intercepts and slopes | aβ [95%CI]: −0.022 [−0.030; −0.013], p <0.001 | Good | Higher IMT is associated with faster decline in executive functioning. |
| Liang (2024)[55] | 932 | 3y | IMT (per 0.1mm and tertiles) | TMT A + TMT B | Linear mixed models | aβ [95%CI]<br>per 0.1mm: -0.006 [-0.020; 0.008], p = 0.396<br>2nd tertile: 0.016 [-0.022; 0.054], p = 0.416<br>3rd tertile: -0.02 [-0.059; 0.018], p = 0.305 | Good | Higher IMT is not significantly associated with faster decline in executive functioning. |
| **Plaque as exposure** | | | | | | | | |
| Gardener (2017)[48] | 836 | 5y | 1: Plaque (dichotomous) | Composite | Linear regression | aβ [SE]<br>1: 0.012 [np], p = 0.873<br>1: -0.0003 [np], p = 0.886 | Good | Plaque is not significantly associated with executive functioning. |

| | | | | | | | | |
|---|---|---|---|---|---|---|---|---|
| | | | 2: Plaque area (continuous) | | | | | |
| Zhong (2012)[42] | 1134 | 10y | Plaque (categorized: none, 1-3, 4-6) | 1: TMT A<br>2: TMT B | Linear regression | aβ [SE]<br>1.<br>1-3 vs 0: -1.03 [1.86], p = 0.58<br>4-6 vs 0: 4.67 [3.01], p = 0.12<br>2.<br>1-3 vs 0: 0.88 [3.84], p = 0.82<br>4-6 vs 0: 7.81 [6.31], p = 0.22 | Good | Plaque is not significantly associated with executive functioning over time. |
| **Stenosis as exposure** | | | | | | | | |
| Wang, Y. (2021)[51] | 128 | 6.9m | Symptomatic stenosis (unclear how this is defined) | Executive screening scale | Linear regression | aβ [SE]: -0.16 [0.14], p >0.05 | Good | Symptomatic stenosis is not significantly associated with executive functioning in ischemic stroke patients. |
| Ghaznawi (2023)[54] | 510 | 4y | Asymptomatic stenosis 1-49% | Composite of 15 word learning test and Rey Ostterrieth Complex Figure test | Linear mixed model | aβ [95%CI] : -0.02 [-0.031; -0.01], p = 0.032 | Good | Mild asymptomatic stenosis is associated with faster decline in executive functioning in patients referred for treatment of other atherosclerotic disease. |
| Box (2026)[57] | 4181 | 7.5y | Asymptomatic stenosis<br>1. 1-49% vs 0%<br>2. 50-99% vs 0%<br>3. 100% vs 0% | Stroop interference | Linear mixed models | aβ [95%CI]:<br>1: -0.005 [-0.015; 0.005], p = 0.29<br>2: -0.013 [-0.018; 0.044], p = 0.41<br>3: -0.005 [-0.118; 0.108], p = 0.93 | Good | Asymptomatic stenosis is not associated with faster decline in executive functioning. |
| **Language as outcome** | | | | | | | | |
| **IMT as exposure** | | | | | | | | |
| Knopman (2001)[33] | 10882 | 6y | IMT (tertiles) | Word fluency test | Linear regression | aβ not provided but all were not statistically significant according to text. | Good | Higher IMT is not significantly associated with language over time. |
| Wendell (2009)[39] | 257 | 4y | IMT (continuous) | 1: Letter fluency<br>2: Category fluency<br>3: Boston Naming Test | Linear mixed model | aβ [SE]<br>1: -0.10 [0.17], p >0.05<br>2: -0.37 [0.16], p <0.05<br>3: -0.13 [0.25], p >0.05 | Good | Mixed results, higher IMT is not significantly associated with faster decline in executive functioning in 2 of 3 tests, 1 test was significant. |
| Zhong (2012)[42] | 1199 | 10y | IMT (per 0.1mm) | Verbal fluency test | Linear regression | aβ [SE]: 0.11 [0.18], p = 0.56 | Good | Higher IMT was not significantly associated with language over time. |
| Ferreira (2023)[53] | 9263 | 8y | IMT (per mm) | Verbal fluency test | Linear mixed models with random intercepts and slopes | aβ [95%CI]: −0.016 [−0.023; −0.009], p <0.001 | Good | Higher IMT is associated with faster decline in language. |

| | | | | | | | | |
|---|---|---|---|---|---|---|---|---|
| Liang (2024)[55] | 932 | 3y | IMT (per 0.1mm and tertiles) | Category fluency test | Linear mixed models | aβ [95%CI]<br>per 0.1mm: -0.026 [-0.043; -0.009], p =0.003,<br>2nd tertile: -0.045 [-0.092; 0.003], p = 0.064,<br>3rd tertile: -0.065, -0.112; -0.017], p = 0.008 | Good | Higher IMT is associated with faster decline in language. |
| **Plaque as exposure** | | | | | | | | |
| Zhong (2012)[42] | 1203 | 10y | Plaque (categorized: none, 1-3, 4-6) | Verbal fluency test | Linear regression | aβ [SE]<br>1-3 vs 0: 1.02 [0.73], p = 0.16<br>4-6 vs 0: 0.04 [1.18], p = 0.98 | Good | Plaque is not significantly associated with language over time. |
| Liang (2024)[55] | 932 | 3y | Plaque (dichotomous) | Category fluency test | Linear mixed models | aβ [95%CI]<br>per 0.1mm: -0.016 [-0.28; -0.004], p = 0.01<br>2nd tertile: -0.013 [-0.062; 0.036], p = 0.593<br>3rd tertile: -0.07 [-0.130; -0.011], p = 0.021 | Good | Plaque is associated with faster decline in language. |
| **Stenosis as exposure** | | | | | | | | |
| Wang, Y. (2021)[51] | 128 | 6.9m | Symptomatic stenosis (unclear how this is defined) | Language screening scale | Linear regression | aβ [SE]: -0.16 [0.16], p >0.05 | Good | Symptomatic stenosis is not significantly associated with language in ischemic stroke patients. |
| Box (2026)[57] | 4181 | 7.5y | Asymptomatic stenosis<br>1. 1-49% vs 0%<br>2. 50-99% vs 0%<br>3. 100% vs 0% | Word fluency test | Linear mixed models | aβ [95%CI]:<br>1: -0.004 [-0.012; 0.004], p = 0.31<br>2: -0.007 [-0.032; 0.017], p = 0.57<br>3: 0.019 [-0.079; 0.118], p = 0.70 | Good | Asymptomatic stenosis is not significantly associated with faster decline in language. |
| **Processing speed as outcome** | | | | | | | | |
| **IMT as exposure** | | | | | | | | |
| Knopman (2001)[33] | 10882 | 6y | IMT (tertiles) | Word fluency test | Linear regression | aβ not provided but all were not statistically significant according to text. | Good | Higher IMT is not significantly associated with processing speed over time. |
| Zhong (2012)[42] | 1160 | 10y | IMT (per 0.1mm) | Digit symbol substitution test | Linear regression | aβ [SE]: -0.12 [0.17], p = 0.47 | Good | Higher IMT is not significantly associated with processing speed. |
| Gardener (2017)[48] | 836 | 5y | IMT (per mm, in area without plaque) | Composite | Linear regression | aβ [95%CI]: -0.157 [np], p = 0.97 | Good | Higher IMT in areas without plaque is not significantly associated with processing speed. |
| **Plaque as exposure** | | | | | | | | |
| Arntzen (2012)[41] | 5939 | 13y | 1: Plaque count<br>2: Plaque area (right sided) | 1. Digit symbol coding test<br>2. Tapping test | Linear regression | aβ [95%CI]: | Good | Plaque is not significantly associated with cognitive speed. |

| | | | | | | | | |
|---|---|---|---|---|---|---|---|---|
| | | | | | | 1.1: 0.02 [-0.03; 0.07], p = 0.51<br>1.2: -0.02 [-0.07; 0.03], p = 0.39<br>2.1: 0.02 [-0.03; 0.07], p = 0.46<br>2.2: -0.01 [-0.06; 0.04], p = 0.71 | | |
| Zhong (2012)[42] | 1140 | 10y | Plaque (categorized: none, 1-3, 4-6) | Digit symbol substitution test | Linear regression | aβ [SE]<br>1-3 vs 0: 0.18 [0.65], p = 0.78<br>4-6 vs 0: -0.50 [1.07], p = 0.64 | Good | Plaque is not significantly associated with processing speed over time. |
| Gardener (2017)[48] | 836 | 5y | 1: Plaque (dichotomous)<br>2: Plaque area (continuous) | Composite | Linear regression | aβ [SE]<br>1: 0.026 [np], p = 0.713<br>1: -0.001 [np], p = 0.375 | Good | Plaque is not significantly associated with processing speed. |
| **Stenosis as exposure** | | | | | | | | |
| Wardlaw (2017)[49] | 697 | 6y | Stenosis (in steps of 10%) | Symbol search + Digit symbol test | Latent growth model (slope for decline) | aβ [SE]: -0.114 [0.066], p = 0.08 | Good | Stenosis is not significantly associated with decline in processing speed. |
| Box (2026)[57] | 4181 | 7.5y | Asymptomatic stenosis<br>1. 1-49% vs 0%<br>2. 50-99% vs 0%<br>3. 100% vs 0% | 1. Letter digit substitution test<br>2. Purdue pegboard test | Linear mixed models | aβ [95%CI]:<br>1.1: -0.003 [-0.009; 0.003], p = 0.29<br>2.1: 0.001 [-0.018; 0.019], p = 0.93<br>3.1: 0.014 [-0.062; 0.091], p = 0.71<br><br>1.2: 0.006 [-0.000; 0.013], p = 0.06<br>2.2: 0.015 [-0.006; 0.035], p = 0.16<br>3.2: 0.083 [-0.001; 0.167], p = 0.052 | Good | Asymptomatic stenosis is not significantly associated with faster decline in processing speed. |
| **Visuo-spatial function as outcome** | | | | | | | | |
| **IMT as exposure** | | | | | | | | |
| Wendell (2009)[39] | 1: 435<br>2: 160 | 4y | IMT (continuous) | 1: Card rotation test<br>2: Rey complex figure test - copy | Linear mixed model | aβ [SE]<br>1: -1.19 [0.92], p >0.05<br>2: -0.23 [0.31], p >0.05 | Good | IMT not significantly associated with faster decline in visuo-spatial functioning. |
| **Stenosis as exposure** | | | | | | | | |
| Wardlaw (2017)[49] | 697 | 6y | Stenosis (in steps of 10%) | Composite | Latent growth model (slope for decline) | aβ [SE]: 0.05 [0.154], p = 0.74 | Good | Stenosis is not significantly associated with decline in visual spatial functioning. |
| Wang, Y. (2021)[51] | 128 | 6.9m | Symptomatic stenosis (unclear how this is defined) | Overlapping diagram of MoCA | Linear regression | aβ [SE]: -0.32 [0.14], p <0.05 | Good | Symptomatic stenosis is associated with worse visuo-spatial functioning in ischemic stroke patients. |
| **Attention as outcome** | | | | | | | | |
| Wendell (2009)[39] | 1: 438<br>2: 440 | 4y | IMT (continuous) | 1: Digit forward test<br>2: Digit backwards test | Linear mixed model | aβ [SE]<br>1: -0.04 [0.06], p >0.05<br>2: -0.02 [0.07], p >0.05 | Good | IMT is not significantly associated with faster decline in attention. |

Legend: N = number of participants in analysis; Fup = follow-up; y = year; m = months; CI = confidence interval; AHQR = Agency for Healthcare Research and Quality assessment; aβ = adjusted β-coefficient; HR = hazard ratio; aHR = adjusted hazard ratio; OR = odds ratio; aOR = adjusted odds ratio; np = not provided; SE = standard error; TMT = trail-making test.

### Table S7: Effect of CEA/CAS on cognition, individual study results

| Surgery type | Author (Year) | Neuropsychological tests | Reference group | n analysis (Total) | n analysis (INT) | n analysis (REF) | Δ Mean± SD (INT) | Δ Mean± SD (REF) | Effect size (Hedges' g) | Variance |
|---|---|---|---|---|---|---|---|---|---|---|
| CEA | Matarazzo (1979)[66] | Halstead Impairment Index Scores | Patients with diffuse cerebrovascular disease | 31 | 15 | 16 | 0.02±0.13 | 0.04±0.09 | -0.26 | 0.36 |
| | | Category Test Scores | | | | | -6.14±20.07 | 1.25±11.19 | | |
| | Van den Burg (1985)[67] | Groningen Intelligence Test | Patients with peripheral vascular surgery and normal control | 40 | 20 | 20 | 0±9.22 | 2.00±12.02 | -0.15 | 0.32 |
| | Fearn (2003)[71] | Overall (word, pic recognition, and memory scanning) | Patients undergoing urological surgery | 178 | 159 | 20 | -0.01±0.23 | -0.06±0.27 | 0.17 | 0.24 |
| | Baracchini (2012)[82] | MMSE | patients undergoing laparoscopic cholecystectomy for gallbladder disease | 213 | 145 | 68 | 0.31±0.62 | -0.1±1.03 | 0.45 | 0.15 |
| | | MoCA | | | | | 0.61±1.11 | 0±1.02 | | |
| | Carta (2015)[87] | Overall IC | patients without surgery* | 40 | 30 | 10 | 7.14±12.06 | 0.5±12.48 | 0.44 | 0.37 |
| | Qu (2015)[89] | MMSE | patients without surgery | 80 | 40 | 40 | 3.50±1.28 | 0.30±1.26 | 2.04 | 0.28 |
| | Kojima (2016)[94] | WAIS-R performance IQ | healthy volunteers | 161 | 116 | 45 | 6.70±4.20 | 4.40±3.00 | 0.83 | 0.18 |
| | Zhang (2016)[96] | MMSE | patients without surgery | 63 | 35 | 28 | 0.10±0.80 | 0.50±0.72 | -0.21 | 0.25 |
| | | CDT | | | | | -0.10±0.48 | -0.10±0.36 | | |
| CAS | Xu (2007)[76] | MMSE | patients without surgery | 104 | 47 | 57 | -0.40±2.50 | 0.40±2.42 | -0.27 | 0.20 |
| | Cheng (2013)[84] | MMSE | patients without surgery | 208 | 144 | 64 | 0.2±1.15 | -0.2±0.98 | 0.36 | 0.15 |
| | | MoCA | | | | | 0.4±1.20 | -0.2±1.08 | | |
| | Fan (2014)[85] | MoCA | patients without surgery | 40 | 22 | 18 | 10.05±2.15 | 4.19±2.05 | 2.27 | 0.41 |
| | Shi (2014)[86] | MoCA | patients without surgery | 43 | 31 | 12 | 2.50±1.82 | 0.30±1.69 | 0.99 | 0.36 |
| | Zhao (2015)[93] | MMSE | Patients with non-stent-assisted angioplasty | 122 | 78 | 44 | 6.49±3.30 | 0.09±1.93 | 1.94 | 0.23 |
| | Yue (2016)[92] | MMSE | | 105 | 83 | 22 | 1.50±3.04 | 0.10±1.82 | 0.42 | 0.24 |

| | | | | | | | | | | |
|---|---|---|---|---|---|---|---|---|---|---|
| | | MoCA | patients without surgery | | | | 2.10±3.94 | 0.10±2.54 | | |
| | Lu (2017)[97] | MMSE | patients without surgery | 30 | 15 | 15 | 1.50±1.68 | -0.60±2.00 | 0.73 | 0.38 |
| | | MoCA | | | | | 1.00±3.00 | -1.00±3.00 | | |
| | Song (2019)[101] | MoCA | patients without surgery | 79 | 48 | 31 | 3.67±1.68 | -1.88±1.40 | 3.03 | 0.33 |
| | Lin, C.J. (2022)[107] | MMSE | patients without surgery | 46 | 23 | 23 | 0.20±0.89 | 0.20±1.03 | 0.00 | 0.29 |
| | Xiao (2023)[110] | MoCA | patients without surgery | 33 | 12 | 21 | 1.60±1.44 | 0.20±1.72 | 0.69 | 0.37 |
| CEA/CAS | Shi, G.M. (2016)[95] | MMSE | patients without surgery | 154 | 77 | 77 | 0.75±0.78 | 0.08±0.62 | 1.12 | 0.17 |
| | | MoCA | | | | | 0.91±0.89 | 0.03±0.51 | | |
| | Pucite (2019)[100] | MoCA | patients without surgery | 213 | 188 | 25 | 1.25±2.30 | 1.00±1.99 | 0.09 | 0.21 |
| | Matyushkin (2021)[104] | MoCA | patients without surgery | 186 | 136 | 50 | 1.56±1.03 | 0.32±1.00 | 1.02 | 0.17 |
| HS | Tang (2023)[109] | MMSE | patients without surgery | 80 | 40 | 40 | 2.60±1.68 | 0.60±3.28 | 0.63 | 0.23 |

Legend: CEA = carotid endarterectomy; CAS = carotid artery stenting; HS= hybrid surgery; INT= intervention group; REF= reference group; MMSE = Mini-Mental State Examination; MoCA = Montreal Cognitive Assessment; Overall IC = Overall Intelligence Composite; WAIS-R performance IQ = Wechsler Adult Intelligence Scale–Revised Performance Intelligence Quotient; CDT = clock drawing test; 3MSE = Modified Mini-Mental State Examination; SD= standard deviation. *Patients from the same source population as the intervention group did not undergo carotid intervention.

**Table S8: Assumptions for effect size computation**

| r | SMD | Lower 95% CI | Upper 95% CI | P-value |
|---|---|---|---|---|
| **Global cognition** | | | | |
| 0.5 | 0.60 | 0.31 | 0.89 | < 0.001 |
| 0.7 | 0.73 | 0.37 | 1.09 | < 0.001 |
| 0.9 | 1.09 | 0.54 | 1.62 | < 0.001 |
| **Attention** | | | | |
| 0.5 | 0.03 | -0.21 | 0.27 | >0.05 |
| 0.7 | 0.02 | -0.27 | 0.31 | >0.05 |
| 0.9 | -0.03 | -0.47 | 0.41 | >0.05 |
| **Memory** | | | | |
| 0.5 | 0.15 | -0.13 | 0.43 | >0.05 |
| 0.7 | 0.16 | -0.18 | 0.51 | >0.05 |
| 0.9 | 0.20 | -0.31 | 0.71 | >0.05 |
| **Processing speed** | | | | |
| 0.5 | 0.06 | -0.15 | 0.27 | >0.05 |
| 0.7 | 0.17 | -0.16 | 0.51 | >0.05 |
| 0.9 | 0.30 | -0.21 | 0.81 | >0.05 |
| **Executive functioning** | | | | |
| 0.5 | 0.06 | -0.15 | 0.27 | >0.05 |
| 0.7 | 0.07 | -0.19 | 0.33 | >0.05 |
| 0.9 | 0.12 | -0.29 | 0.52 | >0.05 |
| **Language** | | | | |
| 0.5 | 0.13 | -0.12 | 0.39 | >0.05 |
| 0.7 | 0.14 | -0.15 | 0.44 | >0.05 |
| 0.9 | 0.18 | -0.25 | 0.62 | >0.05 |
| **Motor speed** | | | | |
| 0.5 | 0.14 | -0.27 | 0.55 | >0.05 |
| 0.7 | 0.15 | -0.28 | 0.58 | >0.05 |
| 0.9 | 0.21 | -0.41 | 0.82 | >0.05 |
| **Visuospatial** | | | | |
| 0.5 | 0.30 | 0.01 | 0.59 | 0.04 |
| 0.7 | 0.35 | 0.06 | 0.64 | 0.02 |
| 0.9 | 0.50 | 0.19 | 0.85 | 0.01 |

Legend: r = the assumed correlation coefficient between the two repeated measurements used to estimate the standard deviation of the change score difference (SD_diff); SMD = Hedge’s standardized mean difference; *p*: the statistical significance of the pooled effect.

**Table S9: Variations in the RVE model correlation parameter**

| ρ | SMD | Lower 95% CI | Upper 95% CI | P-value |
|---|---|---|---|---|
| **Global cognition** | | | | |
| 0 | 0.73 | 0.35 | 1.10 | <0.001 |
| 0.2 | 0.73 | 0.35 | 1.10 | <0.001 |
| 0.4 | 0.73 | 0.35 | 1.10 | <0.001 |
| 0.6 | 0.73 | 0.35 | 1.10 | <0.001 |
| 0.8 | 0.73 | 0.35 | 1.10 | <0.001 |
| 1.0 | 0.73 | 0.35 | 1.10 | <0.001 |
| **Attention** | | | | |
| 0 | 0.03 | -0.31 | 0.36 | >0.05 |
| 0.2 | 0.03 | -0.31 | 0.36 | >0.05 |
| 0.4 | 0.03 | -0.31 | 0.36 | >0.05 |
| 0.6 | 0.03 | -0.31 | 0.36 | >0.05 |
| 0.8 | 0.03 | -0.31 | 0.36 | >0.05 |
| 1.0 | 0.03 | -0.31 | 0.36 | >0.05 |
| **Memory** | | | | |
| 0 | 0.16 | -0.18 | 0.51 | >0.05 |
| 0.2 | 0.16 | -0.18 | 0.51 | >0.05 |
| 0.4 | 0.16 | -0.18 | 0.51 | >0.05 |
| 0.6 | 0.16 | -0.18 | 0.51 | >0.05 |
| 0.8 | 0.16 | -0.18 | 0.51 | >0.05 |
| 1.0 | 0.16 | -0.18 | 0.51 | >0.05 |
| **Processing speed** | | | | |
| 0 | 0.17 | -0.16 | 0.51 | >0.05 |
| 0.2 | 0.17 | -0.16 | 0.51 | >0.05 |
| 0.4 | 0.17 | -0.16 | 0.51 | >0.05 |
| 0.6 | 0.17 | -0.16 | 0.51 | >0.05 |
| 0.8 | 0.17 | -0.16 | 0.51 | >0.05 |
| 1.0 | 0.17 | -0.16 | 0.51 | >0.05 |
| **Executive functioning** | | | | |
| 0 | 0.07 | -0.19 | 0.33 | >0.05 |
| 0.2 | 0.07 | -0.19 | 0.33 | >0.05 |
| 0.4 | 0.07 | -0.19 | 0.33 | >0.05 |
| 0.6 | 0.07 | -0.19 | 0.33 | >0.05 |
| 0.8 | 0.07 | -0.19 | 0.33 | >0.05 |
| 1.0 | 0.07 | -0.19 | 0.33 | >0.05 |
| **Language** | | | | |
| 0 | 0.14 | -0.15 | 0.44 | >0.05 |
| 0.2 | 0.14 | -0.15 | 0.44 | >0.05 |
| 0.4 | 0.14 | -0.15 | 0.44 | >0.05 |

| | | | | |
|---|---|---|---|---|
| 0.6 | 0.14 | -0.15 | 0.44 | >0.05 |
| 0.8 | 0.14 | -0.15 | 0.44 | >0.05 |
| 1.0 | 0.14 | -0.15 | 0.44 | >0.05 |
| **Motor speed** | | | | |
| 0 | 0.15 | -0.29 | 0.59 | >0.05 |
| 0.2 | 0.15 | -0.29 | 0.59 | >0.05 |
| 0.4 | 0.15 | -0.29 | 0.59 | >0.05 |
| 0.6 | 0.15 | -0.29 | 0.59 | >0.05 |
| 0.8 | 0.15 | -0.29 | 0.59 | >0.05 |
| 1.0 | 0.15 | -0.29 | 0.59 | >0.05 |
| **Visuospatial** | | | | |
| 0 | 0.35 | 0.06 | 0.64 | 0.02 |
| 0.2 | 0.35 | 0.06 | 0.64 | 0.02 |
| 0.4 | 0.35 | 0.06 | 0.64 | 0.02 |
| 0.6 | 0.35 | 0.06 | 0.64 | 0.02 |
| 0.8 | 0.35 | 0.06 | 0.64 | 0.02 |
| 1.0 | 0.35 | 0.06 | 0.64 | 0.02 |

Legend: RVE = robust variance estimation; ρ = the assumed within-study correlation among multiple effect sizes in the RVE model; SMD = Hedge’s standardized mean difference; *p* = the statistical significance of the pooled effect.

**Table S10: SMD meta-analysis outliers**

| Trimmed proportion | No. of effect sizes included | No. of studies included | SMD | Lower 95% CI | Upper 95% CI |
|---|---|---|---|---|---|
| **Global cognition** | | | | | |
| Trim 2.5%tails | 27 | 21 | 0.64 | 0.33 | 0.94 |
| Trim 5%tails | 25 | 20 | 0.57 | 0.29 | 0.85 |
| Trim 10%tails | 23 | 18 | 0.55 | 0.30 | 0.80 |
| Full data | 29 | 22 | 0.73 | 0.37 | 1.09 |
| **Attention** | | | | | |
| Trim 2.5%tails | 20 | 11 | 0.03 | -0.21 | 0.28 |
| Trim 5%tails | 18 | 10 | 0.14 | -0.01 | 0.29 |
| Trim 10%tails | 16 | 10 | 0.15 | 0.00 | 0.30 |
| Full data | 22 | 13 | 0.03 | -0.27 | 0.32 |
| **Memory** | | | | | |
| Trim 2.5%tails | 41 | 16 | 0.17 | -0.07 | 0.41 |
| Trim 5%tails | 39 | 15 | 0.10 | -0.09 | 0.29 |
| Trim 10%tails | 35 | 15 | 0.11 | -0.07 | 0.29 |
| Full data | 45 | 17 | 0.17 | -0.15 | 0.48 |
| **Processing speed** | | | | | |
| Trim 2.5%tails | 9 | 9 | 0.21 | -0.01 | 0.42 |
| Trim 5%tails | 9 | 9 | 0.21 | -0.01 | 0.42 |
| Trim 10%tails | 9 | 9 | 0.21 | -0.01 | 0.42 |
| Full data | 11 | 11 | 0.17 | -0.13 | 0.47 |
| **Executive functioning** | | | | | |
| Trim 2.5%tails | 23 | 15 | 0.11 | -0.10 | 0.32 |
| Trim 5%tails | 21 | 14 | 0.16 | -0.03 | 0.34 |
| Trim 10%tails | 19 | 12 | 0.17 | -0.00 | 0.32 |
| Full data | 25 | 16 | 0.07 | -0.16 | 0.31 |
| **Language** | | | | | |
| Trim 2.5%tails | 19 | 11 | 0.18 | 0.01 | 0.36 |
| Trim 5%tails | 19 | 11 | 0.18 | 0.01 | 0.36 |
| Trim 10%tails | 17 | 9 | 0.18 | 0.00 | 0.35 |
| Full data | 21 | 12 | 0.15 | -0.12 | 0.41 |
| **Motor speed** | | | | | |
| Trim 2.5%tails | 17 | 4 | 0.01 | -0.25 | 0.27 |
| Trim 5%tails | 17 | 4 | 0.01 | -0.25 | 0.27 |
| Trim 10%tails | 15 | 4 | 0.01 | -0.25 | 0.27 |
| Full data | 19 | 5 | 0.14 | -0.14 | 0.43 |
| **Visuospatial** | | | | | |
| Trim 2.5%tails | 12 | 7 | 0.28 | 0.10 | 0.45 |
| Trim 5%tails | 12 | 7 | 0.28 | 0.10 | 0.45 |
| Trim 10%tails | 10 | 7 | 0.25 | 0.07 | 0.43 |
| Full data | 14 | 8 | 0.35 | 0.11 | 0.59 |

Legend: CI = confidence interval; SMD = (pooled) standardized mean difference.

**Table S11: Egger's regression test for publication bias in interventional studies**

| Domain | Number of studies | Standard Egger test | | Cluster-robust Egger test | | |
|---|---|---|---|---|---|---|
| | | **z** | **p-value** | **Intercept** | **SE** | **p-value** |
| Global cognition | 22 | 0.69 | 0.49 | 0.89 | 1.77 | 0.63 |
| Attention | 13 | -0.90 | 0.37 | -1.80 | 2.34 | 0.48 |
| Memory | 17 | 0.74 | 0.46 | -0.30 | 3.10 | 0.93 |
| Processing speed | 11 | 0.78 | 0.44 | 5.12 | 5.85 | 0.42 |
| Executive functioning | 16 | -0.36 | 0.72 | -1.83 | 2.99 | 0.56 |
| Language | 12 | -2.13 | 0.03 | -7.63 | 4.62 | 0.15 |
| Motor speed | 5 | -0.32 | 0.75 | -1.80 | 4.69 | 0.74 |
| Visuospatial | 8 | -3.90 | <0.001 | -8.77 | 9.48 | 0.43 |

Legend: Standard Egger test = conventional Egger's regression approach, z = test statistic used to evaluate funnel plot asymmetry; Cluster-robust Egger test = Egger's regression model with cluster-robust standard errors, SE = the standard error of the intercept.

**Figure S1: Inclusion flow diagram**

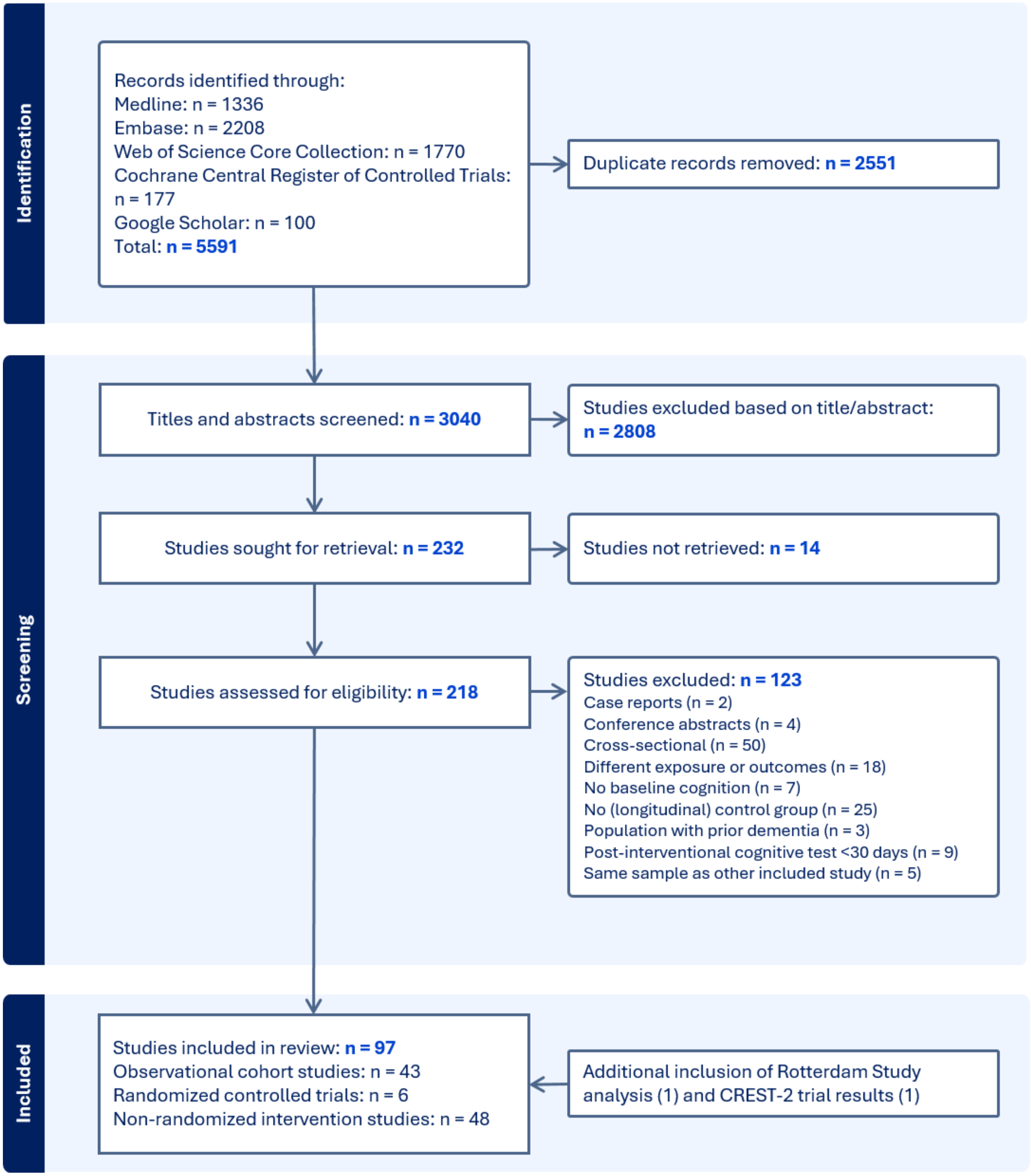

**Figure S2: Association of carotid atherosclerosis with all-cause dementia, other exposures**

A: Carotid IMT (dichotomous)

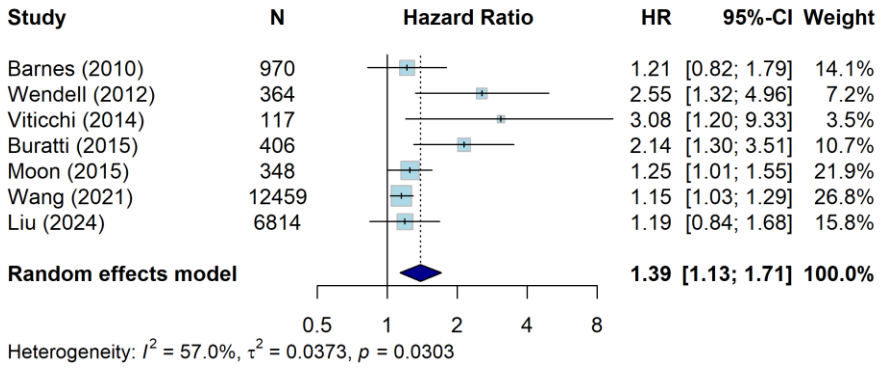


B: Carotid IMT (highest quantile vs lowest)

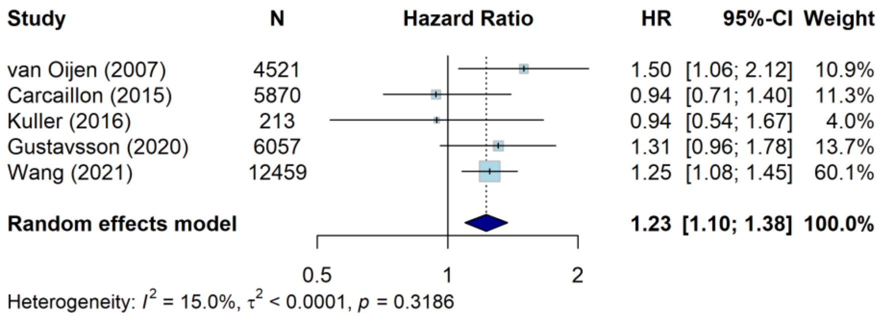


C: Carotid stenosis (dichotomous)

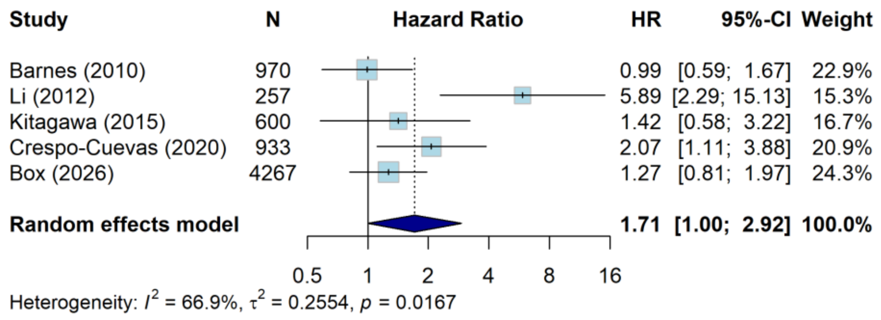


Legend: N = number of participants; HR = hazard ratio; CI = confidence interval, IMT = intimal medial thickness. A: The HR of Barnes (2010)[17] was based on an internal meta-analysis of the HRs for internal carotid IMT and common carotid IMT. The studies of Buratti (2015)[22] and Liu (2024)[32] had Alzheimer's disease as outcome instead of all-cause dementia. The studies by Buratti (2015)[22] and Viticchi (2014)[21] were performed on a population of MCI patients, the others on the general population. The study by Buratti (2015)[22] only controlled for breath-holding index, carotid plaque and hypertension and was therefore deemed of poor quality. The study by Moon (2015)[25] used logistic regression to assess the combined outcome of MCI and dementia. B: Carcaillon (2015)[23] and Gustavsson (2020)[30] compared the highest quartile to the lowest quartile, the studies by van Oijen

(2007)[15] and Wang W. (2021)[31] compared the highest quintile to the lowest quintile. C: The HR for the study by Li (2012)[18] was based on internal meta-analysis of HRs for moderate (30-69%) stenosis and severe (≥70%) stenosis relative to none-to-mild (0-29%) stenosis in hospitalized patients with MCI. The study by Barnes (2010)[17] used a cut-off of 25% stenosis in the general population, the study by Crespo-Cueva (2020)[29] a cut-ff of 50% stenosis in people from the general population with moderate to high vascular risk and Kitagawa (2015)[24] used a cut-off of 50% stenosis in an out-patient clinic population with at least one vascular risk factor. The study by Crespo-Cuevas[29] used a combination of MCI and dementia as outcome. The study by Box (2026) was conducted in the general population.

**Figure S3: Association of carotid atherosclerosis with Alzheimer's dementia**

A: Carotid IMT as a continuous measure

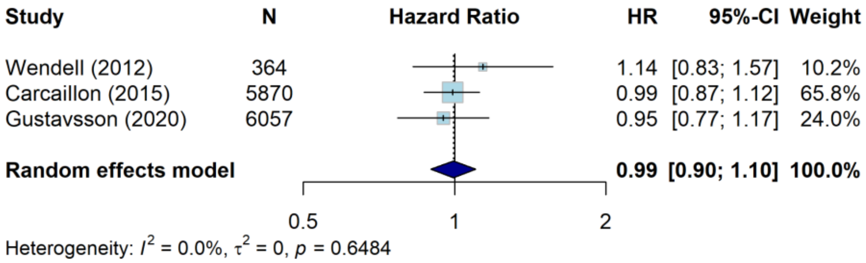


B: Carotid IMT as a dichotomised measure

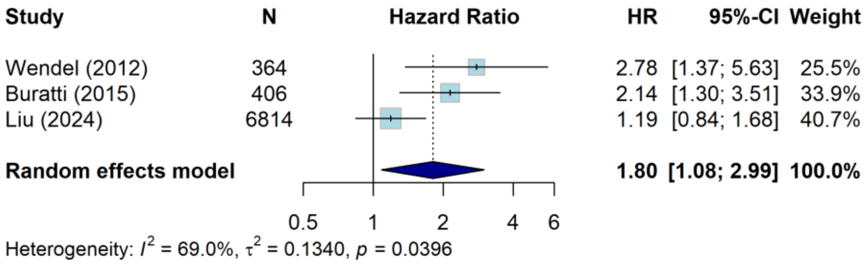


C: Carotid IMT highest quantile vs lowest

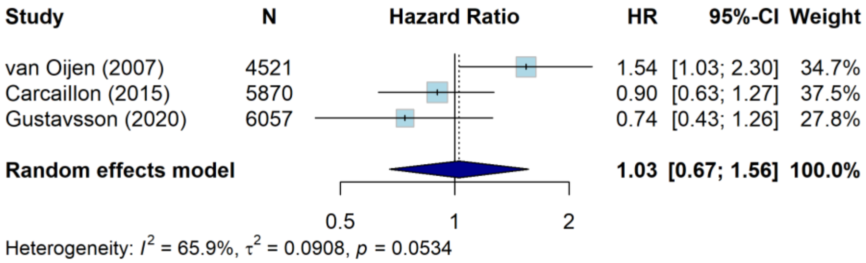


D: Carotid plaque presence (dichotomous)

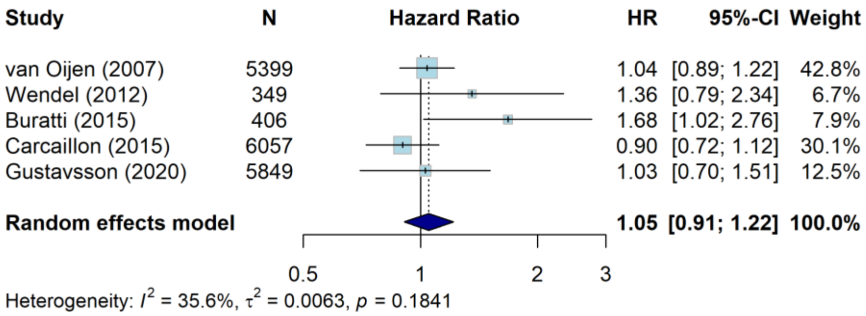


E: Carotid stenosis (dichotomous)

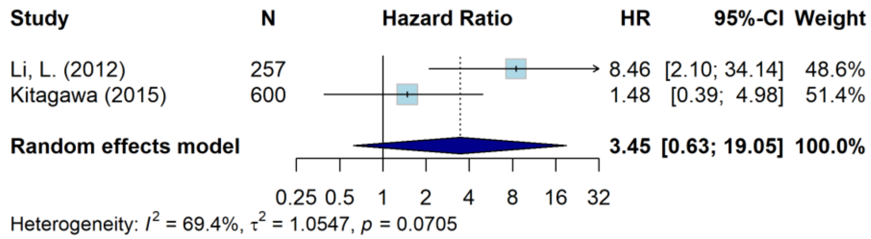


Legend: A: The HR for the study by Wendell (2012)[20] was adjusted to represent IMT per SD instead of IMT per 0.1mm. B: The study by Buratti (2015)[22] was performed on a population of MCI patients, the others on the general population. The study by Buratti (2015)[22] only controlled for breath-holding index, carotid plaque and hypertension. C: The studies by Carcaillon (2015)[23] and Gustavsson (2020)[30] compared the highest quartile to the lowest quartile, the study by van Oijen (2007)[15] compared the highest quintile to the lowest quintile. D: The study by Buratti (2015)[22] used a dichotomized plaque score, comparing no or one small plaque to multiple or one medium/large plaque. The HR for the studies by Carcaillon (2015),[23] van Oijen (2007),[15] and Wendell (2012)[20] were based on internal meta-analyses of multiple categories for plaque presence in order to get a dichotomous measure of plaque presence. E: The study by Li (2012)[18] compared moderate (30-69%) stenosis to mild (0-29%) stenosis in hospitalized patients with MCI. The study by Kitagawa (2015)[24] used a cut-off of 50% stenosis in an out-patient clinic population with at least one vascular risk factor.

**Figure S4: Association of carotid atherosclerosis with vascular or mixed dementia**

A: Carotid IMT as a continuous measure

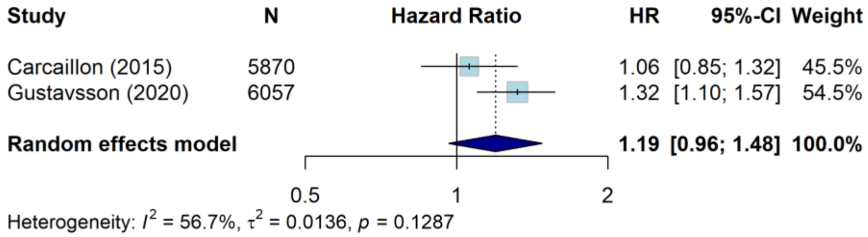


B: Carotid IMT highest quantile vs lowest

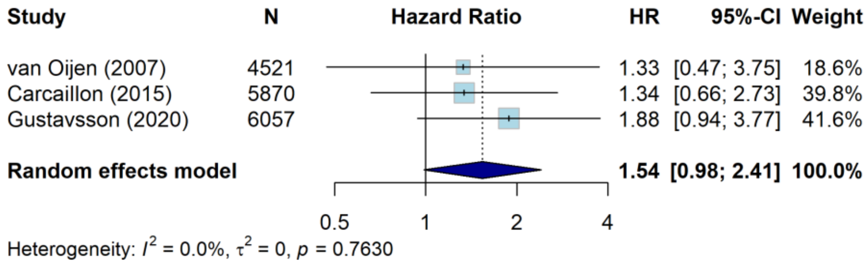


C: Carotid plaque presence (dichotomous)

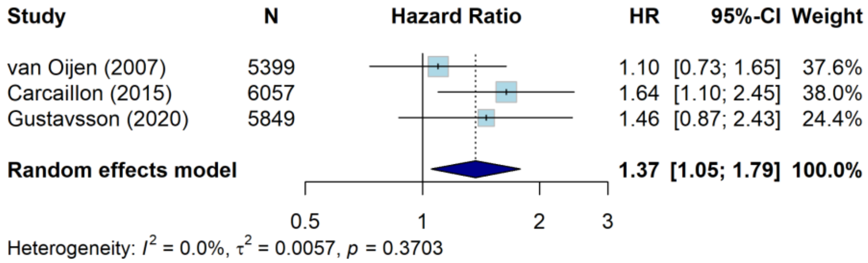


Legend: B: The studies by Carcaillon (2015)[23] and Gustavsson (2020)[30] compared the highest quartile to the lowest quartile, the study by van Oijen (2007)[15] compared the highest quintile to the lowest quintile. C: The HR for the studies by Carcaillon (2015)[23] and van Oijen (2007)[15] were based on internal meta-analyses of multiple categories for plaque presence in order to get a dichotomous measure of plaque presence. There were no studies that investigated carotid IMT as a dichotomous exposure and only one study that investigated the association between carotid stenosis and vascular dementia: Kitagawa (2015)[24] found a HR of 1.37 (95%CI: 0.33–4.92) for ≥50% stenosis vs <50%.

**Figure S5: Effect of CEA/CAS on change in global cognition in randomized controlled trials using Hartung-Knapp adjustment**

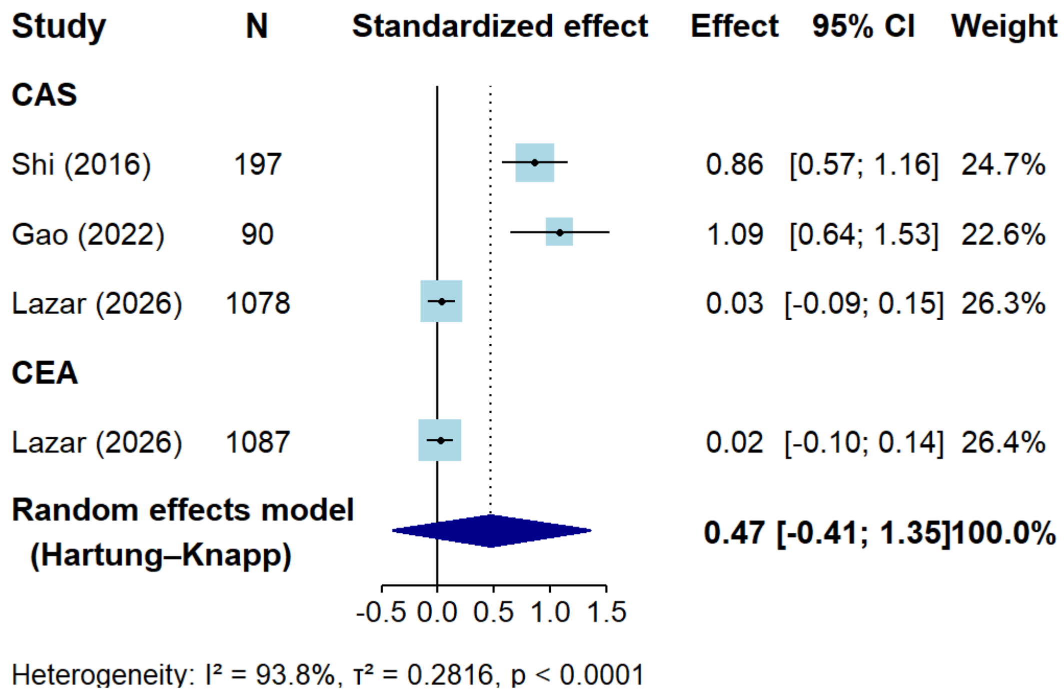


Legend: N = number of participants; CI = confidence interval; CEA = carotid endarterectomy; CAS = carotid artery stenting.

**Figure S6: Effect of carotid revascularization on change in global cognition, subgroup analyses by stenosis status of reference group.**

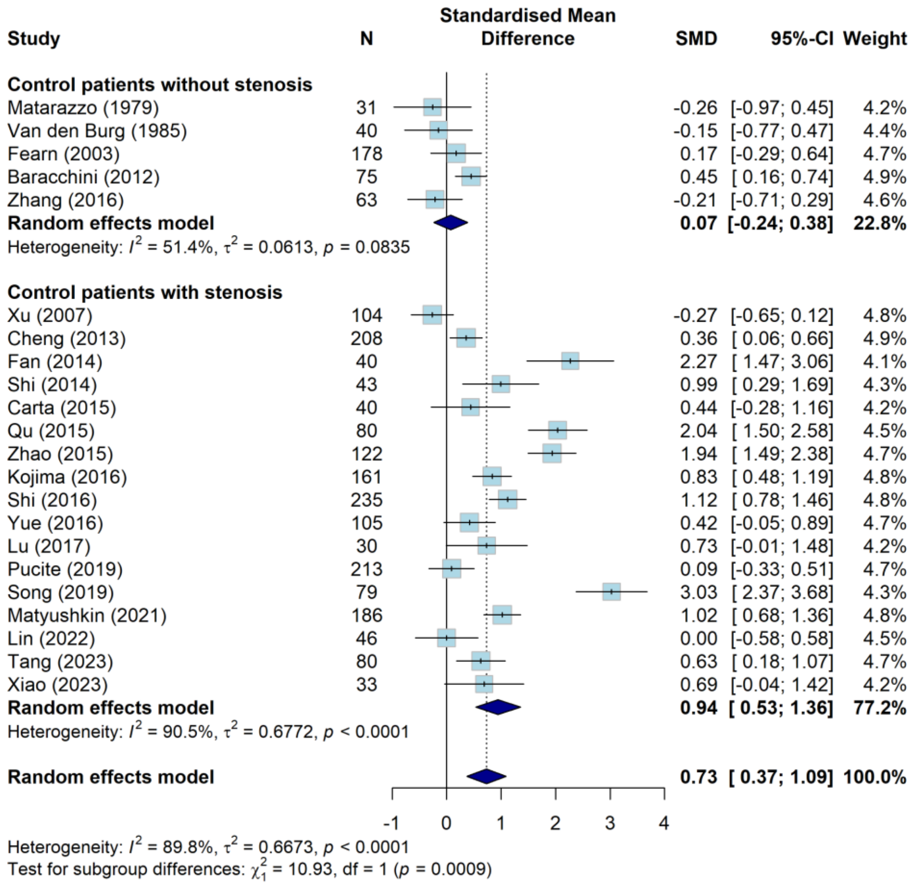


Legend: Non_stenosis = reference groups without carotid stenosis, including healthy participants or patients undergoing other types of surgery. Stenosis = reference groups with carotid stenosis comparable to the intervention group but did not undergo CEA/CAS. N = Number of studies; SMD = standardized mean difference; CI = confidence interval.

**Figure S7: Effect of carotid revascularization on change in global cognition, subgroup analyses by symptom status**

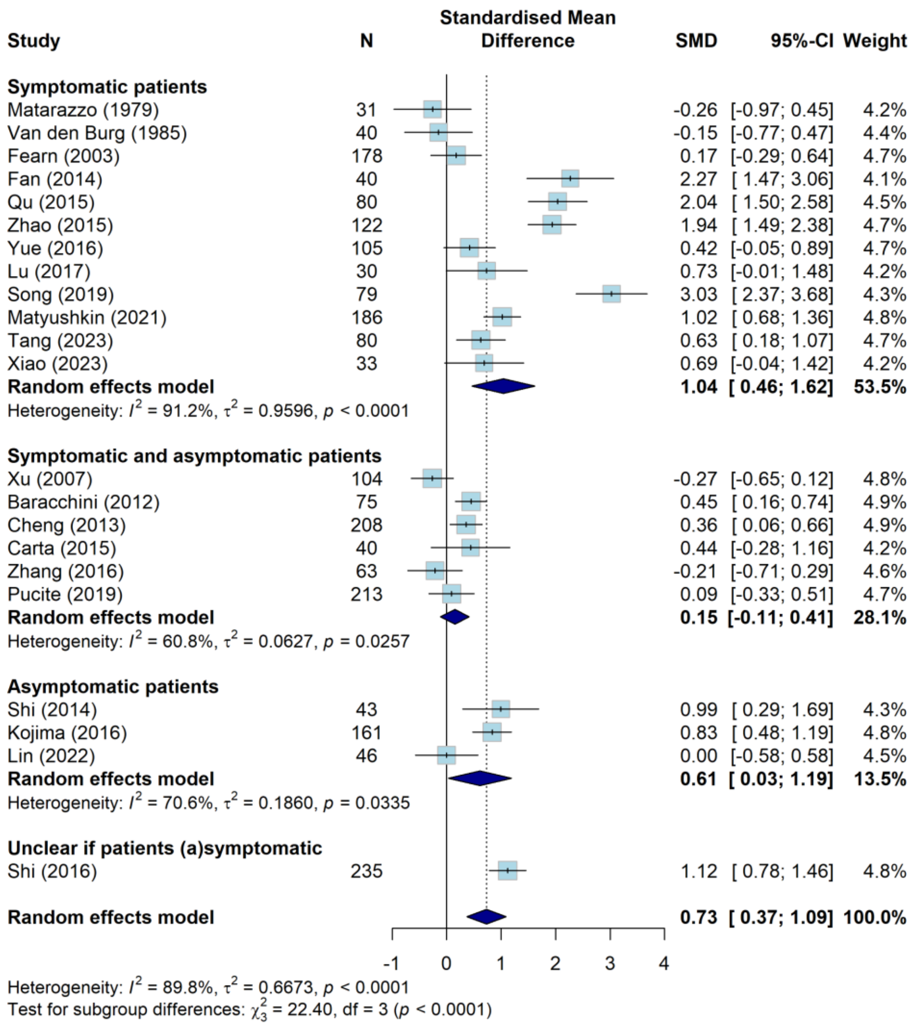


Legend: Both = the intervention group included both symptomatic and asymptomatic participants; unclear = symptom status was not sufficiently reported. N = Number of studies; SMD = standardized mean difference; CI = confidence interval.

**Figure S8: Effect of carotid revascularization on change in domain-specific cognitive performance**

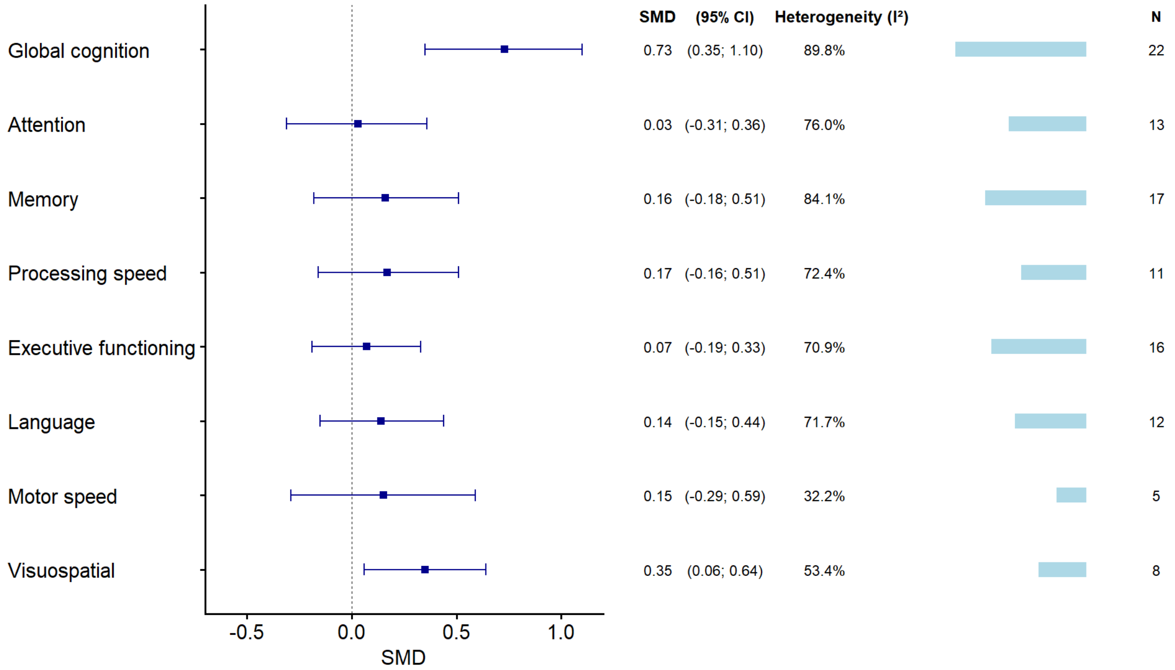


Legend: N = Number of studies; SMD = standardized mean difference; CI = confidence interval.

**Figure S9: Effect of CEA/CAS on change in domain-specific cognitive performance**

A: Effect of CEA/CAS on change in Visuospatial

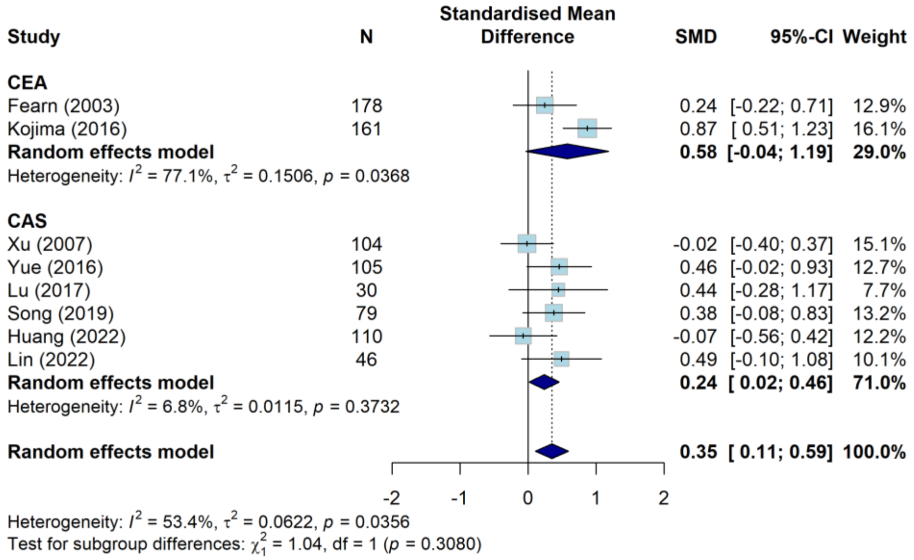


B: Effect of CEA/CAS on change in processing speed

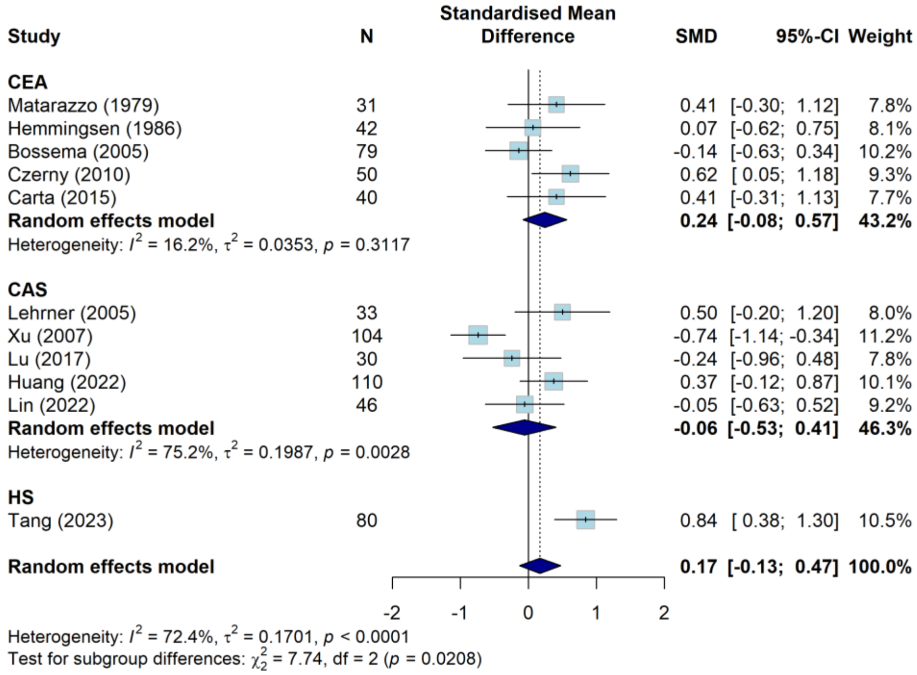


C: Effect of CEA/CAS on change in motor speed

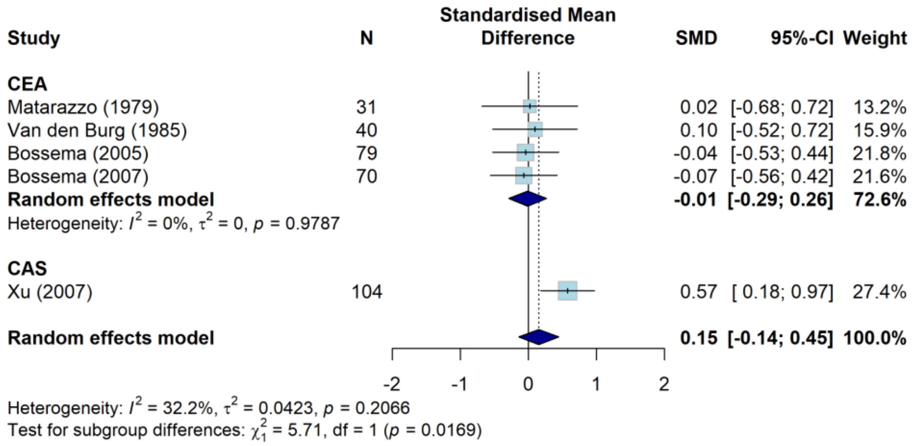


D: Effect of CEA/CAS on change in memory

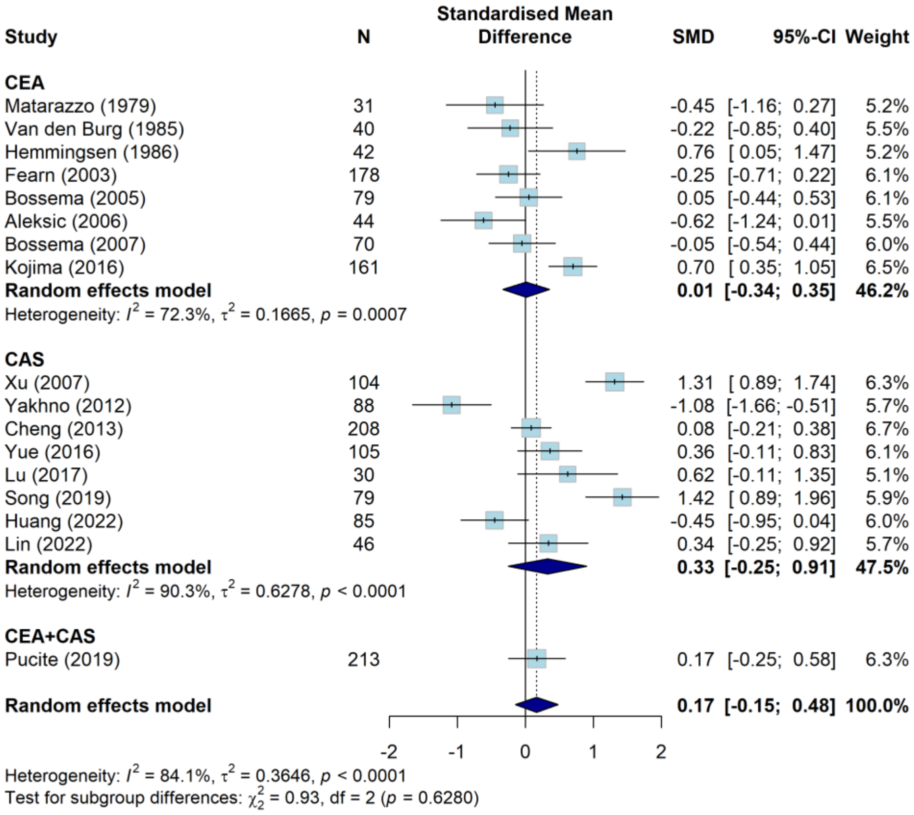


E: Effect of CEA/CAS on change in language

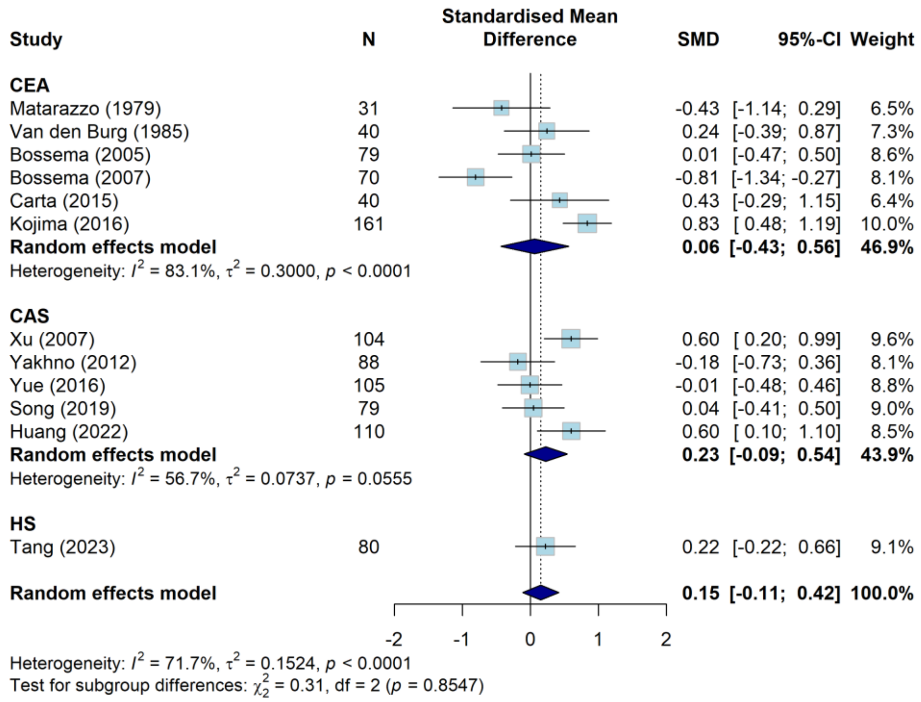


F: Effect of CEA/CAS on change in executive functioning

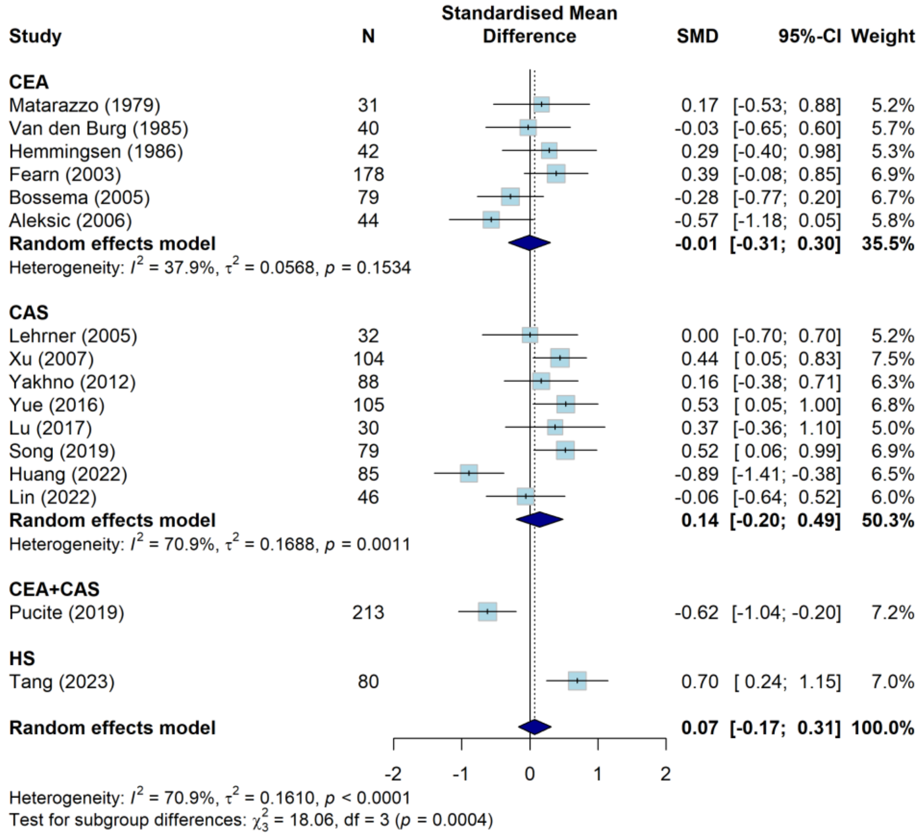


G: Effect of CEA/CAS on change in attention

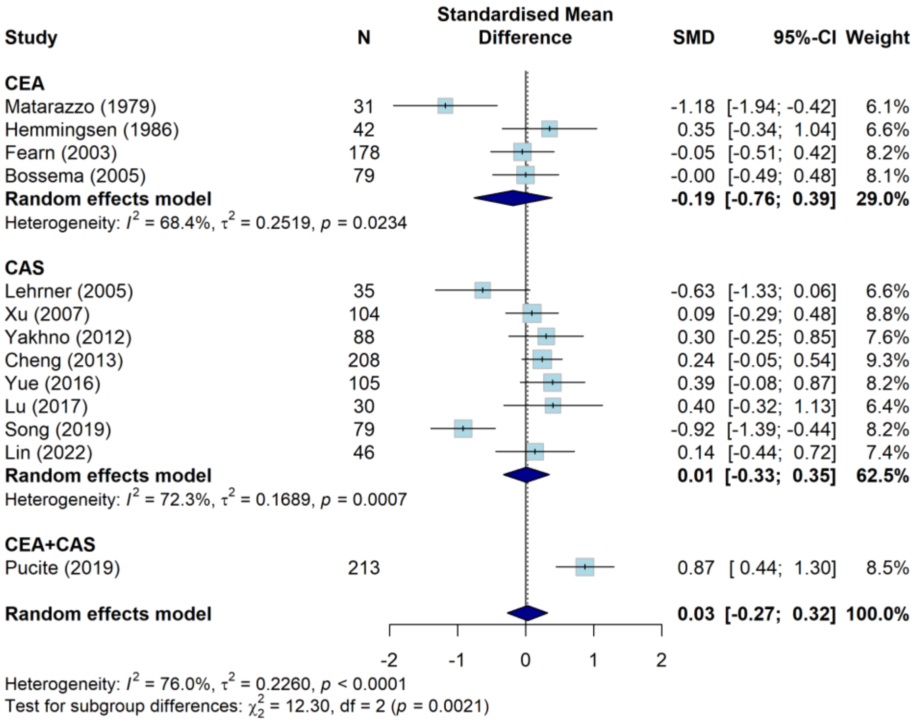


Legend: N = number of participants; SMD = standardized mean difference; CI = confidence interval; CEA = carotid endarterectomy; CAS = carotid artery stenting; CEA+CAS = mixed intervention group including participants who underwent either CEA or CAS; HS = hybrid surgery.

**Figure S10: Funnel plots assessing publication bias in global cognition and domain-specific cognitive performance after carotid revascularization**

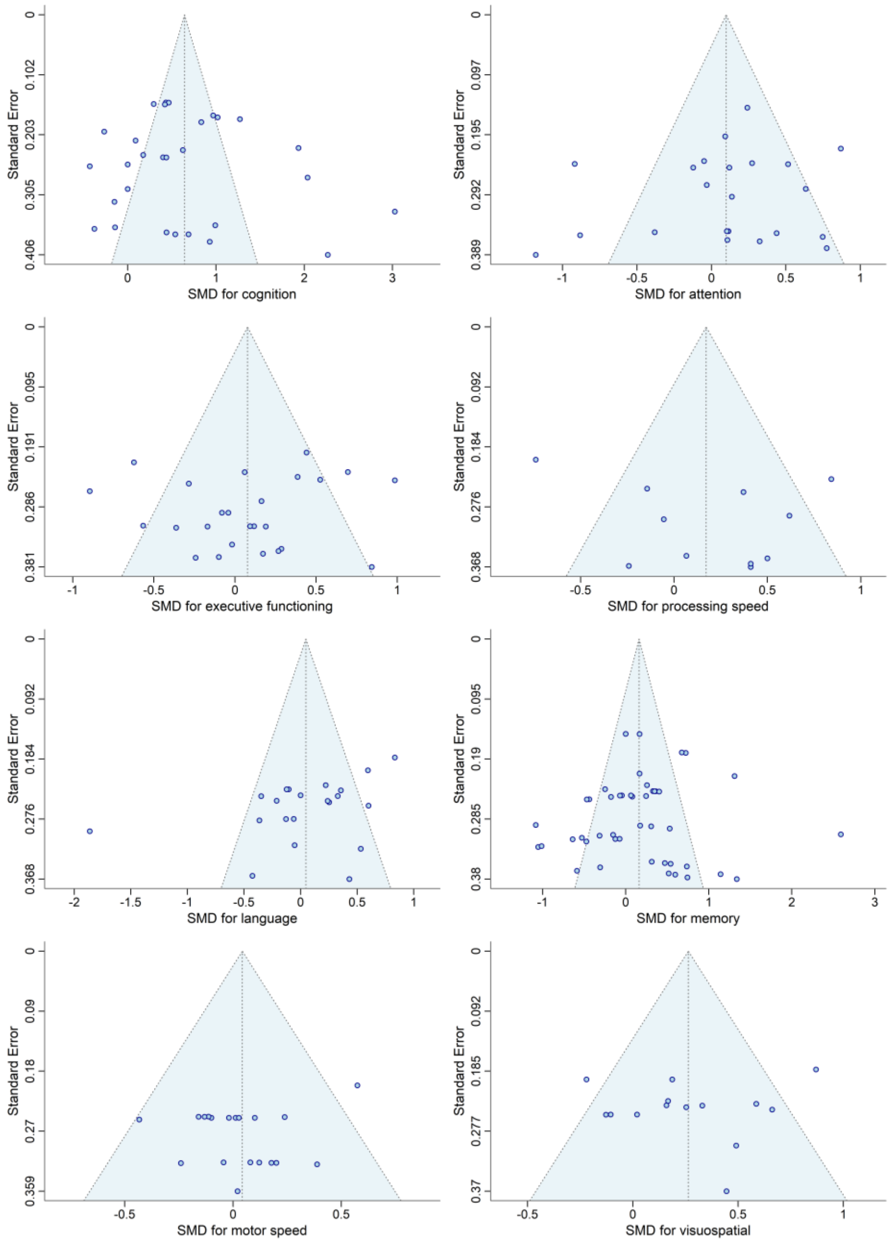


Legend: SMD = standardized mean difference.
